# LinAge2: Providing actionable insights and benchmarking with epigenetic clocks

**DOI:** 10.1101/2024.12.23.24319587

**Authors:** Sheng Fong, Kirill A. Denisov, Brian K. Kennedy, Jan Gruber

**Affiliations:** Population Health Research Office, Ng Teng Fong General Hospital, Singapore; Department of Medicine (Geriatric Medicine), Ng Teng Fong General Hospital, Singapore; Gero PTE, Paya Lebar Square, Singapore; Healthy Longevity Translational Research Program, Yong Loo Lin School of Medicine, National University of Singapore, Singapore; Center for Healthy Longevity, National University Health System, Singapore; Department of Biochemistry, Yong Loo Lin School of Medicine, National University of Singapore, Singapore; Department of Physiology, Yong Loo Lin School of Medicine, National University of Singapore, Singapore

**Author notes:** Corresponding author: (JG).

## Abstract

Biological aging is marked by a decline in resilience at the cellular and systemic levels, driving an exponential increase in mortality risk. Here, we evaluate several clinical and epigenetic clocks for their ability to predict mortality, demonstrating that clocks trained on survival and functional aging outperform those trained on chronological age. We present an enhanced clinical clock that predicts mortality more accurately and provides actionable insights for guiding personalized interventions. These findings highlight the potential of mortality-predicting clocks to inform clinical decision-making and promote strategies for healthy longevity.

## Main

Biological aging is characterized by the progressive decline in intrinsic biological resilience that is associated with an exponential increase in mortality, expressed in the demographic “Gompertz mortality law”^1^. Not all humans age at the same rate since genetics, lifestyle and stochastic factors can affect future mortality and morbidity trajectories. Consequently, individual true biological age (BA) is not identical to calendar or chronological age (CA). The true BA of an individual can be uniquely defined as the age at which subjects of a reference cohort have the same risk of age-dependent disease and all-cause mortality as the subject in question. Tools to accurately track changes in true BA are essential for the development and validation of novel life- and healthspan-optimizing diet, lifestyle, supplement and drug interventions.

Biological aging “clocks” are computational tools that estimate individual true BA based on demographic, clinical, and/or molecular data. CA itself is widely used for both clinical prognostication and decision-making, and can be viewed as a first order approximation of true BA. The ideal BA clock should predict individual Gompertz mortality risk with higher accuracy than CA. Some aging clocks, including most clinical clocks, explicitly include CA as a covariate, using biological features to estimate a correction factor aimed at providing a better estimate of true BA. CA in this case is used as a proxy for effects and mechanisms, such as entropic damage, not captured by the clock itself. Of course, the ideal clock would include all relevant processes, wherein the model would assign zero or negligible weight to CA.

Aging clocks are generalizations of current clinical risk markers that predict disease-specific morbidity and, in some cases, mortality. Aging clocks should similarly enable early detection of hidden or subclinical diseases, surpassing the capabilities of diagnostics by identifying disease processes years or decades before overt disease is present. Secondly, to inform risk-to-benefit estimates (clinical equipoise), aging clocks should capture all-cause mortality holistically, providing value beyond organ or disease-specific risks. Thirdly, aging clocks must be sensitive to individual variations in biological resilience. Finally, aging clocks should provide tools for mechanistic interpretation and provide actionable insights, facilitating targeted interventions. To date, none of the existing clocks meet all these criteria.

To evaluate the performance of aging clocks, we can compare them to a hypothetical “ideal” clock, which we term “CrystalAge”. This optimal clock would predict disease-specific and all-cause mortality at the individual level with near-perfect accuracy, essentially forecasting an individual’s date of death (Fig. 1a and d). While practically impossible, in retrospective studies, we can determine the theoretically optimal performance of CrystalAge and use it as a benchmark to evaluate the performance of existing aging clocks.

**Fig. 1:**
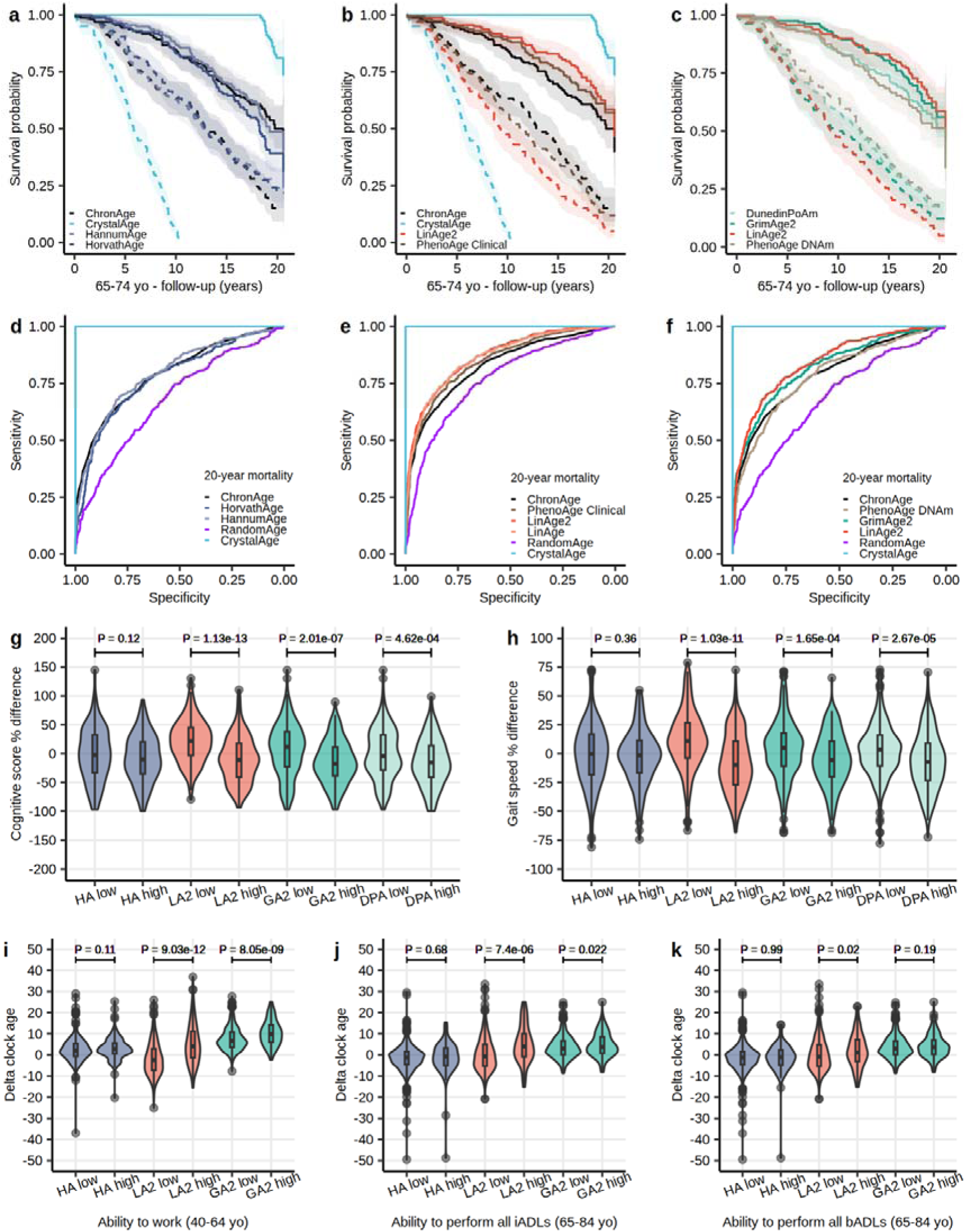
LinAge2 predicts 20-year all-cause mortality and tracks with healthspan markers. **a-c,** Kaplan-Meier survival curves showing 20-year survival in the 65-74 CA bin (*n*=631). For each clock, subjects were stratified by selecting the lowest (best, solid line) and highest (worst, dotted line) 25% quartiles for BA. Clocks within the same quartile were compared using log-rank tests with Benjamini-Hochberg correction. Areas shaded indicate 95% error bands for lines of the same color. **b,** Compared to ChronAge, use of LinAge2 BA results in a significant survival difference for the lowest 25% BA quartile (*P*=6.16E-04), but not for the highest 25% quartile (*P*=0.07). PhenoAge Clinical did not significantly outperform ChronAge in predicting survival in this age bin. **c,** LinAge2 significantly outperformed DunedinPoAm (*P*=1.09E-02) and PhenoAge DNAm (*P*=1.37E-03) in the lowest 25% BA quartile, but not GrimAge2 (*P*=0.22). In the highest 25% quartile, while LinAge2 significantly outperformed PhenoAge DNAm (*P*=0.03), the differences between LinAge2 and DunedinPoAm (*P*=0.11) and GrimAge2 (*P*=0.58) did not reach statistical significance. **e,** ROC analysis revealed that LinAge2 (area under the curve (AUC)=0.8684) was significantly more informative than PhenoAge Clinical (AUC=0.8479, *P*=6.35E-05) and ChronAge (AUC=0.8288, *P*=3.16E-10) in predicting future mortality (*n*=2,036). LinAge2 performed similarly to LinAge (AUC=0.8647). **f,** LinAge2 also outperformed PhenoAge DNAm (AUC=0.7859, *P*=4.44E-07) and GrimAge2 (AUC=0.8233, *P*=0.02) in predicting 20-year mortality (*n*=1,065). Although GrimAge2 outperformed ChronAge (AUC=0.7933, *P*=2.74E-03) in predicting 20-year mortality, PhenoAge DNAm did not (*P*=0.47). **a,d,** HorvathAge, HannumAge, and ChronAge did not significantly differ in predicting mortality risk (AUCs=0.7776, 0.7978 and 0.7933, respectively, *n*=1,065). ROC curves were compared using DeLong’s test. **a,b,d,e,f,** CrystalAge, a theoretical perfect clock shown for reference, accurately identifies individuals at risk of dying (AUC=1), whereas RandomAge adds random gaussian noise of ±10 years to CA. **g-k,** Violin plots for each clock categorized into low (biologically younger/best 25% quartile) and high (biologically older/worst 25% quartile) groups plotted against healthspan markers: cognitive scores (digit symbol substitution test), gait speed, ability to work, and ability to perform all instrumental and basic activities of daily living (iADLs and bADLs). Groups (BA high versus low) were compared using two-sided t-tests. Median value, lower (25^th^) and upper (75^th^) percentiles are indicated. Lines extend to ±1.5 times interquartile range, with points outside this range drawn individually. The violin shape indicates the probability density function. yo, years old. HA, HorvathAge. LA2, LinAge2. GA2, GrimAge2. DPA, DunedinPoAm.

Taking inspiration from Levine’s PhenoAge clinical clock^2^, we recently developed and validated clinical aging clocks (PCAge, LinAge) based on linear dimensionality reduction by matrix factorization (singular value decomposition) and demonstrated them to be highly predictive in terms of future disease-specific and all-cause mortality^3^. These clocks have since been applied in a range of clinical settings and, taking advantage of user feedback, we have implemented several improvements, creating an updated version of these clocks (LinAge2). Like LinAge, we trained LinAge2 in the National Health and Nutrition Examination Survey (NHANES) IV 1999-2000 wave before testing it in the 2001-2002 wave. LinAge2 further reduces the number of rarely measured parameters and emphasizes interpretability. For a detailed description of LinAge2’s features and construction, refer to Methods.

Many aging clocks have been developed, with epigenetic or DNA methylation (DNAm) clocks most widely recognized and well-established. Several epigenetic clocks have been commercially licensed for applications, including estimating CA (HorvathAge^4^, HannumAge^5^), optimizing life insurance policies (PhenoAge DNAm^6^, GrimAge^7^), and monitoring the rate of aging (DunedinPoAm^8^)^9^. Recently, a dataset of pre-calculated epigenetic clock ages has been published for the NHANES 1999-2002 waves, permitting direct comparison of the predictive power of CA, the original LinAge, LinAge2 and PhenoAge clinical clocks, and the HorvathAge, HannumAge, PhenoAge DNAm, GrimAge2^10^ and DunedinPoAm epigenetic clocks.

To compare efficacy in predicting mortality, we performed survival and receiver operating characteristic (ROC) analyses on 20- and 10-year mortality in the NHANES 2001-2002 test cohort. Compared to CA, LinAge2 demonstrated significant survival differences across all age bins, whereas PhenoAge Clinical did not (Fig. 1b, Extended Data Fig. 1b and e). LinAge2 performed similarly to LinAge and demonstrated superior predictive power for future mortality compared to PhenoAge Clinical and CA (Fig. 1e and Extended Data Fig. 2b). Surprisingly, LinAge2 also outperformed PhenoAge DNAm and DunedinPoAm in predicting age-specific survival differences (Fig. 1c, Extended Data Fig. 1c and f) and future mortality (Fig. 1f and Extended Data Fig. 2c). In contrast, PhenoAge DNAm, HorvathAge, and HannumAge did not significantly differ from CA in predicting future mortality (Fig. 1a, d and f, Extended Data Fig. 1a and d, and Extended Data Fig. 2a and c). LinAge2 and GrimAge2 performed similarly in predicting future mortality (Fig. 1f and Extended Data Fig. 2c) and survival across all age bins (Fig. 1c, Extended Data Fig. 1c and f).

While mortality prediction is an important function of aging clocks, it is important to evaluate if clock ages are similarly predictive of functional status and healthspan. We tested this for the same clinical and epigenetic clocks by comparing markers of functional and health status in individuals selected by each clock to be in the lowest 25% BA quartile (biologically younger, low) with those in the highest 25% quartile (biologically older, high). Our analysis revealed that LinAge2 low was associated with superior healthspan markers, including higher cognitive scores, faster gait speed, ability to work, and performance of all instrumental and basic activities of daily living (iADLs and bADLs) (Fig. 1g-k). Conversely, individuals in the LinAge2 high group had poorer healthspan, with statistically significant differences between the two groups across all markers (Fig. 1g-k). Similar trends were observed for GrimAge2 and DunedinPoAm, with statistically significant differences between the low and high groups across most healthspan markers, except for the ability to perform all bADLs (Fig. 1g-k and Extended Data Fig. 3). In contrast, no statistically significant differences were found between HorvathAge low and HorvathAge high across healthspan markers (Fig. 1g-k). Our findings on healthspan markers and mortality, for HorvathAge, HannumAge, PhenoAge DNAm and GrimAge2, corroborate similar findings for 10-year survival in 490 subjects of the Irish Longitudinal Study on Aging^11^.

A significant drawback of many existing aging clocks is that they lack interpretability and actionable insights, making it challenging to develop targeted interventions. However, one key benefit of clinical clocks is that they are built from parameters directly related to the underlying disease mechanisms, enabling easier interpretation of clock residuals and providing actionable insights into disease pathophysiology. Individual age-associated principal components (PCs) identify clusters of features that change in a coordinated manner during aging. Analyzing individual PCs can provide valuable insights into the underlying patterns and trajectories of aging-related changes. Using heatmaps, we visualized the predictive power of individual PCs relative to clinical outcomes, including sex-specific causes of death and chronic diseases (Fig. 2). Supplementary Table 5 provides detailed insights into the interpretation of each PC, including associations with causes of death, chronic diseases, lifestyle factors, and potential aging mechanisms, as well as suggested interventions to optimize each PC and lower BA.

**Fig. 2:**
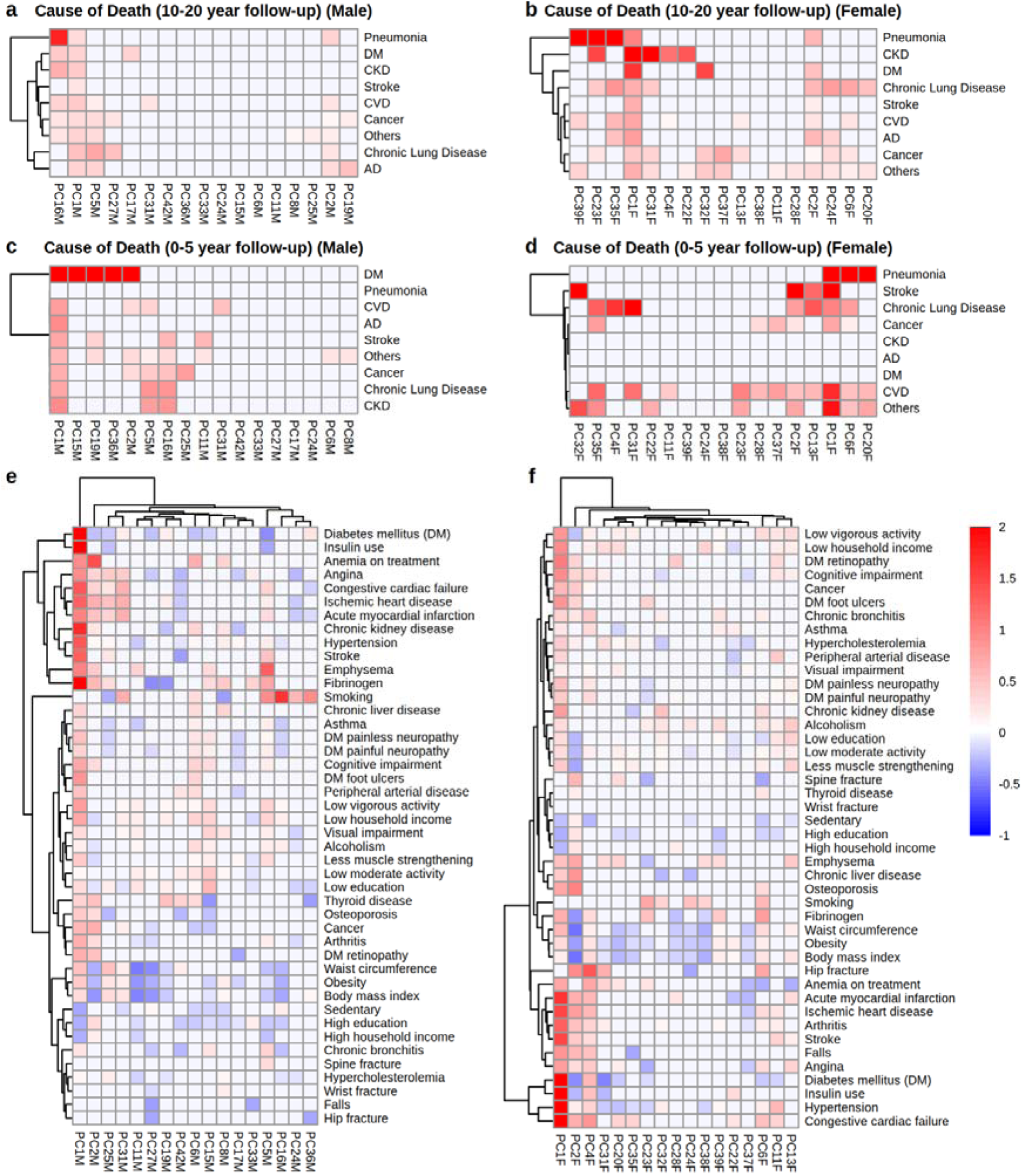
Heatmaps illustrating the associations between clinical outcomes and PCs analyzed using multivariate logistic regression. Associations of PCs with specific causes of death at **a-b,** 10-20 year and **c-d,** 0-5 year follow-up for male and females, respectively. Strength of association (odds ratio) is represented using a red color scale. PCs that are strongly positively associated with a specific cause of death are bright red. For cause of death, negative associations were truncated by setting their values to zero (i.e. no harm). **e-f,** Association of PCs with specific chronic diseases, and measures of lifestyle and socioeconomic status. Strength of association is represented using a blue-red scale ranging from −1 (blue, negative association) to 2 (red, positive association). PCs that are strongly positively associated (positive risk ratios) with a specific disease are bright red, whereas PCs that are strongly negatively associated (negative risk ratios) with a specific disease are bright blue.

Accurately predicting patient outcomes and allocating healthcare resources is a significant challenge in clinical practice^12^. Currently, clinicians rely heavily on CA to make these decisions. However, here we show that mortality-predicting clocks, such as LinAge2 and GrimAge2, outperform CA in predicting mortality risk across timeframes, ranging from 2-20 years (Fig. 1, Extended Data Fig. 4). Moreover, clinical clocks can also predict specific causes of death within a 5-year window (Fig. 2c and d). This illustrates that clock-based BAs are more accurate and informative estimates of true BA than CA itself. By providing a more precise metric of biological status than CA alone, BA can enable clinicians to better support patients and their caregivers in navigating healthcare choices, including end-of-life care.

Overall, our analysis reveals that, regardless of feature space (methylation or clinical), aging clocks trained to predict mortality or functional aging outcomes provide more predictive value in terms of clinical decision-making. Surprisingly, clinical aging clocks still outperform several prominent mortality-predicting and functional epigenetic clocks, including PhenoAge DNAm and DunedinPoAm, in predicting future mortality. A key advantage of LinAge2 lies in its interpretability. Because principal component analysis is a linear matrix factorization technique, the resulting model is easier to interpret than nonlinear alternatives^13^. Latent variables based on linear dimensionality reduction (PCs), especially those based on clinical parameters, are comparatively easy to understand and interpret, making them more actionable. This enables clinical aging clocks like LinAge2 to detect hidden or subclinical diseases and inform primordial prevention strategies. By identifying individuals at high risk of developing specific diseases, healthcare providers can implement targeted interventions early and proactively. By casting specific risk in terms of BA acceleration, aging clocks can significantly increase compliance and adherence with specific health recommendations^14^. For example, male smokers with high PC5M values in LinAge2 are at increased risk of death from chronic lung disease and should be screened and advised to quit smoking (Fig. 2 and Supplementary Table 5).

All current aging clocks, regardless of feature space (e.g. clinical, methylation, proteomics, etc.) and target (mortality, functional outcomes, disease, or CA) share significant limitations. Most importantly, many current clocks employ linear techniques (e.g. principal component analysis/singular value decomposition, regression-based predictions), which limit their ability to distinguish between aging signatures and those of age-dependent diseases, and to learn U-shape response patterns. Current clocks therefore inherently conflate intrinsic biological aging with disease-specific signatures (hidden sickness, primordial disease signatures). Nonlinear approaches including generative artificial intelligence and artificial neural networks are being investigated and could offer improved models, but their increased complexity pose a significant challenge for interpretation^15,16^. Next generation clocks will need to differentiate between disease signatures and intrinsic aging, and quantify intrinsic biological resilience. Further theoretical work will be required to deconvolute these disease-centric signatures from determinants of intrinsic resilience and entropic aging^17–19^. Advancing next generation clocks is crucial to equip healthcare providers with the essential tools needed to make informed decisions regarding targeted interventions that support healthy longevity in populations where healthcare needs are increasingly dominated by aging.

## Methods

### Motivation for enhancing LinAge2

The original PCAge and LinAge^3^ both utilized some parameters that are not routinely collected. LinAge has been utilized by several clinics worldwide, and we have received informal feedback regarding its use. Common suggestions for enhancing the clock include: (i) improving handling of outliers and threshold effects, (ii) further refining the clinical parameters, especially removing serum fibrinogen due to the need for a specialized sodium citrate tube, (iii) providing additional tools to improve the interpretability of PCs, and (iv) providing specific strategies to optimize each PC to lower BA. PCs can also be sensitive to outliers, thresholding and batch effects. We developed LinAge2 in response to these concerns.

We followed the same workflow, as previously described^3^, to construct LinAge2 but with several modifications. To enhance LinAge2, we refined the clinical parameters by reducing the total number to 60, removing serum fibrinogen (Supplementary Table 2). We also addressed outliers and thresholding by capping outliers at six standard deviations and log-transforming additional parameters (Supplementary Table 2). Batch effects were mitigated through z-score normalization by median and median absolute deviation to a younger, generally healthy cohort (age 40-50 years), separately for males and females (Supplementary Table 2), generating sex-specific PCs. The loadings for male and female PCs are provided in Supplementary Table 3, and sex-specific weights of the Cox proportional hazards models are listed in Supplementary Table 4. A parametrized version of LinAge2 is provided as previously described^3^ (Supplementary Table 2). The baseline characteristics of the study participants are listed in Supplementary Table 1.

### PhenoAge Clinical and epigenetic clocks

PhenoAge Clinical was implemented using the equation from the original publication. The dataset of pre-calculated epigenetic clock ages published for the NHANES 1999-2002 waves were obtained from https://wwwn.cdc.gov/nchs/nhanes/dnam/ and analyzed.

### Construction of healthspan markers

The digit symbol substitution test score (NHANES variable ‘CFDRIGHT’) was used as a cognitive measure. Gait speeds were obtained by taking the total distance walked (20 feet or 6.096 meters) divided by the time taken (NHANES variable ‘MSXWTIME’). Differences in cognitive scores and gait speeds were calculated as the percent difference between a control group (middle 50% of all subjects), for younger (best 25% quartile) and older (worst 25% quartile) groups. The ability to work was established using the NHANES variable ‘PFQ048’. The ability to perform all instrumental activities of daily living (iADLs) was a combination of the NHANES variables ‘PFQ060A’, ‘PFQ060F’, ‘PFQ060G’, ‘PFQ060Q’, PFQ060R’ and ‘PFQ060S’, while the ability to perform all basic activities of daily living (bADLs) was a combination of the NHANES variables ‘PFQ060B’, ‘PFQ060C’, ‘PFQ060H’, ‘PFQ060I’, ‘PFQ060J’, ‘PFQ060K’ and ‘PFQ060L’. Participants had to have either no difficulty or some difficulty in all the variables to be deemed able to perform all iADLs or all bADLs.

### Heatmap analysis

Using the ‘nnet’^20^ (version 7.3-19) R package, heatmaps were generated to evaluate the predictive values for PCs included in LinAge2. For each parameter, we attempted to predict status (diseased/compromised or not) using multivariate logistic regression with the clock PCs as covariates. PCs that received a statistically insignificant (*P*≥0.05) weight in the logistic regression model were assigned zero weights (white). The remaining PCs (*P*<0.05) were assigned color values according to their weight in the model (see Fig. 2f legend for color mapping).

### Statistics and reproducibility

For the NHANES IV 1999-2002 waves, we excluded: participants top-coded at age 85 years, as we could not ascertain the exact CAs of these adults, and participants who died from accidental deaths, as these were deemed to be not age-related.

Survival analyses were performed using log-rank tests with Benjamini-Hochberg correction. ROC curves were compared using DeLong’s test. For healthspan markers, two-sided t-tests were used to compare between the low and high clock groups. All statistical analyses were performed using R version 4.2.0 (https://www.R-project.org/).

## Supporting information

LinAge2_Supplementary_Information

## Data availability

All datasets used are publicly available online at https://wwwn.cdc.gov/nchs/nhanes/Default.aspx. There were no restrictions on data availability. This study was reported according to STROBE guidelines for cohort studies.

## Acknowledgements

We thank the National Health and Nutrition Examination Survey participants and staff who made this study possible. We thank C. Chen for her careful reading of this manuscript. This research was funded by the Ministry of Education in Singapore, grant numbers IG21-SG007 and A-0007215-00-00, to J.G. S.F. is supported by the Research Training Fellowship (MOH-001294-00) from the National Medical Research Council Singapore. This work was supported by the Lien Foundation.

## Author contributions

S.F., B.K.K. and J.G. conceived, conceptualized and designed the study. S.F., K.A.D. and J.G. analyzed and interpreted the data. S.F., B.K.K. and J.G. wrote the first draft of the paper.

## Additional information

**Extended Data Fig. 1:**
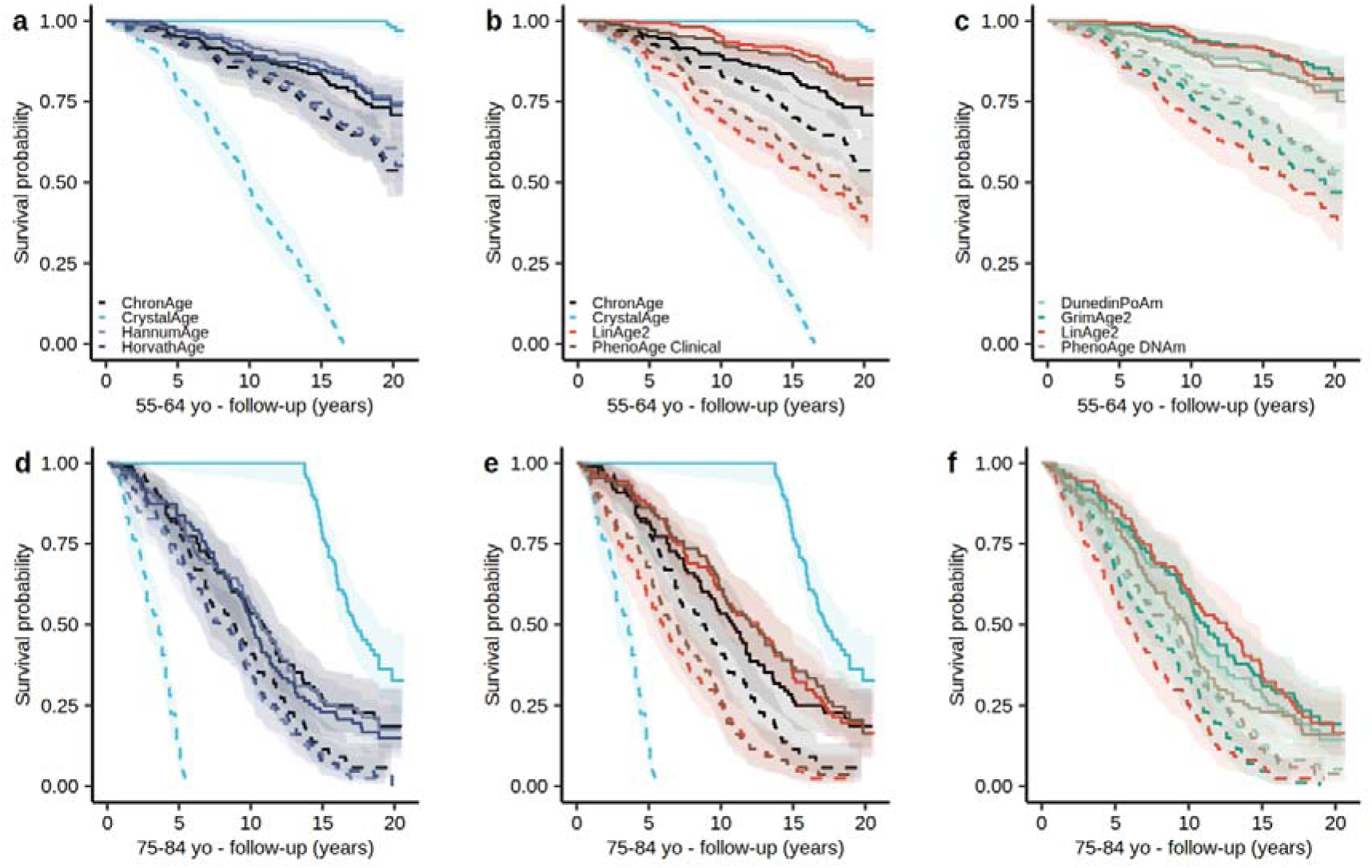
LinAge2 predicts survival in chronologically 55-64 and 75-84 year old individuals. Kaplan-Meier survival curves showing 20-year survival in the **a-c,** 55-64 CA bin (*n*=657) and **d-f,** 75-84 CA bin (*n*=348) in the test cohort. **a,d,** HannumAge, HorvathAge, and ChronAge showed no statistically significant differences in survival. **b,** LinAge2 demonstrated significant survival differences compared to ChronAge in both the best 25% (*P*=2.01E-03) and worst 25% (*P*=0.03) quartiles. In contrast, PhenoAge Clinical showed a significant difference only in the best 25% quartile (*P*=3.80E-02). **c,f,** In the best 25% quartile, LinAge2 outperformed DunedinPoAm (*P*=6.45E-03 and *P*=1.39E-02 in the 55-64 and 75-84 CA bins, respectively) and PhenoAge DNAm (*P*=8.90E-03 and *P*=1.73E-02 in the 55-64 and 75-84 CA bins, respectively). LinAge2 and GrimAge2 performed similarly with no significant differences between them. In the worst 25% quartile, no significant differences in survival were found between LinAge2, DunedinPoAm, PhenoAge DNAm, and GrimAge2. **e,** Compared to ChronAge, both LinAge2 (*P*=4.56E-03) and PhenoAge Clinical (*P*=3.16E-02) showed significant survival differences in the best 25% quartile, but not in the worst 25% quartile. Clocks were compared using log-rank tests with Benjamini-Hochberg correction. Areas shaded indicate 95% error bands for lines of the same color. yo, years old.

**Extended Data Fig. 2:**
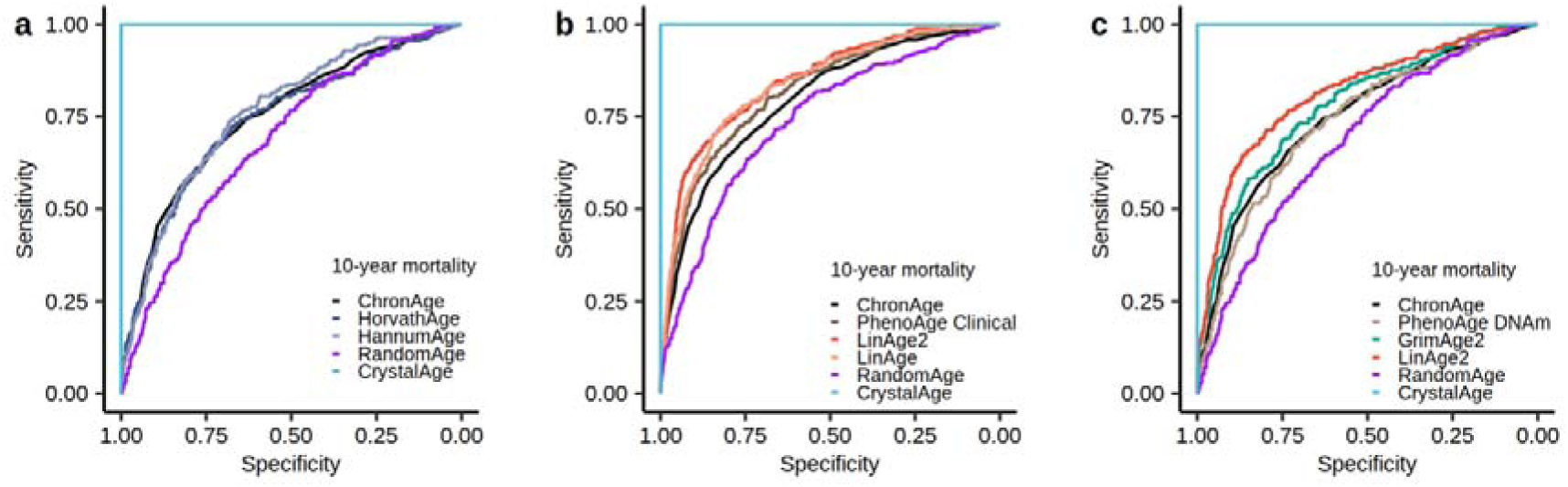
ROC curves for 10-year all-cause mortality in the test cohort. **a,** There were no significant differences in the AUCs between HorvathAge (AUC=0.7425), HannumAge (AUC=0.7612), and ChronAge (AUC=0.7501) (*n*=1,065). **b,** LinAge2 (AUC=0.8468) was significantly more informative than PhenoAge Clinical (AUC=0.8203, *P*=6.91E-05) and ChronAge (AUC=0.7946, *P*=2.65E-09) in predicting future mortality (*n*=2,036). LinAge2 performed similarly to LinAge (AUC=0.8383). **c,** Compared to LinAge2 (AUC=0.8144), PhenoAge DNAm (AUC=0.7390, *P*=2.84E-06) and GrimAge2 (AUC=0.7801, *P*=1.81E-03) were significantly less predictive of 10-year follow-up (*n*=1,065). Although GrimAge2 outperformed ChronAge (AUC=0.7501, *P*=0.01) in predicting 10-year mortality, PhenoAge DNAm did not (*P*=0.39). ROC curves were compared using DeLong’s test.

**Extended Data Fig. 3:**
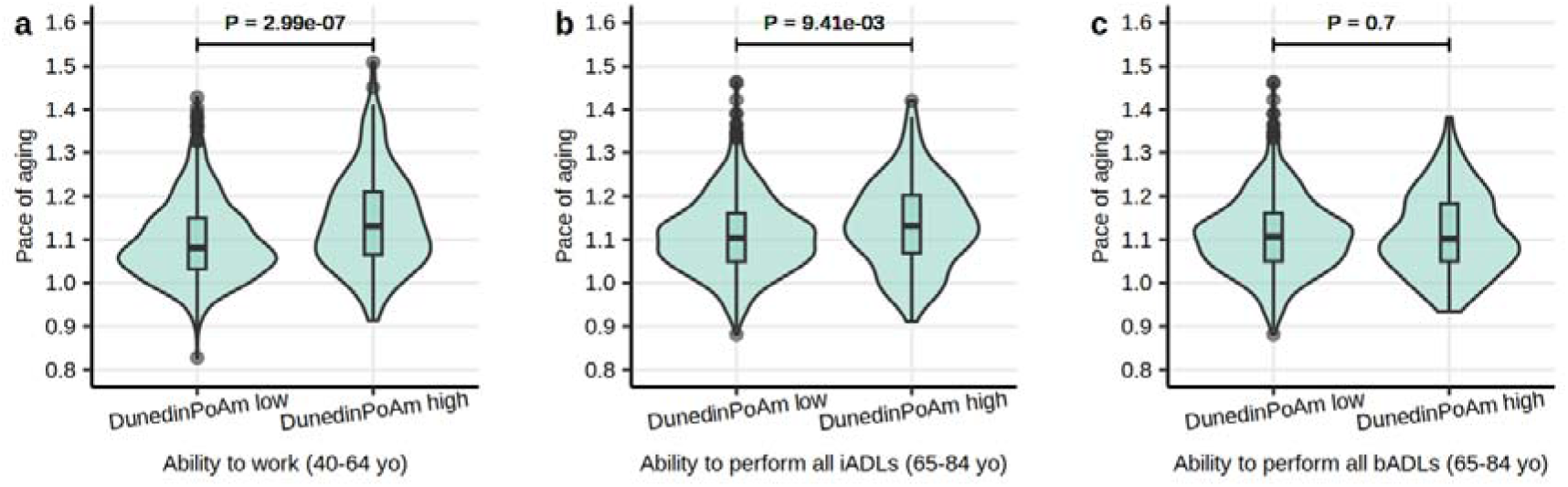
DunedinPoAm tracks with healthspan markers. Differences between DunedinPoAm low (slow aging) and DunedinPoAm high (fast aging) were significant for: **a,** ability to work; **b,** ability to perform instrumental activities of daily living (iADLs); but not for **c,** ability to perform basic activities of daily living (bADLs). Median value, lower (25^th^) and upper (75^th^) percentiles are indicated. Lines extend to ±1.5 times interquartile range, with points outside this range drawn individually. The violin shape indicates the probability density function.

**Extended Data Fig. 4:**
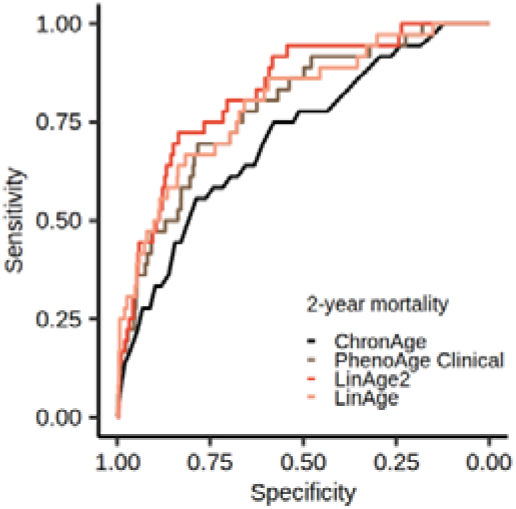
Clinical clocks are better predictors of 2-year mortality than CA. ROC curves for 2-year all-cause mortality in the test cohort. LinAge2 (AUC=0.8294, *P*=4.02E-06), LinAge (AUC=0.8003, *P*=3.91E-04) and PhenoAge Clinical (AUC=0.7870, *P*=3.24E-03) were significantly more predictive of 2-year follow-up than ChronAge (AUC=0.7120) (*n*=2,036). ROC curves were compared using DeLong’s test.

