## Supplementary material for "LinAge2: Providing actionable insights and benchmarking with epigenetic clocks": LinAge2_Supplementary_Information

| **Supplementary Table 1. Baseline characteristics of participants.** | | |
| --- | --- | --- |
|  | **NHANES 1999-2000**  **(training cohort)** | **NHANES 2001-2002**  **(testing cohort)** |
|  | ***n* = 2,079** | ***n* = 2,344** |
| Age (years) (mean ± SD) | 59.85 ± 12.38 | 58.83 ± 12.56 |
| Male sex (%) | 50.26 | 51.24 |
| Race (%) |  |  |
| - Non-Hispanic White | 47.33 | 57.59 |
| - Non-Hispanic Black | 16.64 | 17.45 |
| - Mexican American | 28.09 | 18.98 |
| - Other Hispanic | 5.48 | 3.50 |
| - Other | 2.45 | 2.47 |
| Education (%) |  |  |
| - < High school | 43.39 | 29.96 |
| - High school diploma | 20.49 | 22.92 |
| - > High school | 35.93 | 47.03 |
| - Missing | 0.19 | 0.13 |
| Poverty income ratio (mean ± SD) | 2.65 ± 1.60 | 2.97 ± 1.62 |
| - Missing (%) | 14.67 | 7.23 |
| Smoking (%) |  |  |
| - Current | 18.86 | 19.84 |
| - No | 34.01 | 33.96 |
| - Missing | 47.14 | 46.20 |
| Alcohol (%) |  |  |
| - Yes | 59.45 | 61.09 |
| - No | 23.18 | 23.42 |
| - Missing | 17.36 | 15.49 |
| Body mass index (kg/m^2^) (mean ± SD) | 28.66 ± 5.91 | 28.77 ± 6.03 |
| Mortality status at 20-year follow-up (%) |  |  |
| - Alive | 55.51 | 65.02 |
| - Deceased | 44.44 | 34.94 |
| - Missing | 0.05 | 0.04 |

| **Supplementary Table 2. Median and median absolute deviation (MAD) values utilized for normalization of clinical parameters, as well as 25th quartile (Q25), 75th quartile (Q75), and individual weights for parameters for LinAge2.** | | | | | | | | | | | |
| --- | --- | --- | --- | --- | --- | --- | --- | --- | --- | --- | --- |
| **Variable Names** | **Parameters** | **Male** | | | | | **Female** | | | | |
|  |  | **Median** | **MAD** | **Q25** | **Q75** | **Individual Weights** | **Median** | **MAD** | **Q25** | **Q75** | **Individual Weights** |
| RIDAGEEX | Chronological Age ($\beta_{CA}$) (months) | N.A.^#^ | N.A.^#^ | N.A.^#^ | N.A.^#^ | -0.0156 | N.A.^#^ | N.A.^#^ | N.A.^#^ | N.A.^#^ | -0.0092 |
| BMXBMI | Log Body Mass Index (kg/m^2^) | 3.32 | 0.17 | 3.19 | 3.43 | -0.6649 | 3.35 | 0.25 | 3.20 | 3.54 | -0.9209 |
| BPXSAR | Systolic Blood Pressure (mmHg) | 122.00 | 13.34 | 114.00 | 131.00 | 0.6827 | 118.00 | 14.83 | 108.00 | 130.00 | 0.3529 |
| BPXDAR | Diastolic Blood Pressure (mmHg) | 78.00 | 10.38 | 72.00 | 86.00 | -0.6462 | 75.00 | 10.38 | 68.00 | 82.00 | -0.0401 |
| BPXPLS | Pulse Rate (bpm) | 70.00 | 11.86 | 60.00 | 78.00 | 0.6146 | 72.00 | 11.86 | 66.00 | 80.00 | 0.7537 |
| LBXHGB | Hemoglobin (g/dL) | 15.30 | 0.89 | 14.70 | 16.00 | -0.0711 | 13.50 | 1.04 | 12.80 | 14.22 | -0.2515 |
| LBXRBCSI | Red Blood Cell Count (million cells/µL) | 5.05 | 0.37 | 4.80 | 5.28 | -0.2983 | 4.43 | 0.39 | 4.15 | 4.67 | -0.4521 |
| LBXHCT | Hematocrit (%) | 45.50 | 2.52 | 43.80 | 47.40 | 0.0438 | 39.70 | 3.11 | 37.68 | 41.90 | 0.1851 |
| LBXMCVSI | Mean Cell Volume (fL) | 90.30 | 4.45 | 87.70 | 93.60 | 0.5573 | 90.10 | 5.04 | 86.27 | 93.00 | 0.9786 |
| LBXMCHSI | Mean Cell Hemoglobin (pg) | 30.60 | 1.78 | 29.50 | 31.80 | 0.2522 | 30.60 | 2.08 | 29.00 | 31.80 | 0.3079 |
| LBXMC | Mean Cell Hemoglobin Concentration (g/dL) | 33.80 | 0.74 | 33.30 | 34.20 | -0.5135 | 33.85 | 0.82 | 33.40 | 34.40 | -1.3520 |
| LBXRDW | Red Cell Distribution Width (%) | 12.50 | 0.59 | 12.10 | 12.90 | 1.2308 | 12.45 | 0.82 | 12.00 | 13.10 | 1.0851 |
| LBXPLTSI | Platelet Count (1000 cells/µL) | 243.00 | 48.93 | 213.00 | 279.00 | -0.2833 | 277.50 | 68.20 | 236.25 | 326.25 | -0.2930 |
| LBXMPSI | Mean Platelet Volume (fL) | 8.20 | 0.74 | 7.80 | 8.80 | 0.1654 | 8.25 | 0.82 | 7.80 | 8.90 | -1.2415 |
| LBXWBCSI | Log White Blood Cell Count (1000 cells/µL) | 1.89 | 0.30 | 1.70 | 2.09 | 0.2960 | 1.94 | 0.29 | 1.74 | 2.11 | 0.1576 |
| LBXNEPCT | Segmented Neutrophils Percent (%) | 57.40 | 8.90 | 51.00 | 63.30 | 0.2223 | 57.50 | 9.56 | 51.60 | 64.32 | -0.0336 |
| LBXLYPCT | Lymphocyte Percent (%) | 30.60 | 8.01 | 25.30 | 36.30 | -0.1679 | 31.55 | 8.30 | 25.48 | 36.23 | 0.1625 |
| LBXMOPCT | Monocyte Percent (%) | 8.30 | 2.08 | 6.90 | 9.80 | -0.2468 | 7.20 | 1.78 | 6.20 | 8.72 | -0.0846 |
| LBXEOPCT | Eosinophils Percent (%) | 2.60 | 1.48 | 1.70 | 3.80 | -0.0994 | 2.20 | 1.33 | 1.50 | 3.20 | 0.0620 |
| LBXBAPCT | Basophils Percent (%) | 0.60 | 0.30 | 0.40 | 0.90 | 0.8416 | 0.70 | 0.30 | 0.48 | 0.90 | -1.1977 |
| LBDNENO | Segmented Neutrophils Number (1000 cells/µL) | 3.70 | 1.33 | 3.00 | 4.80 | 0.7111 | 4.10 | 1.63 | 3.00 | 5.10 | 0.1667 |
| LBDLYMNO | Lymphocyte Number (1000 cells/µL) | 2.10 | 0.59 | 1.70 | 2.50 | 0.0369 | 2.10 | 0.74 | 1.70 | 2.60 | 0.4609 |
| LBDMONO | Monocyte Number (1000 cells/µL) | 0.60 | 0.15 | 0.40 | 0.70 | -0.2367 | 0.50 | 0.15 | 0.40 | 0.60 | 0.0060 |
| LBDEONO | Eosinophils Number (1000 cells/µL) | 0.20 | 0.15 | 0.10 | 0.30 | -0.1177 | 0.10 | 0.15 | 0.10 | 0.20 | 0.4001 |
| LBDBANO | Basophils Number (1000 cells/µL) | N.A.^¶^ | N.A.^¶^ | N.A.^¶^ | N.A.^¶^ | 0.0564 | N.A.^¶^ | N.A.^¶^ | N.A.^¶^ | N.A.^¶^ | -0.0087 |
| LBXCRP | Log C-Reactive Protein (mg/dL) | -1.90 | 1.13 | -2.66 | -1.02 | -0.0169 | -1.27 | 1.29 | -2.12 | -0.31 | 0.3697 |
| LBXSLDSI | Log Lactate Dehydrogenase (U/L) | 5.02 | 0.17 | 4.91 | 5.14 | -0.5003 | 4.97 | 0.19 | 4.86 | 5.11 | 1.2617 |
| LBDIRNSI | Iron (µmol/L) | 17.01 | 6.11 | 13.25 | 21.84 | 0.1572 | 13.78 | 5.83 | 9.85 | 17.72 | 0.1110 |
| LBDTIBSI | Total Iron Binding Capacity (µmol/L) | 63.19 | 10.08 | 56.56 | 69.99 | 0.8878 | 66.86 | 11.14 | 59.92 | 75.59 | -0.2346 |
| LBXPCT | Transferrin Saturation (%) | 27.90 | 9.79 | 21.40 | 34.70 | -0.3471 | 21.70 | 9.34 | 15.28 | 27.68 | 0.1989 |
| LBDFERSI | Log Ferritin (µg/L) | 5.01 | 0.77 | 4.53 | 5.59 | -0.1916 | 3.69 | 1.14 | 2.93 | 4.46 | 1.2303 |
| LBDFOLSI | Log Folate (nmol/L) | 3.30 | 0.46 | 3.01 | 3.61 | 0.4887 | 3.34 | 0.47 | 3.03 | 3.66 | 0.0644 |
| LBDB12SI | Log Vitamin B12 (pmol/L) | 5.93 | 0.38 | 5.63 | 6.14 | -0.1330 | 5.83 | 0.43 | 5.54 | 6.12 | -0.2926 |
| LBDSBUSI | Blood Urea Nitrogen (mmol/L) | 5.00 | 1.63 | 3.90 | 6.10 | 0.0037 | 4.30 | 1.04 | 3.60 | 5.00 | 1.0587 |
| LBXSNASI | Sodium (mmol/L) | 139.50 | 2.37 | 138.00 | 141.10 | -0.9851 | 139.30 | 2.52 | 137.48 | 141.00 | 0.1037 |
| LBXSKSI | Potassium (mmol/L) | 4.17 | 0.30 | 3.96 | 4.35 | -0.4695 | 3.96 | 0.29 | 3.81 | 4.20 | -0.1022 |
| LBXSCLSI | Chloride (mmol/L) | 102.10 | 2.97 | 100.30 | 104.30 | -0.9614 | 102.80 | 2.67 | 100.90 | 104.40 | -0.3935 |
| LBXSC3SI | Bicarbonate (mmol/L) | 24.00 | 1.48 | 22.00 | 25.00 | 0.1890 | 23.00 | 1.48 | 22.00 | 25.00 | 1.0227 |
| LBDSCRSI | Creatinine (µmol/L) | 70.70 | 13.05 | 61.90 | 79.60 | 0.0899 | 53.00 | 13.05 | 44.20 | 61.90 | -0.6691 |
| LBDSCASI | Calcium Total (mmol/L) | 2.35 | 0.07 | 2.30 | 2.40 | 1.1378 | 2.33 | 0.11 | 2.25 | 2.38 | -0.6769 |
| LBDSPHSI | Phosphorus (mmol/L) | 1.10 | 0.14 | 1.00 | 1.16 | 0.0550 | 1.10 | 0.19 | 1.00 | 1.23 | 0.5391 |
| LBDSTPSI | Protein Total (g/L) | 76.00 | 4.45 | 73.00 | 79.00 | -0.3174 | 75.00 | 4.45 | 72.00 | 78.00 | -0.4500 |
| LBDSALSI | Albumin (g/L) | 46.00 | 2.97 | 44.00 | 47.00 | -1.8869 | 44.00 | 2.97 | 42.00 | 45.00 | -0.4568 |
| LBDSGBSI | Globulin (g/L) | 30.00 | 4.45 | 28.00 | 33.00 | 0.9143 | 31.00 | 4.45 | 28.75 | 34.00 | -0.1538 |
| LBDSTBSI | Bilirubin (µmol/L) | 10.30 | 2.52 | 8.60 | 13.70 | -0.3771 | 6.80 | 2.67 | 5.10 | 10.30 | -0.6931 |
| LBXSAPSI | Log Alkaline Phosphatase (IU/L) | 4.37 | 0.29 | 4.19 | 4.57 | -0.3960 | 4.32 | 0.32 | 4.06 | 4.52 | 1.6302 |
| LBXSATSI | Log Alanine Aminotransferase (U/L) | 3.37 | 0.48 | 3.09 | 3.71 | -0.4560 | 2.89 | 0.37 | 2.71 | 3.26 | -0.1015 |
| LBXSASSI | Log Aspartate Aminotransferase (U/L) | 3.22 | 0.27 | 3.04 | 3.47 | 0.1985 | 3.00 | 0.24 | 2.83 | 3.18 | 0.6217 |
| LBDSUASI | Uric Acid (µmol/L) | 339.00 | 70.57 | 291.50 | 392.60 | 0.9963 | 255.80 | 61.82 | 214.10 | 303.30 | 0.7039 |
| LBDSGLSI | Glucose (mmol/L) | 5.05 | 0.58 | 4.72 | 5.44 | -0.3060 | 4.88 | 0.49 | 4.55 | 5.22 | 0.7204 |
| LBXGH | Glycohemoglobin (%) | 5.30 | 0.44 | 5.10 | 5.60 | 0.6176 | 5.20 | 0.44 | 5.00 | 5.50 | 0.4117 |
| LDLV | Low-Density Lipoprotein (mmol/L) | 3.84 | 1.05 | 3.09 | 4.52 | 0.1376 | 3.55 | 0.96 | 2.93 | 4.20 | 0.5774 |
| SSBNP | Log N-Terminal Pro-Brain Natriuretic Peptide (pg/mL) | 3.13 | 0.91 | 2.53 | 3.75 | 1.9505 | 3.96 | 0.79 | 3.35 | 4.41 | 1.9210 |
| URXUMASI | Urine Albumin (mg/L) | 7.40 | 5.78 | 4.10 | 12.20 | 0.6967 | 7.20 | 7.49 | 3.20 | 15.03 | 0.0769 |
| URXUCRSI | Log Urine Creatinine (µmol/L) | 9.44 | 0.49 | 8.99 | 9.74 | 0.0999 | 9.17 | 0.76 | 8.48 | 9.59 | -0.1815 |
| crAlbRat | Log Urine Albumin-to-Creatinine Ratio (mg/g) | 1.58 | 0.59 | 1.23 | 2.05 | 0.1664 | 1.85 | 0.70 | 1.44 | 2.40 | 0.2231 |
| LBXCOT | Smoking status / Cotinine (ng/mL) | N.A.* | N.A.* | N.A.* | N.A.* | 2.9341 | N.A.* | N.A.* | N.A.* | N.A.* | 3.0916 |
| fs1Score | Co-morbidity index | N.A.^#^ | N.A.^#^ | N.A.^#^ | N.A.^#^ | 0.0587 | N.A.^#^ | N.A.^#^ | N.A.^#^ | N.A.^#^ | 0.0794 |
| fs2Score | Self-health index | N.A.^#^ | N.A.^#^ | N.A.^#^ | N.A.^#^ | 0.8806 | N.A.^#^ | N.A.^#^ | N.A.^#^ | N.A.^#^ | 1.0748 |
| fs3Score | Healthcare use index | N.A.^#^ | N.A.^#^ | N.A.^#^ | N.A.^#^ | -0.1169 | N.A.^#^ | N.A.^#^ | N.A.^#^ | N.A.^#^ | 0.6267 |
| N.A.^##^ | $C_{0}$ Constant | N.A.^#^ | N.A.^#^ | N.A.^#^ | N.A.^#^ | 2.17 | N.A.^#^ | N.A.^#^ | N.A.^#^ | N.A.^#^ | -3.84 |
| N.A. = not applicable  ^¶^ N.A. because the median and MAD were 0, hence, actual Basophils Number were used instead  * N.A. because smoking status was determined by using actual serum cotinine levels organized into bins – 0-10 ng/mL (non-smokers), 10-99 ng/mL (light smokers), 100-199 (moderate smokers), and ≥ 200 (heavy smokers) – which could be replaced by questionnaire data if cotinine data are not available  ^#^ N.A. because actual scores were used  ^##^ N.A. because the constant does not have a variable name | | | | | | | | | | | |

| **Supplementary Table 3. PC loadings in LinAge2.** | | | |
| --- | --- | --- | --- |
| **PC (Male)** | **PC Loadings** | **Variable Names** | **Parameters** |
| PC1M | 0.463 0.401 0.386 0.292 0.271 0.266 0.188 0.18 0.152 0.128 0.125 0.102 0.089 0.083 0.063 0.061 0.059 0.053 0.052 0.049 0.042 0.033 0.031 0.028 0.028 0.027 0.025 0.025 0.023 0.02 0.019 0.018 0.011 0.009 0.006 0.004 0.002 -0.11 -0.104 -0.1 -0.099 -0.085 -0.069 -0.069 -0.058 -0.046 -0.031 -0.022 -0.022 -0.022 -0.021 -0.02 -0.018 -0.017 -0.014 -0.01 -0.01 -0.007 -0.005 | URXUMASI crAlbRat (logged) fs3Score LBXGH LBDSGLSI SSBNP (logged) fs2Score BPXSAR LBXRDW LBDSCRSI LBDSBUSI LBXCRP (logged) LBDNENO LBXCOT LBDSGBSI LBDSUASI LBXEOPCT LBDFOLSI (logged) LBXNEPCT LBXMPSI LBXSLDSI (logged) LBXWBCSI (logged) LBXMOPCT BMXBMI (logged) LBXMCVSI LBDEONO LBXSAPSI (logged) LBXSKSI LBXBAPCT fs1Score LBDMONO BPXPLS LBXMCHSI LBXSC3SI LBDBANO LBDSTBSI LBDSCASI LBXHCT LBDSALSI LBXHGB LBXRBCSI BPXDAR URXUCRSI (logged) LBXLYPCT LBXSATSI (logged) LBDLYMNO LBDSPHSI LBXPCT LBXMC LBXSCLSI LBDIRNSI LBDB12SI (logged) LDLV LBXSASSI (logged) LBDFERSI (logged) LBDTIBSI LBXSNASI LBXPLTSI LBDSTPSI | Albumin urine (mg/L) Log Urine Albumin-to-Creatinine Ratio (mg/g) (logged) Healthcare use index Glycohemoglobin (%) Glucose (mmol/L) NT-proBNP (pg/ml) (logged) Self-health index SBP average reported to examinee Red cell distribution width (percent) Creatinine (umol/L) Blood Urea Nitrogen (mmol/L) CRP (mg/dL) (logged) Segmented neutrophils number (1000 cell/uL) Cotinine (ng/mL) Globulin (g/L) Uric acid (umol/L) Eosinophils percent Folate serum (nmol/L) (logged) Segmented neutrophils percent Mean platelet volume (fL) Lactate Dehydrogenase (LDH) (U/L) (logged) WBC count (1000 cells/uL) (logged) Monocyte percent Body Mass Index (kg/m2) (logged) Mean cell volume (fL) Eosinophils number (1000 cells/uL) Alkaline Phosphatase (ALP) (IU/L) (logged) Potassium (mmol/L) Basophils percent Co-morbidity index Monocyte number (1000 cells/uL) 60 sec pulse (30 sec pulse X2) Mean cell hemoglobin (pg) Bicarbonate (mmol/L) Basophils number (1000 cells/uL) Bilirubin total (umol/L) Calcium total (mmol/L) Hematocrit Albumin (g/L) Hemoglobin (g/dL) Red blood cell count (million cells/uL) DBP average reported to examinee Creatinine urine (umol/L) (logged) Lymphocyte percent Alanine Aminotransferase (ALT) (U/L) (logged) Lymphocyte number (1000 cells/uL) Phosphorus (mmol/L) Transferrin Saturation (%) Mean Cell Hemoglobin Concentration (g/dL) Chloride (mmol/L) Iron (umol/L) Vitamin B12 serum (pmol/L) (logged) Low-Density Lipoprotein (mmol/L) Aspartate Aminotransferase (AST) (U/L) (logged) Ferritin (ug/L) (logged) Total iron binding capacity (umol/L) Sodium (mmol/L) Platelet count (1000 cells/uL) Protein total (g/L) |
| PC2M | 0.195 0.177 0.143 0.115 0.113 0.08 0.078 0.061 0.053 0.048 0.045 0.036 0.019 0.016 0.015 0.014 0.014 0.006 0.006 0.001 -0.402 -0.397 -0.312 -0.308 -0.215 -0.188 -0.164 -0.162 -0.159 -0.157 -0.122 -0.12 -0.117 -0.115 -0.108 -0.108 -0.102 -0.099 -0.096 -0.095 -0.093 -0.089 -0.077 -0.063 -0.055 -0.054 -0.052 -0.047 -0.041 -0.041 -0.035 -0.03 -0.023 -0.02 -0.019 -0.012 -0.005 -0.004 -0.002 | SSBNP (logged) fs3Score LBXRDW LBDSCRSI LBDSBUSI LBXEOPCT LBXMOPCT LBXSCLSI LBXSC3SI LBDFOLSI (logged) LBXSKSI LBXBAPCT fs2Score LBXMCVSI LBXSNASI LBDEONO LBDSPHSI LBXPLTSI fs1Score LBDBANO LBXHCT LBXHGB URXUMASI LBXRBCSI crAlbRat (logged) LBXGH BPXDAR LBXSATSI (logged) LBDSGLSI LBDFERSI (logged) LBDLYMNO LBDSTPSI LBDIRNSI URXUCRSI (logged) BMXBMI (logged) LBXSASSI (logged) LBDSTBSI LBDSALSI LBDSCASI BPXPLS LBXWBCSI (logged) LBXPCT LBDNENO LDLV LBXMPSI LBDSGBSI LBDMONO LBDSUASI LBDTIBSI LBXMC LBXSAPSI (logged) LBXLYPCT BPXSAR LBDB12SI (logged) LBXSLDSI (logged) LBXCOT LBXCRP (logged) LBXNEPCT LBXMCHSI | NT-proBNP (pg/ml) (logged) Healthcare use index Red cell distribution width (percent) Creatinine (umol/L) Blood Urea Nitrogen (mmol/L) Eosinophils percent Monocyte percent Chloride (mmol/L) Bicarbonate (mmol/L) Folate serum (nmol/L) (logged) Potassium (mmol/L) Basophils percent Self-health index Mean cell volume (fL) Sodium (mmol/L) Eosinophils number (1000 cells/uL) Phosphorus (mmol/L) Platelet count (1000 cells/uL) Co-morbidity index Basophils number (1000 cells/uL) Hematocrit Hemoglobin (g/dL) Albumin urine (mg/L) Red blood cell count (million cells/uL) Log Urine Albumin-to-Creatinine Ratio (mg/g) (logged) Glycohemoglobin (%) DBP average reported to examinee Alanine Aminotransferase (ALT) (U/L) (logged) Glucose (mmol/L) Ferritin (ug/L) (logged) Lymphocyte number (1000 cells/uL) Protein total (g/L) Iron (umol/L) Creatinine urine (umol/L) (logged) Body Mass Index (kg/m2) (logged) Aspartate Aminotransferase (AST) (U/L) (logged) Bilirubin total (umol/L) Albumin (g/L) Calcium total (mmol/L) 60 sec pulse (30 sec pulse X2) WBC count (1000 cells/uL) (logged) Transferrin Saturation (%) Segmented neutrophils number (1000 cell/uL) Low-Density Lipoprotein (mmol/L) Mean platelet volume (fL) Globulin (g/L) Monocyte number (1000 cells/uL) Uric acid (umol/L) Total iron binding capacity (umol/L) Mean Cell Hemoglobin Concentration (g/dL) Alkaline Phosphatase (ALP) (IU/L) (logged) Lymphocyte percent SBP average reported to examinee Vitamin B12 serum (pmol/L) (logged) Lactate Dehydrogenase (LDH) (U/L) (logged) Cotinine (ng/mL) CRP (mg/dL) (logged) Segmented neutrophils percent Mean cell hemoglobin (pg) |
| PC5M | 0.407 0.302 0.253 0.242 0.241 0.21 0.188 0.17 0.157 0.155 0.145 0.129 0.119 0.106 0.093 0.091 0.085 0.082 0.074 0.074 0.065 0.057 0.05 0.044 0.042 0.04 0.036 0.031 0.022 0.022 0.015 0.009 0.008 0.006 0.006 0.005 -0.273 -0.231 -0.204 -0.142 -0.127 -0.122 -0.116 -0.101 -0.094 -0.074 -0.059 -0.057 -0.057 -0.055 -0.048 -0.04 -0.039 -0.039 -0.032 -0.03 -0.025 -0.023 -0.019 | LBDNENO LBXWBCSI (logged) LBDMONO LBXPLTSI LBXNEPCT LBXCOT LBXHGB LBXHCT fs2Score LBXMCVSI LBXMCHSI SSBNP (logged) LBXCRP (logged) fs3Score LBXSKSI LBDSCASI BPXPLS LBDEONO LBDLYMNO LBXSAPSI (logged) BPXSAR LBDTIBSI LBXMC LBXRDW LBDFOLSI (logged) LBDSPHSI LBDSUASI LBDSALSI LBXRBCSI LBDSTPSI LBDSBUSI LDLV LBDBANO LBDSCRSI fs1Score LBDSGBSI LBDSGLSI LBXLYPCT LBXGH URXUCRSI (logged) URXUMASI LBXMPSI LBDSTBSI LBXSC3SI LBXMOPCT LBXPCT LBDFERSI (logged) BMXBMI (logged) LBXSATSI (logged) LBXSASSI (logged) LBXEOPCT BPXDAR LBDIRNSI crAlbRat (logged) LBDB12SI (logged) LBXBAPCT LBXSCLSI LBXSLDSI (logged) LBXSNASI | Segmented neutrophils number (1000 cell/uL) WBC count (1000 cells/uL) (logged) Monocyte number (1000 cells/uL) Platelet count (1000 cells/uL) Segmented neutrophils percent Cotinine (ng/mL) Hemoglobin (g/dL) Hematocrit Self-health index Mean cell volume (fL) Mean cell hemoglobin (pg) NT-proBNP (pg/ml) (logged) CRP (mg/dL) (logged) Healthcare use index Potassium (mmol/L) Calcium total (mmol/L) 60 sec pulse (30 sec pulse X2) Eosinophils number (1000 cells/uL) Lymphocyte number (1000 cells/uL) Alkaline Phosphatase (ALP) (IU/L) (logged) SBP average reported to examinee Total iron binding capacity (umol/L) Mean Cell Hemoglobin Concentration (g/dL) Red cell distribution width (percent) Folate serum (nmol/L) (logged) Phosphorus (mmol/L) Uric acid (umol/L) Albumin (g/L) Red blood cell count (million cells/uL) Protein total (g/L) Blood Urea Nitrogen (mmol/L) Low-Density Lipoprotein (mmol/L) Basophils number (1000 cells/uL) Creatinine (umol/L) Co-morbidity index Globulin (g/L) Glucose (mmol/L) Lymphocyte percent Glycohemoglobin (%) Creatinine urine (umol/L) (logged) Albumin urine (mg/L) Mean platelet volume (fL) Bilirubin total (umol/L) Bicarbonate (mmol/L) Monocyte percent Transferrin Saturation (%) Ferritin (ug/L) (logged) Body Mass Index (kg/m2) (logged) Alanine Aminotransferase (ALT) (U/L) (logged) Aspartate Aminotransferase (AST) (U/L) (logged) Eosinophils percent DBP average reported to examinee Iron (umol/L) Log Urine Albumin-to-Creatinine Ratio (mg/g) (logged) Vitamin B12 serum (pmol/L) (logged) Basophils percent Chloride (mmol/L) Lactate Dehydrogenase (LDH) (U/L) (logged) Sodium (mmol/L) |
| PC6M | 0.349 0.269 0.264 0.254 0.219 0.207 0.197 0.181 0.179 0.176 0.149 0.134 0.127 0.121 0.119 0.115 0.113 0.092 0.088 0.069 0.055 0.045 0.045 0.044 0.037 0.035 0.029 0.02 0.018 0.016 0.016 0.014 0.013 0.008 0.007 0.006 0.005 -0.349 -0.178 -0.155 -0.15 -0.145 -0.143 -0.123 -0.101 -0.099 -0.082 -0.069 -0.061 -0.056 -0.04 -0.038 -0.033 -0.026 -0.022 -0.021 -0.007 -0.007 -0.002 | LBXEOPCT LBDLYMNO LBXLYPCT LBDEONO LBXSASSI (logged) LBDSGBSI LBDSTPSI LBXMOPCT LBDMONO LBXBAPCT LBDSCASI fs3Score LBXRDW LBXCOT fs2Score LBXSATSI (logged) LBDSPHSI BPXDAR LBXSAPSI (logged) LBXPLTSI LBXSLDSI (logged) BPXSAR LBDTIBSI LBXMPSI LBDB12SI (logged) LBDIRNSI BPXPLS LBDFERSI (logged) LBXPCT LBXCRP (logged) LBDSUASI crAlbRat (logged) LBXGH LBDBANO LBXWBCSI (logged) LBXSKSI fs1Score LBXNEPCT LBDSGLSI LBDNENO URXUCRSI (logged) LBXSC3SI LBXSNASI LBDSBUSI LBXMC LBXSCLSI URXUMASI LBDSTBSI LBDSCRSI LBXHGB LBXMCHSI SSBNP (logged) LBDFOLSI (logged) LBDSALSI LBXHCT BMXBMI (logged) LBXRBCSI LDLV LBXMCVSI | Eosinophils percent Lymphocyte number (1000 cells/uL) Lymphocyte percent Eosinophils number (1000 cells/uL) Aspartate Aminotransferase (AST) (U/L) (logged) Globulin (g/L) Protein total (g/L) Monocyte percent Monocyte number (1000 cells/uL) Basophils percent Calcium total (mmol/L) Healthcare use index Red cell distribution width (percent) Cotinine (ng/mL) Self-health index Alanine Aminotransferase (ALT) (U/L) (logged) Phosphorus (mmol/L) DBP average reported to examinee Alkaline Phosphatase (ALP) (IU/L) (logged) Platelet count (1000 cells/uL) Lactate Dehydrogenase (LDH) (U/L) (logged) SBP average reported to examinee Total iron binding capacity (umol/L) Mean platelet volume (fL) Vitamin B12 serum (pmol/L) (logged) Iron (umol/L) 60 sec pulse (30 sec pulse X2) Ferritin (ug/L) (logged) Transferrin Saturation (%) CRP (mg/dL) (logged) Uric acid (umol/L) Log Urine Albumin-to-Creatinine Ratio (mg/g) (logged) Glycohemoglobin (%) Basophils number (1000 cells/uL) WBC count (1000 cells/uL) (logged) Potassium (mmol/L) Co-morbidity index Segmented neutrophils percent Glucose (mmol/L) Segmented neutrophils number (1000 cell/uL) Creatinine urine (umol/L) (logged) Bicarbonate (mmol/L) Sodium (mmol/L) Blood Urea Nitrogen (mmol/L) Mean Cell Hemoglobin Concentration (g/dL) Chloride (mmol/L) Albumin urine (mg/L) Bilirubin total (umol/L) Creatinine (umol/L) Hemoglobin (g/dL) Mean cell hemoglobin (pg) NT-proBNP (pg/ml) (logged) Folate serum (nmol/L) (logged) Albumin (g/L) Hematocrit Body Mass Index (kg/m2) (logged) Red blood cell count (million cells/uL) Low-Density Lipoprotein (mmol/L) Mean cell volume (fL) |
| PC8M | 0.346 0.26 0.226 0.225 0.199 0.189 0.179 0.153 0.153 0.126 0.122 0.114 0.106 0.091 0.091 0.082 0.078 0.06 0.056 0.047 0.039 0.036 0.031 0.031 0.02 0.004 0.002 0.002 -0.32 -0.261 -0.2 -0.199 -0.197 -0.18 -0.155 -0.131 -0.12 -0.109 -0.1 -0.091 -0.091 -0.088 -0.077 -0.077 -0.075 -0.067 -0.061 -0.047 -0.04 -0.038 -0.038 -0.029 -0.019 -0.018 -0.018 -0.013 -0.011 -0.005 -0.001 | LBXSASSI (logged) LBXSLDSI (logged) LBXSATSI (logged) LBDSTBSI LBDSGBSI LBDSTPSI LBXNEPCT LBXRDW LBDSUASI LBDB12SI (logged) BPXDAR LBDSCASI LBXSAPSI (logged) BPXPLS fs2Score LBXCRP (logged) LBDTIBSI BPXSAR fs3Score LBDSCRSI LBDSPHSI LBDFERSI (logged) LBDFOLSI (logged) LBDNENO BMXBMI (logged) LBDSBUSI SSBNP (logged) fs1Score LBXEOPCT LBDEONO LBXSNASI LBXSC3SI LBDLYMNO LBXSCLSI URXUMASI LBXLYPCT LBDMONO LBXHGB LBXHCT LBXMCVSI crAlbRat (logged) LBXMCHSI LBXWBCSI (logged) LBXSKSI LBXPCT LBXBAPCT LBXCOT LBDIRNSI URXUCRSI (logged) LBXMC LBXPLTSI LDLV LBDSALSI LBXMOPCT LBXGH LBXRBCSI LBDSGLSI LBDBANO LBXMPSI | Aspartate Aminotransferase (AST) (U/L) (logged) Lactate Dehydrogenase (LDH) (U/L) (logged) Alanine Aminotransferase (ALT) (U/L) (logged) Bilirubin total (umol/L) Globulin (g/L) Protein total (g/L) Segmented neutrophils percent Red cell distribution width (percent) Uric acid (umol/L) Vitamin B12 serum (pmol/L) (logged) DBP average reported to examinee Calcium total (mmol/L) Alkaline Phosphatase (ALP) (IU/L) (logged) 60 sec pulse (30 sec pulse X2) Self-health index CRP (mg/dL) (logged) Total iron binding capacity (umol/L) SBP average reported to examinee Healthcare use index Creatinine (umol/L) Phosphorus (mmol/L) Ferritin (ug/L) (logged) Folate serum (nmol/L) (logged) Segmented neutrophils number (1000 cell/uL) Body Mass Index (kg/m2) (logged) Blood Urea Nitrogen (mmol/L) NT-proBNP (pg/ml) (logged) Co-morbidity index Eosinophils percent Eosinophils number (1000 cells/uL) Sodium (mmol/L) Bicarbonate (mmol/L) Lymphocyte number (1000 cells/uL) Chloride (mmol/L) Albumin urine (mg/L) Lymphocyte percent Monocyte number (1000 cells/uL) Hemoglobin (g/dL) Hematocrit Mean cell volume (fL) Log Urine Albumin-to-Creatinine Ratio (mg/g) (logged) Mean cell hemoglobin (pg) WBC count (1000 cells/uL) (logged) Potassium (mmol/L) Transferrin Saturation (%) Basophils percent Cotinine (ng/mL) Iron (umol/L) Creatinine urine (umol/L) (logged) Mean Cell Hemoglobin Concentration (g/dL) Platelet count (1000 cells/uL) Low-Density Lipoprotein (mmol/L) Albumin (g/L) Monocyte percent Glycohemoglobin (%) Red blood cell count (million cells/uL) Glucose (mmol/L) Basophils number (1000 cells/uL) Mean platelet volume (fL) |
| PC11M | 0.267 0.207 0.205 0.19 0.186 0.181 0.122 0.103 0.101 0.091 0.089 0.086 0.079 0.06 0.054 0.037 0.034 0.026 0.018 0.009 -0.269 -0.248 -0.246 -0.243 -0.234 -0.225 -0.218 -0.181 -0.171 -0.141 -0.138 -0.136 -0.135 -0.117 -0.112 -0.108 -0.106 -0.101 -0.1 -0.085 -0.085 -0.08 -0.08 -0.078 -0.068 -0.062 -0.041 -0.039 -0.038 -0.036 -0.034 -0.031 -0.031 -0.027 -0.025 -0.014 -0.011 -0.005 -0.003 | LBDSTBSI LBXPCT LBDIRNSI LBXCOT LBXRDW LBDSCASI LBXPLTSI fs2Score URXUMASI LBXLYPCT LBDSTPSI crAlbRat (logged) LBDSGBSI LBXSKSI SSBNP (logged) LBDLYMNO LBDSPHSI LBXRBCSI LBDSALSI URXUCRSI (logged) LBXSLDSI (logged) LBXSATSI (logged) LBXSASSI (logged) LBXSNASI LBXSC3SI BMXBMI (logged) BPXDAR BPXSAR LBXEOPCT LBDSUASI LBXSCLSI LBDEONO fs3Score LBXBAPCT LBDFOLSI (logged) LBXCRP (logged) LBXMPSI LBXMC LBDSBUSI LBDB12SI (logged) LBXMCHSI LBDMONO LDLV LBDSCRSI LBXHGB LBXMCVSI LBXNEPCT LBXMOPCT LBDNENO LBXGH LBXWBCSI (logged) LBXHCT LBXSAPSI (logged) LBDFERSI (logged) BPXPLS LBDSGLSI LBDTIBSI fs1Score LBDBANO | Bilirubin total (umol/L) Transferrin Saturation (%) Iron (umol/L) Cotinine (ng/mL) Red cell distribution width (percent) Calcium total (mmol/L) Platelet count (1000 cells/uL) Self-health index Albumin urine (mg/L) Lymphocyte percent Protein total (g/L) Log Urine Albumin-to-Creatinine Ratio (mg/g) (logged) Globulin (g/L) Potassium (mmol/L) NT-proBNP (pg/ml) (logged) Lymphocyte number (1000 cells/uL) Phosphorus (mmol/L) Red blood cell count (million cells/uL) Albumin (g/L) Creatinine urine (umol/L) (logged) Lactate Dehydrogenase (LDH) (U/L) (logged) Alanine Aminotransferase (ALT) (U/L) (logged) Aspartate Aminotransferase (AST) (U/L) (logged) Sodium (mmol/L) Bicarbonate (mmol/L) Body Mass Index (kg/m2) (logged) DBP average reported to examinee SBP average reported to examinee Eosinophils percent Uric acid (umol/L) Chloride (mmol/L) Eosinophils number (1000 cells/uL) Healthcare use index Basophils percent Folate serum (nmol/L) (logged) CRP (mg/dL) (logged) Mean platelet volume (fL) Mean Cell Hemoglobin Concentration (g/dL) Blood Urea Nitrogen (mmol/L) Vitamin B12 serum (pmol/L) (logged) Mean cell hemoglobin (pg) Monocyte number (1000 cells/uL) Low-Density Lipoprotein (mmol/L) Creatinine (umol/L) Hemoglobin (g/dL) Mean cell volume (fL) Segmented neutrophils percent Monocyte percent Segmented neutrophils number (1000 cell/uL) Glycohemoglobin (%) WBC count (1000 cells/uL) (logged) Hematocrit Alkaline Phosphatase (ALP) (IU/L) (logged) Ferritin (ug/L) (logged) 60 sec pulse (30 sec pulse X2) Glucose (mmol/L) Total iron binding capacity (umol/L) Co-morbidity index Basophils number (1000 cells/uL) |
| PC15M | 0.48 0.429 0.204 0.202 0.137 0.137 0.117 0.117 0.115 0.11 0.108 0.098 0.089 0.086 0.08 0.071 0.07 0.068 0.055 0.049 0.045 0.043 0.035 0.035 0.023 0.018 0.01 0.004 0.001 -0.265 -0.218 -0.215 -0.188 -0.156 -0.149 -0.143 -0.142 -0.101 -0.09 -0.082 -0.074 -0.072 -0.067 -0.058 -0.054 -0.048 -0.043 -0.041 -0.04 -0.029 -0.025 -0.021 -0.018 -0.016 -0.007 -0.005 -0.002 -0.002 -0.001 | LBXSC3SI fs2Score LBXPCT LBDFERSI (logged) URXUCRSI (logged) LBDSCRSI LBXRDW LBXSAPSI (logged) LBDB12SI (logged) LBXSLDSI (logged) LBXMPSI LBDLYMNO LBDSGBSI LBDIRNSI LBXCRP (logged) LBDSUASI LBDSBUSI LBXLYPCT LBXCOT SSBNP (logged) LBXWBCSI (logged) LBDNENO LBDSTPSI LDLV LBXHCT LBXRBCSI LBDEONO BPXPLS LBXMCVSI LBDTIBSI LBDFOLSI (logged) LBDSTBSI LBXSCLSI LBXBAPCT LBXMOPCT fs3Score LBDSCASI LBXPLTSI LBDMONO LBDSALSI LBXGH LBXSNASI crAlbRat (logged) LBDSGLSI LBXMC LBXSKSI BPXDAR URXUMASI BMXBMI (logged) BPXSAR LBXEOPCT LBXNEPCT LBXMCHSI LBXSATSI (logged) LBDBANO LBXHGB LBXSASSI (logged) fs1Score LBDSPHSI | Bicarbonate (mmol/L) Self-health index Transferrin Saturation (%) Ferritin (ug/L) (logged) Creatinine urine (umol/L) (logged) Creatinine (umol/L) Red cell distribution width (percent) Alkaline Phosphatase (ALP) (IU/L) (logged) Vitamin B12 serum (pmol/L) (logged) Lactate Dehydrogenase (LDH) (U/L) (logged) Mean platelet volume (fL) Lymphocyte number (1000 cells/uL) Globulin (g/L) Iron (umol/L) CRP (mg/dL) (logged) Uric acid (umol/L) Blood Urea Nitrogen (mmol/L) Lymphocyte percent Cotinine (ng/mL) NT-proBNP (pg/ml) (logged) WBC count (1000 cells/uL) (logged) Segmented neutrophils number (1000 cell/uL) Protein total (g/L) Low-Density Lipoprotein (mmol/L) Hematocrit Red blood cell count (million cells/uL) Eosinophils number (1000 cells/uL) 60 sec pulse (30 sec pulse X2) Mean cell volume (fL) Total iron binding capacity (umol/L) Folate serum (nmol/L) (logged) Bilirubin total (umol/L) Chloride (mmol/L) Basophils percent Monocyte percent Healthcare use index Calcium total (mmol/L) Platelet count (1000 cells/uL) Monocyte number (1000 cells/uL) Albumin (g/L) Glycohemoglobin (%) Sodium (mmol/L) Log Urine Albumin-to-Creatinine Ratio (mg/g) (logged) Glucose (mmol/L) Mean Cell Hemoglobin Concentration (g/dL) Potassium (mmol/L) DBP average reported to examinee Albumin urine (mg/L) Body Mass Index (kg/m2) (logged) SBP average reported to examinee Eosinophils percent Segmented neutrophils percent Mean cell hemoglobin (pg) Alanine Aminotransferase (ALT) (U/L) (logged) Basophils number (1000 cells/uL) Hemoglobin (g/dL) Aspartate Aminotransferase (AST) (U/L) (logged) Co-morbidity index Phosphorus (mmol/L) |
| PC16M | 0.306 0.234 0.221 0.219 0.165 0.149 0.147 0.146 0.143 0.119 0.118 0.091 0.086 0.085 0.08 0.076 0.072 0.067 0.066 0.059 0.059 0.046 0.044 0.028 0.025 0.023 0.019 0.018 0.01 0.005 -0.432 -0.224 -0.218 -0.217 -0.194 -0.158 -0.156 -0.146 -0.132 -0.124 -0.119 -0.113 -0.102 -0.085 -0.072 -0.071 -0.068 -0.067 -0.062 -0.052 -0.046 -0.041 -0.035 -0.03 -0.026 -0.018 -0.005 -0.005 -0.003 | LBXMOPCT LBXSKSI LBXHCT LBXCOT BPXSAR LBXRDW SSBNP (logged) LBXSCLSI LBXHGB LBDMONO LBXMCVSI LBXRBCSI LBDSGBSI LBDSCRSI LBXSNASI LBDB12SI (logged) LBDFERSI (logged) LBXMPSI LBDSCASI LBXSASSI (logged) LBDSGLSI LBXPCT LBXSAPSI (logged) LBXSLDSI (logged) LBXMCHSI LBDSTPSI LBDIRNSI LBXGH LBDSBUSI LBXLYPCT LBDSTBSI LBXEOPCT fs2Score LBDEONO LBXMC LBXPLTSI LBDNENO LBXWBCSI (logged) BMXBMI (logged) LBDLYMNO LBDSPHSI LBXSC3SI LBDSALSI LBDSUASI LBDTIBSI BPXPLS BPXDAR LBDFOLSI (logged) URXUMASI LDLV LBXBAPCT crAlbRat (logged) LBXNEPCT LBXSATSI (logged) fs3Score URXUCRSI (logged) LBDBANO fs1Score LBXCRP (logged) | Monocyte percent Potassium (mmol/L) Hematocrit Cotinine (ng/mL) SBP average reported to examinee Red cell distribution width (percent) NT-proBNP (pg/ml) (logged) Chloride (mmol/L) Hemoglobin (g/dL) Monocyte number (1000 cells/uL) Mean cell volume (fL) Red blood cell count (million cells/uL) Globulin (g/L) Creatinine (umol/L) Sodium (mmol/L) Vitamin B12 serum (pmol/L) (logged) Ferritin (ug/L) (logged) Mean platelet volume (fL) Calcium total (mmol/L) Aspartate Aminotransferase (AST) (U/L) (logged) Glucose (mmol/L) Transferrin Saturation (%) Alkaline Phosphatase (ALP) (IU/L) (logged) Lactate Dehydrogenase (LDH) (U/L) (logged) Mean cell hemoglobin (pg) Protein total (g/L) Iron (umol/L) Glycohemoglobin (%) Blood Urea Nitrogen (mmol/L) Lymphocyte percent Bilirubin total (umol/L) Eosinophils percent Self-health index Eosinophils number (1000 cells/uL) Mean Cell Hemoglobin Concentration (g/dL) Platelet count (1000 cells/uL) Segmented neutrophils number (1000 cell/uL) WBC count (1000 cells/uL) (logged) Body Mass Index (kg/m2) (logged) Lymphocyte number (1000 cells/uL) Phosphorus (mmol/L) Bicarbonate (mmol/L) Albumin (g/L) Uric acid (umol/L) Total iron binding capacity (umol/L) 60 sec pulse (30 sec pulse X2) DBP average reported to examinee Folate serum (nmol/L) (logged) Albumin urine (mg/L) Low-Density Lipoprotein (mmol/L) Basophils percent Log Urine Albumin-to-Creatinine Ratio (mg/g) (logged) Segmented neutrophils percent Alanine Aminotransferase (ALT) (U/L) (logged) Healthcare use index Creatinine urine (umol/L) (logged) Basophils number (1000 cells/uL) Co-morbidity index CRP (mg/dL) (logged) |
| PC17M | 0.243 0.242 0.235 0.213 0.201 0.185 0.184 0.171 0.162 0.135 0.133 0.108 0.107 0.097 0.086 0.08 0.072 0.072 0.057 0.056 0.04 0.039 0.038 0.035 0.034 0.024 0.023 0.017 0.017 0.008 0.003 -0.455 -0.323 -0.283 -0.209 -0.117 -0.109 -0.094 -0.09 -0.079 -0.075 -0.058 -0.058 -0.054 -0.048 -0.043 -0.04 -0.035 -0.035 -0.028 -0.026 -0.024 -0.015 -0.012 -0.011 -0.008 -0.006 -0.005 -0.002 | LBDSGLSI LBXPLTSI LBXSC3SI URXUCRSI (logged) LBDSUASI BPXSAR LBXCOT LBXBAPCT BPXDAR LBDTIBSI LBDIRNSI LBDSCASI LBXRDW LBDSGBSI LBDSTPSI LBXMCVSI LBXPCT LBXEOPCT LDLV LBXMCHSI LBXNEPCT LBXSASSI (logged) BPXPLS LBDNENO LBDSCRSI LBDEONO SSBNP (logged) LBXGH fs3Score LBDBANO LBXSLDSI (logged) LBDB12SI (logged) LBDFOLSI (logged) fs2Score crAlbRat (logged) LBDFERSI (logged) LBXMPSI LBDMONO LBXSCLSI LBDSPHSI LBXRBCSI LBXMOPCT LBXSKSI LBDLYMNO LBDSBUSI LBXLYPCT URXUMASI LBXHGB LBXHCT LBDSTBSI LBXMC LBXSNASI BMXBMI (logged) LBDSALSI LBXWBCSI (logged) LBXCRP (logged) LBXSAPSI (logged) LBXSATSI (logged) fs1Score | Glucose (mmol/L) Platelet count (1000 cells/uL) Bicarbonate (mmol/L) Creatinine urine (umol/L) (logged) Uric acid (umol/L) SBP average reported to examinee Cotinine (ng/mL) Basophils percent DBP average reported to examinee Total iron binding capacity (umol/L) Iron (umol/L) Calcium total (mmol/L) Red cell distribution width (percent) Globulin (g/L) Protein total (g/L) Mean cell volume (fL) Transferrin Saturation (%) Eosinophils percent Low-Density Lipoprotein (mmol/L) Mean cell hemoglobin (pg) Segmented neutrophils percent Aspartate Aminotransferase (AST) (U/L) (logged) 60 sec pulse (30 sec pulse X2) Segmented neutrophils number (1000 cell/uL) Creatinine (umol/L) Eosinophils number (1000 cells/uL) NT-proBNP (pg/ml) (logged) Glycohemoglobin (%) Healthcare use index Basophils number (1000 cells/uL) Lactate Dehydrogenase (LDH) (U/L) (logged) Vitamin B12 serum (pmol/L) (logged) Folate serum (nmol/L) (logged) Self-health index Log Urine Albumin-to-Creatinine Ratio (mg/g) (logged) Ferritin (ug/L) (logged) Mean platelet volume (fL) Monocyte number (1000 cells/uL) Chloride (mmol/L) Phosphorus (mmol/L) Red blood cell count (million cells/uL) Monocyte percent Potassium (mmol/L) Lymphocyte number (1000 cells/uL) Blood Urea Nitrogen (mmol/L) Lymphocyte percent Albumin urine (mg/L) Hemoglobin (g/dL) Hematocrit Bilirubin total (umol/L) Mean Cell Hemoglobin Concentration (g/dL) Sodium (mmol/L) Body Mass Index (kg/m2) (logged) Albumin (g/L) WBC count (1000 cells/uL) (logged) CRP (mg/dL) (logged) Alkaline Phosphatase (ALP) (IU/L) (logged) Alanine Aminotransferase (ALT) (U/L) (logged) Co-morbidity index |
| PC19M | 0.503 0.185 0.142 0.116 0.101 0.098 0.088 0.086 0.077 0.075 0.075 0.071 0.07 0.063 0.054 0.052 0.052 0.049 0.038 0.037 0.035 0.031 0.029 0.027 0.027 0.004 0.003 -0.392 -0.3 -0.25 -0.172 -0.162 -0.162 -0.15 -0.147 -0.136 -0.136 -0.134 -0.128 -0.109 -0.106 -0.098 -0.091 -0.088 -0.088 -0.084 -0.083 -0.079 -0.056 -0.054 -0.048 -0.041 -0.038 -0.032 -0.028 -0.026 -0.017 -0.005 -0.001 | LBXMPSI LBDSCASI LBDSTPSI LBDSALSI LBDTIBSI LBXMCHSI LBXSASSI (logged) LBXMCVSI LBXMC LBXNEPCT LBDSPHSI crAlbRat (logged) LBDEONO LBDSGBSI LBXEOPCT LBXCOT LBDNENO LBXRDW LBXSATSI (logged) LBXGH LBXSNASI SSBNP (logged) LBXHGB LBXWBCSI (logged) LBXSC3SI LBXHCT URXUMASI LBXPLTSI LBDB12SI (logged) BPXDAR LBXMOPCT LBDMONO LBXSKSI LBDSBUSI LBXBAPCT LBDSGLSI LBDSTBSI LBXPCT URXUCRSI (logged) LBDFERSI (logged) LDLV LBXCRP (logged) LBDIRNSI BPXSAR fs2Score LBXSAPSI (logged) LBXSCLSI LBDSCRSI LBDFOLSI (logged) LBXRBCSI LBXLYPCT BMXBMI (logged) LBXSLDSI (logged) BPXPLS LBDLYMNO LBDSUASI fs3Score LBDBANO fs1Score | Mean platelet volume (fL) Calcium total (mmol/L) Protein total (g/L) Albumin (g/L) Total iron binding capacity (umol/L) Mean cell hemoglobin (pg) Aspartate Aminotransferase (AST) (U/L) (logged) Mean cell volume (fL) Mean Cell Hemoglobin Concentration (g/dL) Segmented neutrophils percent Phosphorus (mmol/L) Log Urine Albumin-to-Creatinine Ratio (mg/g) (logged) Eosinophils number (1000 cells/uL) Globulin (g/L) Eosinophils percent Cotinine (ng/mL) Segmented neutrophils number (1000 cell/uL) Red cell distribution width (percent) Alanine Aminotransferase (ALT) (U/L) (logged) Glycohemoglobin (%) Sodium (mmol/L) NT-proBNP (pg/ml) (logged) Hemoglobin (g/dL) WBC count (1000 cells/uL) (logged) Bicarbonate (mmol/L) Hematocrit Albumin urine (mg/L) Platelet count (1000 cells/uL) Vitamin B12 serum (pmol/L) (logged) DBP average reported to examinee Monocyte percent Monocyte number (1000 cells/uL) Potassium (mmol/L) Blood Urea Nitrogen (mmol/L) Basophils percent Glucose (mmol/L) Bilirubin total (umol/L) Transferrin Saturation (%) Creatinine urine (umol/L) (logged) Ferritin (ug/L) (logged) Low-Density Lipoprotein (mmol/L) CRP (mg/dL) (logged) Iron (umol/L) SBP average reported to examinee Self-health index Alkaline Phosphatase (ALP) (IU/L) (logged) Chloride (mmol/L) Creatinine (umol/L) Folate serum (nmol/L) (logged) Red blood cell count (million cells/uL) Lymphocyte percent Body Mass Index (kg/m2) (logged) Lactate Dehydrogenase (LDH) (U/L) (logged) 60 sec pulse (30 sec pulse X2) Lymphocyte number (1000 cells/uL) Uric acid (umol/L) Healthcare use index Basophils number (1000 cells/uL) Co-morbidity index |
| PC24M | 0.376 0.36 0.315 0.256 0.239 0.235 0.186 0.161 0.154 0.145 0.13 0.114 0.105 0.089 0.087 0.08 0.057 0.052 0.051 0.044 0.028 0.019 0.018 0.008 0.007 0.005 0.002 -0.18 -0.179 -0.176 -0.167 -0.145 -0.138 -0.135 -0.111 -0.098 -0.089 -0.085 -0.081 -0.079 -0.076 -0.074 -0.067 -0.063 -0.058 -0.051 -0.05 -0.047 -0.047 -0.047 -0.032 -0.031 -0.03 -0.028 -0.024 -0.023 -0.015 -0.012 -0.004 | LBXRDW LBDSPHSI LBXCOT LBXBAPCT BPXDAR LBDSTBSI LBDB12SI (logged) LBXMCVSI LBXCRP (logged) LBXMCHSI LBXSC3SI LBXGH LBXSKSI BPXPLS LBXHGB LBXHCT LBXMPSI LBDSBUSI LBXLYPCT crAlbRat (logged) LBXMC LBDFERSI (logged) LBDLYMNO LBDBANO URXUMASI BMXBMI (logged) LBDSCRSI fs2Score LBXPCT LBDIRNSI LBDSTPSI LBDMONO LBDSCASI LBDSGBSI LBXPLTSI LBXSCLSI LBDSGLSI LBDSUASI BPXSAR LBXMOPCT LBXSASSI (logged) fs3Score LBDFOLSI (logged) SSBNP (logged) LBDSALSI URXUCRSI (logged) LBDEONO LBXEOPCT LBXRBCSI LBXSLDSI (logged) LBXWBCSI (logged) LBXSATSI (logged) LBDNENO LBXNEPCT LBXSNASI LDLV LBXSAPSI (logged) LBDTIBSI fs1Score | Red cell distribution width (percent) Phosphorus (mmol/L) Cotinine (ng/mL) Basophils percent DBP average reported to examinee Bilirubin total (umol/L) Vitamin B12 serum (pmol/L) (logged) Mean cell volume (fL) CRP (mg/dL) (logged) Mean cell hemoglobin (pg) Bicarbonate (mmol/L) Glycohemoglobin (%) Potassium (mmol/L) 60 sec pulse (30 sec pulse X2) Hemoglobin (g/dL) Hematocrit Mean platelet volume (fL) Blood Urea Nitrogen (mmol/L) Lymphocyte percent Log Urine Albumin-to-Creatinine Ratio (mg/g) (logged) Mean Cell Hemoglobin Concentration (g/dL) Ferritin (ug/L) (logged) Lymphocyte number (1000 cells/uL) Basophils number (1000 cells/uL) Albumin urine (mg/L) Body Mass Index (kg/m2) (logged) Creatinine (umol/L) Self-health index Transferrin Saturation (%) Iron (umol/L) Protein total (g/L) Monocyte number (1000 cells/uL) Calcium total (mmol/L) Globulin (g/L) Platelet count (1000 cells/uL) Chloride (mmol/L) Glucose (mmol/L) Uric acid (umol/L) SBP average reported to examinee Monocyte percent Aspartate Aminotransferase (AST) (U/L) (logged) Healthcare use index Folate serum (nmol/L) (logged) NT-proBNP (pg/ml) (logged) Albumin (g/L) Creatinine urine (umol/L) (logged) Eosinophils number (1000 cells/uL) Eosinophils percent Red blood cell count (million cells/uL) Lactate Dehydrogenase (LDH) (U/L) (logged) WBC count (1000 cells/uL) (logged) Alanine Aminotransferase (ALT) (U/L) (logged) Segmented neutrophils number (1000 cell/uL) Segmented neutrophils percent Sodium (mmol/L) Low-Density Lipoprotein (mmol/L) Alkaline Phosphatase (ALP) (IU/L) (logged) Total iron binding capacity (umol/L) Co-morbidity index |
| PC25M | 0.409 0.313 0.302 0.278 0.237 0.201 0.153 0.137 0.124 0.119 0.112 0.101 0.09 0.09 0.075 0.069 0.053 0.05 0.048 0.03 0.025 0.019 0.016 0.014 0.009 0.009 0.007 0.006 0.003 -0.279 -0.263 -0.198 -0.163 -0.157 -0.15 -0.129 -0.094 -0.082 -0.077 -0.072 -0.068 -0.067 -0.067 -0.061 -0.057 -0.057 -0.05 -0.05 -0.045 -0.042 -0.029 -0.023 -0.021 -0.02 -0.017 -0.012 -0.008 -0.008 -0.006 | SSBNP (logged) LBXSKSI LBDFOLSI (logged) URXUCRSI (logged) LBXBAPCT LBXCRP (logged) BMXBMI (logged) LBDSPHSI LBXMPSI LBDSGBSI BPXSAR LBDSTPSI LBDFERSI (logged) LBDSUASI LBXMC LBDLYMNO LDLV LBXLYPCT URXUMASI LBXRBCSI LBXSC3SI LBDSCASI LBXSAPSI (logged) LBXWBCSI (logged) LBDBANO fs2Score LBXHGB fs1Score LBDTIBSI LBDSCRSI BPXPLS LBXSLDSI (logged) LBXSNASI crAlbRat (logged) LBXRDW LBXSCLSI LBXCOT fs3Score LBXPCT LBDIRNSI LBXEOPCT LBDB12SI (logged) LBXMCVSI LBXMOPCT LBDMONO LBDEONO LBXSASSI (logged) LBDSTBSI LBXPLTSI LBXGH LBXMCHSI LBDSBUSI LBXNEPCT LBXHCT LBDSALSI LBDNENO LBXSATSI (logged) LBDSGLSI BPXDAR | NT-proBNP (pg/ml) (logged) Potassium (mmol/L) Folate serum (nmol/L) (logged) Creatinine urine (umol/L) (logged) Basophils percent CRP (mg/dL) (logged) Body Mass Index (kg/m2) (logged) Phosphorus (mmol/L) Mean platelet volume (fL) Globulin (g/L) SBP average reported to examinee Protein total (g/L) Ferritin (ug/L) (logged) Uric acid (umol/L) Mean Cell Hemoglobin Concentration (g/dL) Lymphocyte number (1000 cells/uL) Low-Density Lipoprotein (mmol/L) Lymphocyte percent Albumin urine (mg/L) Red blood cell count (million cells/uL) Bicarbonate (mmol/L) Calcium total (mmol/L) Alkaline Phosphatase (ALP) (IU/L) (logged) WBC count (1000 cells/uL) (logged) Basophils number (1000 cells/uL) Self-health index Hemoglobin (g/dL) Co-morbidity index Total iron binding capacity (umol/L) Creatinine (umol/L) 60 sec pulse (30 sec pulse X2) Lactate Dehydrogenase (LDH) (U/L) (logged) Sodium (mmol/L) Log Urine Albumin-to-Creatinine Ratio (mg/g) (logged) Red cell distribution width (percent) Chloride (mmol/L) Cotinine (ng/mL) Healthcare use index Transferrin Saturation (%) Iron (umol/L) Eosinophils percent Vitamin B12 serum (pmol/L) (logged) Mean cell volume (fL) Monocyte percent Monocyte number (1000 cells/uL) Eosinophils number (1000 cells/uL) Aspartate Aminotransferase (AST) (U/L) (logged) Bilirubin total (umol/L) Platelet count (1000 cells/uL) Glycohemoglobin (%) Mean cell hemoglobin (pg) Blood Urea Nitrogen (mmol/L) Segmented neutrophils percent Hematocrit Albumin (g/L) Segmented neutrophils number (1000 cell/uL) Alanine Aminotransferase (ALT) (U/L) (logged) Glucose (mmol/L) DBP average reported to examinee |
| PC27M | 0.43 0.273 0.244 0.203 0.166 0.125 0.12 0.115 0.108 0.099 0.098 0.098 0.096 0.091 0.079 0.078 0.071 0.058 0.051 0.048 0.037 0.032 0.03 0.026 0.018 0.016 0.016 0.014 0.011 0.002 0 -0.302 -0.292 -0.214 -0.2 -0.197 -0.196 -0.196 -0.19 -0.133 -0.125 -0.09 -0.063 -0.059 -0.055 -0.054 -0.049 -0.043 -0.039 -0.038 -0.036 -0.028 -0.024 -0.017 -0.017 -0.016 -0.013 -0.007 -0.004 | LBXBAPCT LBDFOLSI (logged) LBDSGLSI LBXCOT LBDSCRSI LBXMPSI fs2Score LBDB12SI (logged) LBXSASSI (logged) LBDTIBSI BPXSAR LBDSALSI LBDSUASI LBDSBUSI LBDSTPSI LBXRDW LBDMONO LBDNENO LBDLYMNO LBXWBCSI (logged) URXUMASI LBXSATSI (logged) URXUCRSI (logged) LBXRBCSI LBXHGB LBDBANO LBXHCT LBDSGBSI LBXMOPCT LBXMC LBXLYPCT LBXSKSI LBDSCASI BMXBMI (logged) LBXGH LBDSPHSI BPXDAR fs3Score LBXSLDSI (logged) LBXCRP (logged) SSBNP (logged) LBDFERSI (logged) LBXPCT LBXSC3SI crAlbRat (logged) LBXSCLSI LBXSAPSI (logged) LDLV LBXPLTSI LBXSNASI LBXEOPCT BPXPLS LBDEONO LBXMCVSI LBDSTBSI LBDIRNSI LBXMCHSI LBXNEPCT fs1Score | Basophils percent Folate serum (nmol/L) (logged) Glucose (mmol/L) Cotinine (ng/mL) Creatinine (umol/L) Mean platelet volume (fL) Self-health index Vitamin B12 serum (pmol/L) (logged) Aspartate Aminotransferase (AST) (U/L) (logged) Total iron binding capacity (umol/L) SBP average reported to examinee Albumin (g/L) Uric acid (umol/L) Blood Urea Nitrogen (mmol/L) Protein total (g/L) Red cell distribution width (percent) Monocyte number (1000 cells/uL) Segmented neutrophils number (1000 cell/uL) Lymphocyte number (1000 cells/uL) WBC count (1000 cells/uL) (logged) Albumin urine (mg/L) Alanine Aminotransferase (ALT) (U/L) (logged) Creatinine urine (umol/L) (logged) Red blood cell count (million cells/uL) Hemoglobin (g/dL) Basophils number (1000 cells/uL) Hematocrit Globulin (g/L) Monocyte percent Mean Cell Hemoglobin Concentration (g/dL) Lymphocyte percent Potassium (mmol/L) Calcium total (mmol/L) Body Mass Index (kg/m2) (logged) Glycohemoglobin (%) Phosphorus (mmol/L) DBP average reported to examinee Healthcare use index Lactate Dehydrogenase (LDH) (U/L) (logged) CRP (mg/dL) (logged) NT-proBNP (pg/ml) (logged) Ferritin (ug/L) (logged) Transferrin Saturation (%) Bicarbonate (mmol/L) Log Urine Albumin-to-Creatinine Ratio (mg/g) (logged) Chloride (mmol/L) Alkaline Phosphatase (ALP) (IU/L) (logged) Low-Density Lipoprotein (mmol/L) Platelet count (1000 cells/uL) Sodium (mmol/L) Eosinophils percent 60 sec pulse (30 sec pulse X2) Eosinophils number (1000 cells/uL) Mean cell volume (fL) Bilirubin total (umol/L) Iron (umol/L) Mean cell hemoglobin (pg) Segmented neutrophils percent Co-morbidity index |
| PC31M | 0.521 0.312 0.195 0.193 0.182 0.172 0.171 0.142 0.131 0.11 0.099 0.097 0.085 0.062 0.052 0.052 0.049 0.039 0.035 0.034 0.032 0.02 0.017 0.011 0.008 0.007 0.003 0.001 0.001 0.001 -0.373 -0.255 -0.191 -0.148 -0.12 -0.116 -0.109 -0.099 -0.099 -0.083 -0.08 -0.079 -0.067 -0.065 -0.061 -0.056 -0.056 -0.051 -0.045 -0.044 -0.033 -0.033 -0.029 -0.027 -0.025 -0.015 -0.006 -0.006 -0.003 | SSBNP (logged) LBDB12SI (logged) LBDTIBSI LBDSUASI LBXSLDSI (logged) BPXPLS LBDSCASI LBXBAPCT LBDIRNSI LBXSNASI LBXMC LBXCOT LBXGH BPXDAR LBXPCT LBXCRP (logged) LBXRBCSI LBXHGB LBXMOPCT URXUCRSI (logged) fs2Score LDLV BMXBMI (logged) LBXLYPCT LBDSALSI fs1Score LBDBANO LBDEONO LBXHCT LBXSCLSI LBXSKSI BPXSAR LBDSBUSI LBXSAPSI (logged) LBDSTBSI LBDSGBSI LBDSTPSI LBDFERSI (logged) LBXSATSI (logged) LBXSASSI (logged) fs3Score LBDFOLSI (logged) LBDSCRSI LBXMCVSI crAlbRat (logged) LBDNENO LBXRDW LBXMPSI LBXPLTSI LBXSC3SI LBXWBCSI (logged) LBDSPHSI LBXNEPCT LBDLYMNO LBDSGLSI LBXMCHSI LBXEOPCT LBDMONO URXUMASI | NT-proBNP (pg/ml) (logged) Vitamin B12 serum (pmol/L) (logged) Total iron binding capacity (umol/L) Uric acid (umol/L) Lactate Dehydrogenase (LDH) (U/L) (logged) 60 sec pulse (30 sec pulse X2) Calcium total (mmol/L) Basophils percent Iron (umol/L) Sodium (mmol/L) Mean Cell Hemoglobin Concentration (g/dL) Cotinine (ng/mL) Glycohemoglobin (%) DBP average reported to examinee Transferrin Saturation (%) CRP (mg/dL) (logged) Red blood cell count (million cells/uL) Hemoglobin (g/dL) Monocyte percent Creatinine urine (umol/L) (logged) Self-health index Low-Density Lipoprotein (mmol/L) Body Mass Index (kg/m2) (logged) Lymphocyte percent Albumin (g/L) Co-morbidity index Basophils number (1000 cells/uL) Eosinophils number (1000 cells/uL) Hematocrit Chloride (mmol/L) Potassium (mmol/L) SBP average reported to examinee Blood Urea Nitrogen (mmol/L) Alkaline Phosphatase (ALP) (IU/L) (logged) Bilirubin total (umol/L) Globulin (g/L) Protein total (g/L) Ferritin (ug/L) (logged) Alanine Aminotransferase (ALT) (U/L) (logged) Aspartate Aminotransferase (AST) (U/L) (logged) Healthcare use index Folate serum (nmol/L) (logged) Creatinine (umol/L) Mean cell volume (fL) Log Urine Albumin-to-Creatinine Ratio (mg/g) (logged) Segmented neutrophils number (1000 cell/uL) Red cell distribution width (percent) Mean platelet volume (fL) Platelet count (1000 cells/uL) Bicarbonate (mmol/L) WBC count (1000 cells/uL) (logged) Phosphorus (mmol/L) Segmented neutrophils percent Lymphocyte number (1000 cells/uL) Glucose (mmol/L) Mean cell hemoglobin (pg) Eosinophils percent Monocyte number (1000 cells/uL) Albumin urine (mg/L) |
| PC33M | 0.411 0.207 0.187 0.133 0.116 0.114 0.107 0.105 0.104 0.103 0.091 0.084 0.082 0.081 0.078 0.076 0.062 0.051 0.046 0.017 0.016 0.014 0.003 0.003 0.002 0.001 0 -0.457 -0.262 -0.251 -0.186 -0.172 -0.171 -0.156 -0.155 -0.153 -0.152 -0.125 -0.124 -0.115 -0.109 -0.074 -0.068 -0.065 -0.056 -0.051 -0.051 -0.049 -0.043 -0.041 -0.041 -0.033 -0.015 -0.014 -0.013 -0.01 -0.008 -0.006 -0.006 | LBDFOLSI (logged) LBXMCVSI LBXCOT LBXSASSI (logged) LBXHCT LBDSUASI LBXCRP (logged) LBXPLTSI LBDSCRSI LBDSPHSI LDLV URXUCRSI (logged) LBXSATSI (logged) LBXGH LBDSCASI fs2Score LBXEOPCT LBXLYPCT BPXDAR LBDEONO BPXSAR SSBNP (logged) BPXPLS fs1Score URXUMASI LBDSGLSI LBXMCHSI LBXMC LBXBAPCT LBDSTPSI LBDSALSI LBDB12SI (logged) LBXSKSI fs3Score LBDTIBSI LBDFERSI (logged) LBXRDW LBDSBUSI LBDSGBSI LBXSAPSI (logged) LBDMONO LBDIRNSI LBXSCLSI LBXSC3SI LBXSNASI LBXRBCSI LBXHGB LBXMOPCT LBXWBCSI (logged) LBDNENO LBXMPSI LBXNEPCT BMXBMI (logged) LBXSLDSI (logged) LBXPCT LBDBANO crAlbRat (logged) LBDLYMNO LBDSTBSI | Folate serum (nmol/L) (logged) Mean cell volume (fL) Cotinine (ng/mL) Aspartate Aminotransferase (AST) (U/L) (logged) Hematocrit Uric acid (umol/L) CRP (mg/dL) (logged) Platelet count (1000 cells/uL) Creatinine (umol/L) Phosphorus (mmol/L) Low-Density Lipoprotein (mmol/L) Creatinine urine (umol/L) (logged) Alanine Aminotransferase (ALT) (U/L) (logged) Glycohemoglobin (%) Calcium total (mmol/L) Self-health index Eosinophils percent Lymphocyte percent DBP average reported to examinee Eosinophils number (1000 cells/uL) SBP average reported to examinee NT-proBNP (pg/ml) (logged) 60 sec pulse (30 sec pulse X2) Co-morbidity index Albumin urine (mg/L) Glucose (mmol/L) Mean cell hemoglobin (pg) Mean Cell Hemoglobin Concentration (g/dL) Basophils percent Protein total (g/L) Albumin (g/L) Vitamin B12 serum (pmol/L) (logged) Potassium (mmol/L) Healthcare use index Total iron binding capacity (umol/L) Ferritin (ug/L) (logged) Red cell distribution width (percent) Blood Urea Nitrogen (mmol/L) Globulin (g/L) Alkaline Phosphatase (ALP) (IU/L) (logged) Monocyte number (1000 cells/uL) Iron (umol/L) Chloride (mmol/L) Bicarbonate (mmol/L) Sodium (mmol/L) Red blood cell count (million cells/uL) Hemoglobin (g/dL) Monocyte percent WBC count (1000 cells/uL) (logged) Segmented neutrophils number (1000 cell/uL) Mean platelet volume (fL) Segmented neutrophils percent Body Mass Index (kg/m2) (logged) Lactate Dehydrogenase (LDH) (U/L) (logged) Transferrin Saturation (%) Basophils number (1000 cells/uL) Log Urine Albumin-to-Creatinine Ratio (mg/g) (logged) Lymphocyte number (1000 cells/uL) Bilirubin total (umol/L) |
| PC36M | 0.349 0.277 0.232 0.219 0.201 0.173 0.153 0.137 0.137 0.134 0.115 0.112 0.097 0.048 0.042 0.04 0.039 0.033 0.032 0.031 0.03 0.023 0.021 0.018 0.018 0.015 0.014 0.012 0.005 0.004 0.002 0.001 -0.269 -0.251 -0.244 -0.228 -0.206 -0.189 -0.171 -0.161 -0.156 -0.149 -0.129 -0.127 -0.119 -0.085 -0.067 -0.062 -0.052 -0.046 -0.045 -0.042 -0.036 -0.028 -0.024 -0.022 -0.005 -0.004 -0.004 | LBXMC LBXCOT LBXGH LBDSCRSI LDLV BPXPLS LBXSASSI (logged) BPXSAR LBXSATSI (logged) LBXSKSI LBDSGBSI LBXCRP (logged) LBXSC3SI LBXRBCSI LBXPCT LBDIRNSI LBXMOPCT LBDSTBSI LBDSCASI URXUCRSI (logged) BMXBMI (logged) LBDFOLSI (logged) LBDNENO LBXNEPCT fs2Score LBXHGB LBXEOPCT LBDB12SI (logged) LBDMONO fs1Score LBDBANO LBXSCLSI LBXSAPSI (logged) LBDSGLSI LBXSLDSI (logged) LBXMCVSI LBDSALSI LBXPLTSI LBXMPSI LBDSPHSI LBDSUASI LBDFERSI (logged) LBXRDW LBXHCT LBDSBUSI BPXDAR SSBNP (logged) LBXMCHSI crAlbRat (logged) fs3Score LBXSNASI LBDLYMNO LBDTIBSI LBXLYPCT LBDSTPSI LBXBAPCT LBDEONO URXUMASI LBXWBCSI (logged) | Mean Cell Hemoglobin Concentration (g/dL) Cotinine (ng/mL) Glycohemoglobin (%) Creatinine (umol/L) Low-Density Lipoprotein (mmol/L) 60 sec pulse (30 sec pulse X2) Aspartate Aminotransferase (AST) (U/L) (logged) SBP average reported to examinee Alanine Aminotransferase (ALT) (U/L) (logged) Potassium (mmol/L) Globulin (g/L) CRP (mg/dL) (logged) Bicarbonate (mmol/L) Red blood cell count (million cells/uL) Transferrin Saturation (%) Iron (umol/L) Monocyte percent Bilirubin total (umol/L) Calcium total (mmol/L) Creatinine urine (umol/L) (logged) Body Mass Index (kg/m2) (logged) Folate serum (nmol/L) (logged) Segmented neutrophils number (1000 cell/uL) Segmented neutrophils percent Self-health index Hemoglobin (g/dL) Eosinophils percent Vitamin B12 serum (pmol/L) (logged) Monocyte number (1000 cells/uL) Co-morbidity index Basophils number (1000 cells/uL) Chloride (mmol/L) Alkaline Phosphatase (ALP) (IU/L) (logged) Glucose (mmol/L) Lactate Dehydrogenase (LDH) (U/L) (logged) Mean cell volume (fL) Albumin (g/L) Platelet count (1000 cells/uL) Mean platelet volume (fL) Phosphorus (mmol/L) Uric acid (umol/L) Ferritin (ug/L) (logged) Red cell distribution width (percent) Hematocrit Blood Urea Nitrogen (mmol/L) DBP average reported to examinee NT-proBNP (pg/ml) (logged) Mean cell hemoglobin (pg) Log Urine Albumin-to-Creatinine Ratio (mg/g) (logged) Healthcare use index Sodium (mmol/L) Lymphocyte number (1000 cells/uL) Total iron binding capacity (umol/L) Lymphocyte percent Protein total (g/L) Basophils percent Eosinophils number (1000 cells/uL) Albumin urine (mg/L) WBC count (1000 cells/uL) (logged) |
| PC42M | 0.289 0.285 0.248 0.185 0.141 0.141 0.094 0.083 0.077 0.069 0.065 0.065 0.054 0.052 0.048 0.046 0.035 0.033 0.031 0.028 0.023 0.021 0.02 0.017 0.012 0.01 0.01 0.002 0.001 0 -0.514 -0.429 -0.258 -0.192 -0.151 -0.11 -0.097 -0.095 -0.09 -0.084 -0.074 -0.073 -0.062 -0.057 -0.053 -0.046 -0.044 -0.04 -0.04 -0.016 -0.016 -0.013 -0.012 -0.01 -0.007 -0.007 -0.006 -0.005 -0.002 | LBDTIBSI LBDFERSI (logged) LBDSBUSI LBDSCASI LBDSUASI LBDSGBSI LBXSC3SI BPXSAR LBXCOT LBDNENO BPXPLS LBXSCLSI LBXSAPSI (logged) LBXEOPCT LBDB12SI (logged) LBDFOLSI (logged) LBDSTBSI LBXSLDSI (logged) URXUMASI LBXBAPCT LBXMCVSI fs2Score BMXBMI (logged) LBDEONO LBXNEPCT LBXWBCSI (logged) LBXGH LBDBANO LBXHCT LBDIRNSI LBDSALSI LBXCRP (logged) LBDSCRSI LBDSTPSI LBXPCT LBDSGLSI LBDSPHSI fs3Score LBXPLTSI SSBNP (logged) LBXSNASI LBXSATSI (logged) BPXDAR LBXMC LBXSASSI (logged) LBXMPSI LBXRDW LBXMOPCT LBDMONO URXUCRSI (logged) LBXHGB LBXLYPCT LBXSKSI LBXRBCSI LBDLYMNO crAlbRat (logged) LDLV LBXMCHSI fs1Score | Total iron binding capacity (umol/L) Ferritin (ug/L) (logged) Blood Urea Nitrogen (mmol/L) Calcium total (mmol/L) Uric acid (umol/L) Globulin (g/L) Bicarbonate (mmol/L) SBP average reported to examinee Cotinine (ng/mL) Segmented neutrophils number (1000 cell/uL) 60 sec pulse (30 sec pulse X2) Chloride (mmol/L) Alkaline Phosphatase (ALP) (IU/L) (logged) Eosinophils percent Vitamin B12 serum (pmol/L) (logged) Folate serum (nmol/L) (logged) Bilirubin total (umol/L) Lactate Dehydrogenase (LDH) (U/L) (logged) Albumin urine (mg/L) Basophils percent Mean cell volume (fL) Self-health index Body Mass Index (kg/m2) (logged) Eosinophils number (1000 cells/uL) Segmented neutrophils percent WBC count (1000 cells/uL) (logged) Glycohemoglobin (%) Basophils number (1000 cells/uL) Hematocrit Iron (umol/L) Albumin (g/L) CRP (mg/dL) (logged) Creatinine (umol/L) Protein total (g/L) Transferrin Saturation (%) Glucose (mmol/L) Phosphorus (mmol/L) Healthcare use index Platelet count (1000 cells/uL) NT-proBNP (pg/ml) (logged) Sodium (mmol/L) Alanine Aminotransferase (ALT) (U/L) (logged) DBP average reported to examinee Mean Cell Hemoglobin Concentration (g/dL) Aspartate Aminotransferase (AST) (U/L) (logged) Mean platelet volume (fL) Red cell distribution width (percent) Monocyte percent Monocyte number (1000 cells/uL) Creatinine urine (umol/L) (logged) Hemoglobin (g/dL) Lymphocyte percent Potassium (mmol/L) Red blood cell count (million cells/uL) Lymphocyte number (1000 cells/uL) Log Urine Albumin-to-Creatinine Ratio (mg/g) (logged) Low-Density Lipoprotein (mmol/L) Mean cell hemoglobin (pg) Co-morbidity index |

| **PC (Female)** | **PC Loadings** | **Variable Names** | **Parameters** |
| --- | --- | --- | --- |
| PC1F | 0.479 0.327 0.317 0.314 0.267 0.259 0.23 0.21 0.182 0.172 0.128 0.112 0.107 0.1 0.096 0.091 0.089 0.085 0.082 0.081 0.079 0.074 0.064 0.063 0.062 0.058 0.056 0.055 0.053 0.051 0.049 0.042 0.034 0.03 0.03 0.029 0.025 0.025 0.024 0.023 0.02 0.017 0.015 0.015 0.014 0.012 0.01 0.008 -0.083 -0.065 -0.064 -0.052 -0.033 -0.023 -0.022 -0.018 -0.007 -0.004 -0.001 | fs3Score LBXGH URXUMASI LBDSGLSI BPXSAR crAlbRat (logged) LBDSBUSI fs2Score SSBNP (logged) LBDSUASI LBXSC3SI LBDFERSI (logged) LBDEONO LBXRDW LBDSCRSI LBDSTBSI LBXSLDSI (logged) LBDFOLSI (logged) LBXSAPSI (logged) LBXSASSI (logged) LBDMONO LBXSKSI LBDSGBSI LBXSATSI (logged) LBXCOT LBXMOPCT LDLV LBDB12SI (logged) LBXEOPCT LBDSCASI LBXCRP (logged) LBXMPSI LBDNENO LBDLYMNO LBXHCT LBDSTPSI BMXBMI (logged) fs1Score LBXRBCSI LBDSPHSI LBXWBCSI (logged) LBXNEPCT BPXPLS LBXPCT LBXHGB LBDIRNSI LBXSNASI LBDBANO BPXDAR URXUCRSI (logged) LBXSCLSI LBDSALSI LBDTIBSI LBXLYPCT LBXMC LBXMCHSI LBXPLTSI LBXMCVSI LBXBAPCT | Healthcare use index Glycohemoglobin (%) Albumin urine (mg/L) Glucose (mmol/L) SBP average reported to examinee Log Urine Albumin-to-Creatinine Ratio (mg/g) (logged) Blood Urea Nitrogen (mmol/L) Self-health index NT-proBNP (pg/ml) (logged) Uric acid (umol/L) Bicarbonate (mmol/L) Ferritin (ug/L) (logged) Eosinophils number (1000 cells/uL) Red cell distribution width (percent) Creatinine (umol/L) Bilirubin total (umol/L) Lactate Dehydrogenase (LDH) (U/L) (logged) Folate serum (nmol/L) (logged) Alkaline Phosphatase (ALP) (IU/L) (logged) Aspartate Aminotransferase (AST) (U/L) (logged) Monocyte number (1000 cells/uL) Potassium (mmol/L) Globulin (g/L) Alanine Aminotransferase (ALT) (U/L) (logged) Cotinine (ng/mL) Monocyte percent Low-Density Lipoprotein (mmol/L) Vitamin B12 serum (pmol/L) (logged) Eosinophils percent Calcium total (mmol/L) CRP (mg/dL) (logged) Mean platelet volume (fL) Segmented neutrophils number (1000 cell/uL) Lymphocyte number (1000 cells/uL) Hematocrit Protein total (g/L) Body Mass Index (kg/m2) (logged) Co-morbidity index Red blood cell count (million cells/uL) Phosphorus (mmol/L) WBC count (1000 cells/uL) (logged) Segmented neutrophils percent 60 sec pulse (30 sec pulse X2) Transferrin Saturation (%) Hemoglobin (g/dL) Iron (umol/L) Sodium (mmol/L) Basophils number (1000 cells/uL) DBP average reported to examinee Creatinine urine (umol/L) (logged) Chloride (mmol/L) Albumin (g/L) Total iron binding capacity (umol/L) Lymphocyte percent Mean Cell Hemoglobin Concentration (g/dL) Mean cell hemoglobin (pg) Platelet count (1000 cells/uL) Mean cell volume (fL) Basophils percent |
| PC2F | 0.339 0.204 0.191 0.19 0.189 0.185 0.178 0.171 0.167 0.157 0.114 0.109 0.101 0.094 0.09 0.089 0.087 0.083 0.083 0.078 0.074 0.067 0.059 0.058 0.055 0.047 0.045 0.04 0.034 0.032 0.03 0.028 0.013 0.01 0.009 0.004 -0.338 -0.302 -0.272 -0.154 -0.146 -0.135 -0.133 -0.124 -0.122 -0.117 -0.106 -0.104 -0.097 -0.094 -0.084 -0.074 -0.067 -0.06 -0.051 -0.037 -0.029 -0.01 -0.005 | fs3Score LBDSTBSI LBXPCT LBDIRNSI LBXSC3SI LBXSASSI (logged) LBDFOLSI (logged) LBXMCVSI LBXMCHSI SSBNP (logged) LBDSBUSI LBXMOPCT LBXSKSI LBXMC LBDFERSI (logged) LBDB12SI (logged) BPXSAR LBXSATSI (logged) LBXSNASI LBXHGB LBDSALSI LBXSLDSI (logged) LBXEOPCT LBXBAPCT LBXHCT LBDSCASI LBXSCLSI LBDSCRSI LBDEONO LBXCOT fs2Score LBDSPHSI LDLV LBXLYPCT fs1Score LBDBANO LBDSGLSI LBXGH URXUMASI crAlbRat (logged) LBXWBCSI (logged) URXUCRSI (logged) LBDSGBSI BMXBMI (logged) LBDNENO LBXCRP (logged) LBXRDW LBXPLTSI BPXPLS LBDLYMNO LBDSTPSI LBXSAPSI (logged) LBXRBCSI LBDMONO BPXDAR LBDTIBSI LBXNEPCT LBXMPSI LBDSUASI | Healthcare use index Bilirubin total (umol/L) Transferrin Saturation (%) Iron (umol/L) Bicarbonate (mmol/L) Aspartate Aminotransferase (AST) (U/L) (logged) Folate serum (nmol/L) (logged) Mean cell volume (fL) Mean cell hemoglobin (pg) NT-proBNP (pg/ml) (logged) Blood Urea Nitrogen (mmol/L) Monocyte percent Potassium (mmol/L) Mean Cell Hemoglobin Concentration (g/dL) Ferritin (ug/L) (logged) Vitamin B12 serum (pmol/L) (logged) SBP average reported to examinee Alanine Aminotransferase (ALT) (U/L) (logged) Sodium (mmol/L) Hemoglobin (g/dL) Albumin (g/L) Lactate Dehydrogenase (LDH) (U/L) (logged) Eosinophils percent Basophils percent Hematocrit Calcium total (mmol/L) Chloride (mmol/L) Creatinine (umol/L) Eosinophils number (1000 cells/uL) Cotinine (ng/mL) Self-health index Phosphorus (mmol/L) Low-Density Lipoprotein (mmol/L) Lymphocyte percent Co-morbidity index Basophils number (1000 cells/uL) Glucose (mmol/L) Glycohemoglobin (%) Albumin urine (mg/L) Log Urine Albumin-to-Creatinine Ratio (mg/g) (logged) WBC count (1000 cells/uL) (logged) Creatinine urine (umol/L) (logged) Globulin (g/L) Body Mass Index (kg/m2) (logged) Segmented neutrophils number (1000 cell/uL) CRP (mg/dL) (logged) Red cell distribution width (percent) Platelet count (1000 cells/uL) 60 sec pulse (30 sec pulse X2) Lymphocyte number (1000 cells/uL) Protein total (g/L) Alkaline Phosphatase (ALP) (IU/L) (logged) Red blood cell count (million cells/uL) Monocyte number (1000 cells/uL) DBP average reported to examinee Total iron binding capacity (umol/L) Segmented neutrophils percent Mean platelet volume (fL) Uric acid (umol/L) |
| PC4F | 0.339 0.24 0.197 0.098 0.091 0.087 0.086 0.082 0.082 0.076 0.072 0.059 0.058 0.053 0.042 0.037 0.036 0.029 0.024 0.021 0.016 0.015 0.014 0.012 0.011 0.006 0.004 0.002 -0.494 -0.388 -0.227 -0.191 -0.185 -0.178 -0.173 -0.167 -0.142 -0.141 -0.11 -0.1 -0.099 -0.089 -0.073 -0.067 -0.063 -0.06 -0.057 -0.052 -0.046 -0.044 -0.023 -0.017 -0.016 -0.014 -0.014 -0.013 -0.012 -0.009 -0.004 | fs3Score LBXRDW fs2Score LBDMONO LBDEONO LBXGH LBXMOPCT LBXEOPCT LBXCRP (logged) LBDSGBSI LBDLYMNO BMXBMI (logged) LBDTIBSI LBXPLTSI LBDSUASI LBXLYPCT LBXMPSI LBXCOT LBXWBCSI (logged) LBDSBUSI LBDSCRSI LBDSPHSI LBXSC3SI fs1Score SSBNP (logged) LBDNENO LBDBANO LBXSKSI URXUMASI crAlbRat (logged) LBXHGB LBXHCT LBXPCT LBDIRNSI LBXMCHSI LBXMCVSI LBDSTBSI LBDSALSI LBXSASSI (logged) LBXSATSI (logged) LBXMC LBDFERSI (logged) URXUCRSI (logged) LBXRBCSI BPXDAR LBXSNASI LBDFOLSI (logged) LBXNEPCT BPXSAR LBDSCASI LBXSCLSI LBDSTPSI LDLV LBXBAPCT LBDSGLSI BPXPLS LBXSLDSI (logged) LBDB12SI (logged) LBXSAPSI (logged) | Healthcare use index Red cell distribution width (percent) Self-health index Monocyte number (1000 cells/uL) Eosinophils number (1000 cells/uL) Glycohemoglobin (%) Monocyte percent Eosinophils percent CRP (mg/dL) (logged) Globulin (g/L) Lymphocyte number (1000 cells/uL) Body Mass Index (kg/m2) (logged) Total iron binding capacity (umol/L) Platelet count (1000 cells/uL) Uric acid (umol/L) Lymphocyte percent Mean platelet volume (fL) Cotinine (ng/mL) WBC count (1000 cells/uL) (logged) Blood Urea Nitrogen (mmol/L) Creatinine (umol/L) Phosphorus (mmol/L) Bicarbonate (mmol/L) Co-morbidity index NT-proBNP (pg/ml) (logged) Segmented neutrophils number (1000 cell/uL) Basophils number (1000 cells/uL) Potassium (mmol/L) Albumin urine (mg/L) Log Urine Albumin-to-Creatinine Ratio (mg/g) (logged) Hemoglobin (g/dL) Hematocrit Transferrin Saturation (%) Iron (umol/L) Mean cell hemoglobin (pg) Mean cell volume (fL) Bilirubin total (umol/L) Albumin (g/L) Aspartate Aminotransferase (AST) (U/L) (logged) Alanine Aminotransferase (ALT) (U/L) (logged) Mean Cell Hemoglobin Concentration (g/dL) Ferritin (ug/L) (logged) Creatinine urine (umol/L) (logged) Red blood cell count (million cells/uL) DBP average reported to examinee Sodium (mmol/L) Folate serum (nmol/L) (logged) Segmented neutrophils percent SBP average reported to examinee Calcium total (mmol/L) Chloride (mmol/L) Protein total (g/L) Low-Density Lipoprotein (mmol/L) Basophils percent Glucose (mmol/L) 60 sec pulse (30 sec pulse X2) Lactate Dehydrogenase (LDH) (U/L) (logged) Vitamin B12 serum (pmol/L) (logged) Alkaline Phosphatase (ALP) (IU/L) (logged) |
| PC6F | 0.295 0.293 0.29 0.259 0.249 0.23 0.194 0.184 0.183 0.168 0.133 0.131 0.119 0.113 0.111 0.096 0.089 0.085 0.083 0.08 0.076 0.06 0.057 0.056 0.051 0.046 0.041 0.038 0.036 0.034 0.019 0.017 0.009 0.003 0.003 0 -0.291 -0.176 -0.176 -0.154 -0.136 -0.128 -0.115 -0.112 -0.092 -0.088 -0.085 -0.079 -0.078 -0.077 -0.065 -0.055 -0.049 -0.047 -0.046 -0.033 -0.025 -0.024 -0.022 | LBXWBCSI (logged) LBDSBUSI LBDNENO LBDSUASI LBDSCRSI LBDMONO LBXHGB SSBNP (logged) LBXNEPCT LBXHCT LBXCRP (logged) LBXSKSI LBXMCHSI LBXMCVSI LBDLYMNO LBDSCASI LBXCOT LBXPLTSI LBDFERSI (logged) LBXRBCSI LBXMC BPXPLS LBDEONO LBDSPHSI LDLV BMXBMI (logged) LBXSAPSI (logged) LBXPCT LBDIRNSI LBDSGBSI fs2Score LBDSTPSI URXUCRSI (logged) LBXMPSI fs1Score LBDBANO LBXSC3SI LBXLYPCT LBDSGLSI LBXBAPCT LBXSASSI (logged) LBXGH URXUMASI LBXMOPCT LBXSATSI (logged) crAlbRat (logged) BPXDAR LBDSTBSI fs3Score LBDB12SI (logged) LBXRDW LBXSNASI LBXEOPCT LBXSLDSI (logged) LBDFOLSI (logged) LBXSCLSI LBDTIBSI LBDSALSI BPXSAR | WBC count (1000 cells/uL) (logged) Blood Urea Nitrogen (mmol/L) Segmented neutrophils number (1000 cell/uL) Uric acid (umol/L) Creatinine (umol/L) Monocyte number (1000 cells/uL) Hemoglobin (g/dL) NT-proBNP (pg/ml) (logged) Segmented neutrophils percent Hematocrit CRP (mg/dL) (logged) Potassium (mmol/L) Mean cell hemoglobin (pg) Mean cell volume (fL) Lymphocyte number (1000 cells/uL) Calcium total (mmol/L) Cotinine (ng/mL) Platelet count (1000 cells/uL) Ferritin (ug/L) (logged) Red blood cell count (million cells/uL) Mean Cell Hemoglobin Concentration (g/dL) 60 sec pulse (30 sec pulse X2) Eosinophils number (1000 cells/uL) Phosphorus (mmol/L) Low-Density Lipoprotein (mmol/L) Body Mass Index (kg/m2) (logged) Alkaline Phosphatase (ALP) (IU/L) (logged) Transferrin Saturation (%) Iron (umol/L) Globulin (g/L) Self-health index Protein total (g/L) Creatinine urine (umol/L) (logged) Mean platelet volume (fL) Co-morbidity index Basophils number (1000 cells/uL) Bicarbonate (mmol/L) Lymphocyte percent Glucose (mmol/L) Basophils percent Aspartate Aminotransferase (AST) (U/L) (logged) Glycohemoglobin (%) Albumin urine (mg/L) Monocyte percent Alanine Aminotransferase (ALT) (U/L) (logged) Log Urine Albumin-to-Creatinine Ratio (mg/g) (logged) DBP average reported to examinee Bilirubin total (umol/L) Healthcare use index Vitamin B12 serum (pmol/L) (logged) Red cell distribution width (percent) Sodium (mmol/L) Eosinophils percent Lactate Dehydrogenase (LDH) (U/L) (logged) Folate serum (nmol/L) (logged) Chloride (mmol/L) Total iron binding capacity (umol/L) Albumin (g/L) SBP average reported to examinee |
| PC11F | 0.539 0.342 0.222 0.139 0.122 0.111 0.109 0.085 0.08 0.061 0.059 0.053 0.05 0.046 0.043 0.041 0.037 0.037 0.021 0.021 0.019 0.016 0.013 0.009 0.004 0.003 0.002 -0.333 -0.222 -0.22 -0.216 -0.177 -0.169 -0.13 -0.126 -0.113 -0.11 -0.105 -0.095 -0.092 -0.09 -0.085 -0.083 -0.076 -0.066 -0.054 -0.052 -0.05 -0.049 -0.046 -0.038 -0.024 -0.018 -0.017 -0.007 -0.007 -0.007 -0.007 -0.001 | BPXSAR BPXDAR fs2Score SSBNP (logged) LBXMCHSI LBXMCVSI LBDSGBSI LBXMC LBDSTPSI LBXPCT LBXLYPCT LBDFERSI (logged) LBXPLTSI LBDLYMNO LBDIRNSI LBXCRP (logged) LBXSLDSI (logged) LBDSTBSI BMXBMI (logged) crAlbRat (logged) BPXPLS LBXSAPSI (logged) LBDNENO LBXWBCSI (logged) fs1Score LBXNEPCT LBXMOPCT LBXEOPCT LBXSKSI LBDEONO LBDSBUSI LBXBAPCT LBXRBCSI LBXSCLSI LBXSNASI LBXMPSI LBXHCT LBXSATSI (logged) LBXSASSI (logged) URXUMASI LBXSC3SI URXUCRSI (logged) LBXHGB LBXGH LBDSPHSI LBDTIBSI LBDSCASI LBDFOLSI (logged) LBDSCRSI LBDSALSI LBXRDW LBDSGLSI LBDSUASI LBDB12SI (logged) LBXCOT LBDBANO fs3Score LDLV LBDMONO | SBP average reported to examinee DBP average reported to examinee Self-health index NT-proBNP (pg/ml) (logged) Mean cell hemoglobin (pg) Mean cell volume (fL) Globulin (g/L) Mean Cell Hemoglobin Concentration (g/dL) Protein total (g/L) Transferrin Saturation (%) Lymphocyte percent Ferritin (ug/L) (logged) Platelet count (1000 cells/uL) Lymphocyte number (1000 cells/uL) Iron (umol/L) CRP (mg/dL) (logged) Lactate Dehydrogenase (LDH) (U/L) (logged) Bilirubin total (umol/L) Body Mass Index (kg/m2) (logged) Log Urine Albumin-to-Creatinine Ratio (mg/g) (logged) 60 sec pulse (30 sec pulse X2) Alkaline Phosphatase (ALP) (IU/L) (logged) Segmented neutrophils number (1000 cell/uL) WBC count (1000 cells/uL) (logged) Co-morbidity index Segmented neutrophils percent Monocyte percent Eosinophils percent Potassium (mmol/L) Eosinophils number (1000 cells/uL) Blood Urea Nitrogen (mmol/L) Basophils percent Red blood cell count (million cells/uL) Chloride (mmol/L) Sodium (mmol/L) Mean platelet volume (fL) Hematocrit Alanine Aminotransferase (ALT) (U/L) (logged) Aspartate Aminotransferase (AST) (U/L) (logged) Albumin urine (mg/L) Bicarbonate (mmol/L) Creatinine urine (umol/L) (logged) Hemoglobin (g/dL) Glycohemoglobin (%) Phosphorus (mmol/L) Total iron binding capacity (umol/L) Calcium total (mmol/L) Folate serum (nmol/L) (logged) Creatinine (umol/L) Albumin (g/L) Red cell distribution width (percent) Glucose (mmol/L) Uric acid (umol/L) Vitamin B12 serum (pmol/L) (logged) Cotinine (ng/mL) Basophils number (1000 cells/uL) Healthcare use index Low-Density Lipoprotein (mmol/L) Monocyte number (1000 cells/uL) |
| PC13F | 0.427 0.308 0.284 0.267 0.238 0.184 0.167 0.156 0.154 0.13 0.121 0.092 0.089 0.086 0.082 0.074 0.074 0.067 0.063 0.059 0.05 0.043 0.036 0.034 0.029 0.028 0.028 0.012 0.002 0 -0.273 -0.237 -0.208 -0.134 -0.129 -0.12 -0.119 -0.111 -0.111 -0.108 -0.094 -0.072 -0.065 -0.065 -0.052 -0.052 -0.045 -0.043 -0.043 -0.038 -0.021 -0.019 -0.018 -0.013 -0.011 -0.01 -0.01 -0.008 -0.001 | LBXSC3SI LBXSNASI LBDMONO fs2Score LBXSCLSI LBDLYMNO LBXSATSI (logged) LBXWBCSI (logged) LBXSASSI (logged) SSBNP (logged) LBXMOPCT LBXMCVSI LBXMCHSI LBDNENO URXUCRSI (logged) LBDFERSI (logged) LBXCOT LBXLYPCT LBXSAPSI (logged) LBXSLDSI (logged) LBXMPSI LBXCRP (logged) LBXGH LBXMC URXUMASI BMXBMI (logged) LBDSPHSI LBDB12SI (logged) LBXHGB fs1Score LBDSTBSI fs3Score LBDSTPSI LBDFOLSI (logged) LBDSGBSI LBXEOPCT LBDSALSI BPXDAR LBDSBUSI LBDTIBSI LBDSCASI LBXRBCSI LBDIRNSI LBXNEPCT BPXPLS LBDSCRSI LBDEONO LBXBAPCT LBDSGLSI LBXRDW LBDSUASI crAlbRat (logged) LBXPCT BPXSAR LBXHCT LBXSKSI LDLV LBXPLTSI LBDBANO | Bicarbonate (mmol/L) Sodium (mmol/L) Monocyte number (1000 cells/uL) Self-health index Chloride (mmol/L) Lymphocyte number (1000 cells/uL) Alanine Aminotransferase (ALT) (U/L) (logged) WBC count (1000 cells/uL) (logged) Aspartate Aminotransferase (AST) (U/L) (logged) NT-proBNP (pg/ml) (logged) Monocyte percent Mean cell volume (fL) Mean cell hemoglobin (pg) Segmented neutrophils number (1000 cell/uL) Creatinine urine (umol/L) (logged) Ferritin (ug/L) (logged) Cotinine (ng/mL) Lymphocyte percent Alkaline Phosphatase (ALP) (IU/L) (logged) Lactate Dehydrogenase (LDH) (U/L) (logged) Mean platelet volume (fL) CRP (mg/dL) (logged) Glycohemoglobin (%) Mean Cell Hemoglobin Concentration (g/dL) Albumin urine (mg/L) Body Mass Index (kg/m2) (logged) Phosphorus (mmol/L) Vitamin B12 serum (pmol/L) (logged) Hemoglobin (g/dL) Co-morbidity index Bilirubin total (umol/L) Healthcare use index Protein total (g/L) Folate serum (nmol/L) (logged) Globulin (g/L) Eosinophils percent Albumin (g/L) DBP average reported to examinee Blood Urea Nitrogen (mmol/L) Total iron binding capacity (umol/L) Calcium total (mmol/L) Red blood cell count (million cells/uL) Iron (umol/L) Segmented neutrophils percent 60 sec pulse (30 sec pulse X2) Creatinine (umol/L) Eosinophils number (1000 cells/uL) Basophils percent Glucose (mmol/L) Red cell distribution width (percent) Uric acid (umol/L) Log Urine Albumin-to-Creatinine Ratio (mg/g) (logged) Transferrin Saturation (%) SBP average reported to examinee Hematocrit Potassium (mmol/L) Low-Density Lipoprotein (mmol/L) Platelet count (1000 cells/uL) Basophils number (1000 cells/uL) |
| PC20F | 0.331 0.243 0.233 0.231 0.225 0.18 0.172 0.17 0.165 0.157 0.136 0.105 0.09 0.089 0.088 0.065 0.056 0.054 0.045 0.042 0.03 0.027 0.024 0.021 0.018 0.009 0.008 0.005 0.003 -0.401 -0.212 -0.207 -0.183 -0.174 -0.158 -0.146 -0.139 -0.126 -0.106 -0.097 -0.09 -0.08 -0.07 -0.067 -0.067 -0.066 -0.054 -0.05 -0.047 -0.042 -0.034 -0.026 -0.024 -0.024 -0.014 -0.01 -0.009 -0.005 -0.001 | LBXPLTSI LBXRDW fs2Score LBXSKSI SSBNP (logged) LBXBAPCT LBXPCT LDLV LBDSCRSI LBDIRNSI LBXSAPSI (logged) LBXSLDSI (logged) LBXSASSI (logged) LBXCOT LBXSC3SI LBDSGLSI LBDSPHSI LBXLYPCT LBDSCASI LBDSALSI LBDSTPSI LBDSTBSI LBXGH BPXPLS crAlbRat (logged) LBXMCVSI LBXEOPCT LBDBANO LBDSGBSI LBXMPSI LBDMONO fs3Score LBDFOLSI (logged) LBXMOPCT LBDB12SI (logged) BPXDAR LBXSCLSI BMXBMI (logged) BPXSAR LBXMC LBXCRP (logged) LBDSBUSI LBDFERSI (logged) URXUMASI URXUCRSI (logged) LBXSNASI LBDNENO LBXWBCSI (logged) LBDSUASI LBXSATSI (logged) LBXHGB LBDEONO LBXNEPCT LBXMCHSI LBDTIBSI LBXRBCSI LBDLYMNO LBXHCT fs1Score | Platelet count (1000 cells/uL) Red cell distribution width (percent) Self-health index Potassium (mmol/L) NT-proBNP (pg/ml) (logged) Basophils percent Transferrin Saturation (%) Low-Density Lipoprotein (mmol/L) Creatinine (umol/L) Iron (umol/L) Alkaline Phosphatase (ALP) (IU/L) (logged) Lactate Dehydrogenase (LDH) (U/L) (logged) Aspartate Aminotransferase (AST) (U/L) (logged) Cotinine (ng/mL) Bicarbonate (mmol/L) Glucose (mmol/L) Phosphorus (mmol/L) Lymphocyte percent Calcium total (mmol/L) Albumin (g/L) Protein total (g/L) Bilirubin total (umol/L) Glycohemoglobin (%) 60 sec pulse (30 sec pulse X2) Log Urine Albumin-to-Creatinine Ratio (mg/g) (logged) Mean cell volume (fL) Eosinophils percent Basophils number (1000 cells/uL) Globulin (g/L) Mean platelet volume (fL) Monocyte number (1000 cells/uL) Healthcare use index Folate serum (nmol/L) (logged) Monocyte percent Vitamin B12 serum (pmol/L) (logged) DBP average reported to examinee Chloride (mmol/L) Body Mass Index (kg/m2) (logged) SBP average reported to examinee Mean Cell Hemoglobin Concentration (g/dL) CRP (mg/dL) (logged) Blood Urea Nitrogen (mmol/L) Ferritin (ug/L) (logged) Albumin urine (mg/L) Creatinine urine (umol/L) (logged) Sodium (mmol/L) Segmented neutrophils number (1000 cell/uL) WBC count (1000 cells/uL) (logged) Uric acid (umol/L) Alanine Aminotransferase (ALT) (U/L) (logged) Hemoglobin (g/dL) Eosinophils number (1000 cells/uL) Segmented neutrophils percent Mean cell hemoglobin (pg) Total iron binding capacity (umol/L) Red blood cell count (million cells/uL) Lymphocyte number (1000 cells/uL) Hematocrit Co-morbidity index |
| PC22F | 0.169 0.154 0.14 0.122 0.105 0.105 0.102 0.096 0.09 0.088 0.076 0.066 0.06 0.053 0.053 0.05 0.049 0.048 0.042 0.039 0.039 0.035 0.024 0.015 0.015 0.011 0.011 0.01 0.01 0.002 -0.64 -0.312 -0.241 -0.184 -0.175 -0.166 -0.137 -0.135 -0.122 -0.118 -0.117 -0.11 -0.109 -0.099 -0.092 -0.089 -0.084 -0.076 -0.068 -0.066 -0.065 -0.055 -0.053 -0.039 -0.025 -0.022 -0.016 -0.006 -0.005 | LBXEOPCT LBXSLDSI (logged) LBXSC3SI LBDEONO LBXHGB fs3Score LBXLYPCT LBXMC LBXSATSI (logged) LBXSASSI (logged) LBXHCT LBXGH SSBNP (logged) LBXMCHSI LBDSBUSI BMXBMI (logged) LBXRBCSI crAlbRat (logged) LBDSPHSI LBDLYMNO LBXSKSI LBXRDW LBXMCVSI BPXSAR BPXDAR URXUMASI LBXSAPSI (logged) URXUCRSI (logged) LBXCOT fs1Score LBXBAPCT LBDFOLSI (logged) fs2Score LBXSCLSI LBDMONO LBXSNASI LBDSUASI LBDB12SI (logged) LBXMOPCT LBDSTPSI LBDSGLSI LBXMPSI LBDSTBSI LBDSALSI LBDSCASI LBDIRNSI LBXPCT LBDNENO LBXNEPCT LDLV LBXPLTSI LBXWBCSI (logged) LBDSGBSI BPXPLS LBDFERSI (logged) LBDBANO LBDTIBSI LBDSCRSI LBXCRP (logged) | Eosinophils percent Lactate Dehydrogenase (LDH) (U/L) (logged) Bicarbonate (mmol/L) Eosinophils number (1000 cells/uL) Hemoglobin (g/dL) Healthcare use index Lymphocyte percent Mean Cell Hemoglobin Concentration (g/dL) Alanine Aminotransferase (ALT) (U/L) (logged) Aspartate Aminotransferase (AST) (U/L) (logged) Hematocrit Glycohemoglobin (%) NT-proBNP (pg/ml) (logged) Mean cell hemoglobin (pg) Blood Urea Nitrogen (mmol/L) Body Mass Index (kg/m2) (logged) Red blood cell count (million cells/uL) Log Urine Albumin-to-Creatinine Ratio (mg/g) (logged) Phosphorus (mmol/L) Lymphocyte number (1000 cells/uL) Potassium (mmol/L) Red cell distribution width (percent) Mean cell volume (fL) SBP average reported to examinee DBP average reported to examinee Albumin urine (mg/L) Alkaline Phosphatase (ALP) (IU/L) (logged) Creatinine urine (umol/L) (logged) Cotinine (ng/mL) Co-morbidity index Basophils percent Folate serum (nmol/L) (logged) Self-health index Chloride (mmol/L) Monocyte number (1000 cells/uL) Sodium (mmol/L) Uric acid (umol/L) Vitamin B12 serum (pmol/L) (logged) Monocyte percent Protein total (g/L) Glucose (mmol/L) Mean platelet volume (fL) Bilirubin total (umol/L) Albumin (g/L) Calcium total (mmol/L) Iron (umol/L) Transferrin Saturation (%) Segmented neutrophils number (1000 cell/uL) Segmented neutrophils percent Low-Density Lipoprotein (mmol/L) Platelet count (1000 cells/uL) WBC count (1000 cells/uL) (logged) Globulin (g/L) 60 sec pulse (30 sec pulse X2) Ferritin (ug/L) (logged) Basophils number (1000 cells/uL) Total iron binding capacity (umol/L) Creatinine (umol/L) CRP (mg/dL) (logged) |
| PC23F | 0.346 0.304 0.251 0.219 0.215 0.157 0.147 0.141 0.135 0.122 0.122 0.12 0.11 0.105 0.099 0.097 0.089 0.089 0.087 0.08 0.071 0.064 0.06 0.052 0.047 0.045 0.039 0.033 0.032 0.024 0.013 0.005 0 -0.387 -0.26 -0.186 -0.165 -0.151 -0.118 -0.113 -0.107 -0.106 -0.102 -0.09 -0.087 -0.08 -0.077 -0.07 -0.062 -0.049 -0.04 -0.04 -0.031 -0.025 -0.018 -0.012 -0.01 -0.007 -0.005 | LBDB12SI (logged) LBXSAPSI (logged) LBXSKSI LBDSGBSI LBXCOT LBXRDW LBDFERSI (logged) LBXPCT URXUCRSI (logged) LBXMPSI LBDSTPSI fs3Score LBXCRP (logged) LBXNEPCT LBDNENO BPXDAR LBXMCVSI LBXSC3SI LBXSCLSI LDLV LBDSPHSI LBDIRNSI LBDSGLSI LBXBAPCT LBXPLTSI LBXWBCSI (logged) LBXMCHSI LBXSNASI LBDSTBSI URXUMASI BMXBMI (logged) LBDBANO fs1Score fs2Score LBDSUASI LBDTIBSI LBDFOLSI (logged) LBDSALSI BPXPLS crAlbRat (logged) LBDSCRSI LBXEOPCT LBXRBCSI LBXMC LBXMOPCT LBXLYPCT LBXHGB LBXGH LBXSATSI (logged) LBXHCT LBXSASSI (logged) LBDSCASI LBDEONO LBDSBUSI LBDMONO LBXSLDSI (logged) LBDLYMNO BPXSAR SSBNP (logged) | Vitamin B12 serum (pmol/L) (logged) Alkaline Phosphatase (ALP) (IU/L) (logged) Potassium (mmol/L) Globulin (g/L) Cotinine (ng/mL) Red cell distribution width (percent) Ferritin (ug/L) (logged) Transferrin Saturation (%) Creatinine urine (umol/L) (logged) Mean platelet volume (fL) Protein total (g/L) Healthcare use index CRP (mg/dL) (logged) Segmented neutrophils percent Segmented neutrophils number (1000 cell/uL) DBP average reported to examinee Mean cell volume (fL) Bicarbonate (mmol/L) Chloride (mmol/L) Low-Density Lipoprotein (mmol/L) Phosphorus (mmol/L) Iron (umol/L) Glucose (mmol/L) Basophils percent Platelet count (1000 cells/uL) WBC count (1000 cells/uL) (logged) Mean cell hemoglobin (pg) Sodium (mmol/L) Bilirubin total (umol/L) Albumin urine (mg/L) Body Mass Index (kg/m2) (logged) Basophils number (1000 cells/uL) Co-morbidity index Self-health index Uric acid (umol/L) Total iron binding capacity (umol/L) Folate serum (nmol/L) (logged) Albumin (g/L) 60 sec pulse (30 sec pulse X2) Log Urine Albumin-to-Creatinine Ratio (mg/g) (logged) Creatinine (umol/L) Eosinophils percent Red blood cell count (million cells/uL) Mean Cell Hemoglobin Concentration (g/dL) Monocyte percent Lymphocyte percent Hemoglobin (g/dL) Glycohemoglobin (%) Alanine Aminotransferase (ALT) (U/L) (logged) Hematocrit Aspartate Aminotransferase (AST) (U/L) (logged) Calcium total (mmol/L) Eosinophils number (1000 cells/uL) Blood Urea Nitrogen (mmol/L) Monocyte number (1000 cells/uL) Lactate Dehydrogenase (LDH) (U/L) (logged) Lymphocyte number (1000 cells/uL) SBP average reported to examinee NT-proBNP (pg/ml) (logged) |
| PC24F | 0.455 0.304 0.282 0.271 0.192 0.184 0.168 0.098 0.097 0.091 0.087 0.076 0.074 0.064 0.049 0.039 0.035 0.033 0.029 0.02 0.013 0.011 0.01 0.007 -0.342 -0.286 -0.247 -0.129 -0.119 -0.114 -0.094 -0.094 -0.089 -0.083 -0.082 -0.079 -0.075 -0.073 -0.072 -0.07 -0.058 -0.056 -0.054 -0.053 -0.038 -0.033 -0.031 -0.03 -0.029 -0.028 -0.027 -0.023 -0.021 -0.017 -0.017 -0.009 -0.005 -0.003 -0.002 | LBDSUASI LBXCOT LBXRDW LBDB12SI (logged) LBXPCT crAlbRat (logged) LBDIRNSI LBXMPSI BPXPLS LBDFOLSI (logged) LBDSCRSI LBDSGLSI LBXEOPCT LBDSCASI LBDEONO LBDSALSI BPXDAR LBXMOPCT LBXSNASI LBXSCLSI LBXSKSI LBDSPHSI LBXSC3SI LBXMCVSI LBDSBUSI LBXBAPCT URXUCRSI (logged) LBDSTBSI LBDSGBSI SSBNP (logged) BMXBMI (logged) LBDSTPSI LBXSLDSI (logged) fs3Score LBXMC fs2Score LBXPLTSI URXUMASI LBXSAPSI (logged) BPXSAR LDLV LBXSATSI (logged) LBXSASSI (logged) LBXHGB LBXGH LBXCRP (logged) LBXHCT LBXRBCSI LBDTIBSI LBDNENO LBXWBCSI (logged) LBDFERSI (logged) LBXMCHSI LBDLYMNO LBDMONO LBDBANO LBXLYPCT LBXNEPCT fs1Score | Uric acid (umol/L) Cotinine (ng/mL) Red cell distribution width (percent) Vitamin B12 serum (pmol/L) (logged) Transferrin Saturation (%) Log Urine Albumin-to-Creatinine Ratio (mg/g) (logged) Iron (umol/L) Mean platelet volume (fL) 60 sec pulse (30 sec pulse X2) Folate serum (nmol/L) (logged) Creatinine (umol/L) Glucose (mmol/L) Eosinophils percent Calcium total (mmol/L) Eosinophils number (1000 cells/uL) Albumin (g/L) DBP average reported to examinee Monocyte percent Sodium (mmol/L) Chloride (mmol/L) Potassium (mmol/L) Phosphorus (mmol/L) Bicarbonate (mmol/L) Mean cell volume (fL) Blood Urea Nitrogen (mmol/L) Basophils percent Creatinine urine (umol/L) (logged) Bilirubin total (umol/L) Globulin (g/L) NT-proBNP (pg/ml) (logged) Body Mass Index (kg/m2) (logged) Protein total (g/L) Lactate Dehydrogenase (LDH) (U/L) (logged) Healthcare use index Mean Cell Hemoglobin Concentration (g/dL) Self-health index Platelet count (1000 cells/uL) Albumin urine (mg/L) Alkaline Phosphatase (ALP) (IU/L) (logged) SBP average reported to examinee Low-Density Lipoprotein (mmol/L) Alanine Aminotransferase (ALT) (U/L) (logged) Aspartate Aminotransferase (AST) (U/L) (logged) Hemoglobin (g/dL) Glycohemoglobin (%) CRP (mg/dL) (logged) Hematocrit Red blood cell count (million cells/uL) Total iron binding capacity (umol/L) Segmented neutrophils number (1000 cell/uL) WBC count (1000 cells/uL) (logged) Ferritin (ug/L) (logged) Mean cell hemoglobin (pg) Lymphocyte number (1000 cells/uL) Monocyte number (1000 cells/uL) Basophils number (1000 cells/uL) Lymphocyte percent Segmented neutrophils percent Co-morbidity index |
| PC28F | 0.248 0.239 0.221 0.202 0.195 0.183 0.182 0.18 0.167 0.158 0.155 0.154 0.15 0.127 0.105 0.104 0.078 0.076 0.069 0.054 0.047 0.044 0.042 0.04 0.031 0.021 0.02 0.019 0.018 0.009 0.002 0.002 0.002 -0.502 -0.178 -0.172 -0.169 -0.131 -0.12 -0.115 -0.108 -0.107 -0.107 -0.097 -0.08 -0.08 -0.079 -0.057 -0.054 -0.044 -0.04 -0.036 -0.032 -0.028 -0.025 -0.023 -0.014 -0.009 -0.009 | LBDB12SI (logged) LBDSGLSI LBXCOT LBXSLDSI (logged) SSBNP (logged) LBDSCRSI URXUCRSI (logged) LBXMCVSI LBDTIBSI LBXHGB LBXRDW LBXHCT LBXMCHSI LBXSAPSI (logged) LBXSNASI LBXBAPCT fs3Score BPXDAR BPXPLS LBDSALSI URXUMASI LBXEOPCT LBDSBUSI LBXLYPCT LBXSCLSI LBXRBCSI LBDLYMNO LBDEONO LBDSCASI LBXMC LBDBANO LBXSASSI (logged) fs1Score LBXSKSI LBDFERSI (logged) LBXPCT crAlbRat (logged) LBXGH LBXCRP (logged) LDLV LBDIRNSI BMXBMI (logged) LBXMPSI LBXSATSI (logged) LBXSC3SI LBDSGBSI BPXSAR fs2Score LBDSUASI LBDSTPSI LBXNEPCT LBDSTBSI LBDMONO LBXMOPCT LBDNENO LBDFOLSI (logged) LBDSPHSI LBXWBCSI (logged) LBXPLTSI | Vitamin B12 serum (pmol/L) (logged) Glucose (mmol/L) Cotinine (ng/mL) Lactate Dehydrogenase (LDH) (U/L) (logged) NT-proBNP (pg/ml) (logged) Creatinine (umol/L) Creatinine urine (umol/L) (logged) Mean cell volume (fL) Total iron binding capacity (umol/L) Hemoglobin (g/dL) Red cell distribution width (percent) Hematocrit Mean cell hemoglobin (pg) Alkaline Phosphatase (ALP) (IU/L) (logged) Sodium (mmol/L) Basophils percent Healthcare use index DBP average reported to examinee 60 sec pulse (30 sec pulse X2) Albumin (g/L) Albumin urine (mg/L) Eosinophils percent Blood Urea Nitrogen (mmol/L) Lymphocyte percent Chloride (mmol/L) Red blood cell count (million cells/uL) Lymphocyte number (1000 cells/uL) Eosinophils number (1000 cells/uL) Calcium total (mmol/L) Mean Cell Hemoglobin Concentration (g/dL) Basophils number (1000 cells/uL) Aspartate Aminotransferase (AST) (U/L) (logged) Co-morbidity index Potassium (mmol/L) Ferritin (ug/L) (logged) Transferrin Saturation (%) Log Urine Albumin-to-Creatinine Ratio (mg/g) (logged) Glycohemoglobin (%) CRP (mg/dL) (logged) Low-Density Lipoprotein (mmol/L) Iron (umol/L) Body Mass Index (kg/m2) (logged) Mean platelet volume (fL) Alanine Aminotransferase (ALT) (U/L) (logged) Bicarbonate (mmol/L) Globulin (g/L) SBP average reported to examinee Self-health index Uric acid (umol/L) Protein total (g/L) Segmented neutrophils percent Bilirubin total (umol/L) Monocyte number (1000 cells/uL) Monocyte percent Segmented neutrophils number (1000 cell/uL) Folate serum (nmol/L) (logged) Phosphorus (mmol/L) WBC count (1000 cells/uL) (logged) Platelet count (1000 cells/uL) |
| PC31F | 0.47 0.322 0.243 0.124 0.114 0.105 0.093 0.087 0.086 0.082 0.071 0.067 0.065 0.064 0.064 0.057 0.053 0.053 0.047 0.045 0.018 0.016 0.014 0.012 0.009 0.007 0.005 0.005 0.004 0.004 0.002 -0.47 -0.21 -0.209 -0.18 -0.177 -0.164 -0.161 -0.132 -0.114 -0.086 -0.085 -0.084 -0.081 -0.08 -0.07 -0.07 -0.061 -0.04 -0.034 -0.033 -0.026 -0.026 -0.026 -0.021 -0.011 -0.003 -0.003 -0.001 | LDLV LBDSGLSI LBDFOLSI (logged) LBDLYMNO LBXRDW crAlbRat (logged) LBXSASSI (logged) LBDEONO LBXLYPCT LBDFERSI (logged) LBXEOPCT LBDSBUSI LBXSCLSI LBXCRP (logged) LBDSALSI LBXWBCSI (logged) LBDMONO fs2Score BPXPLS LBDSTPSI LBXMPSI LBXSAPSI (logged) LBDNENO SSBNP (logged) LBXRBCSI LBXPCT LBXMOPCT LBXMCVSI LBXHCT fs3Score LBDSGBSI LBXGH LBDTIBSI LBDSUASI LBDSCASI LBDSPHSI LBXBAPCT LBDB12SI (logged) LBXPLTSI URXUCRSI (logged) LBXMC LBDIRNSI BPXSAR LBXSC3SI LBXNEPCT LBXSNASI LBXSKSI LBDSTBSI BPXDAR URXUMASI LBDSCRSI LBXHGB LBXMCHSI LBXSLDSI (logged) LBXCOT BMXBMI (logged) LBDBANO LBXSATSI (logged) fs1Score | Low-Density Lipoprotein (mmol/L) Glucose (mmol/L) Folate serum (nmol/L) (logged) Lymphocyte number (1000 cells/uL) Red cell distribution width (percent) Log Urine Albumin-to-Creatinine Ratio (mg/g) (logged) Aspartate Aminotransferase (AST) (U/L) (logged) Eosinophils number (1000 cells/uL) Lymphocyte percent Ferritin (ug/L) (logged) Eosinophils percent Blood Urea Nitrogen (mmol/L) Chloride (mmol/L) CRP (mg/dL) (logged) Albumin (g/L) WBC count (1000 cells/uL) (logged) Monocyte number (1000 cells/uL) Self-health index 60 sec pulse (30 sec pulse X2) Protein total (g/L) Mean platelet volume (fL) Alkaline Phosphatase (ALP) (IU/L) (logged) Segmented neutrophils number (1000 cell/uL) NT-proBNP (pg/ml) (logged) Red blood cell count (million cells/uL) Transferrin Saturation (%) Monocyte percent Mean cell volume (fL) Hematocrit Healthcare use index Globulin (g/L) Glycohemoglobin (%) Total iron binding capacity (umol/L) Uric acid (umol/L) Calcium total (mmol/L) Phosphorus (mmol/L) Basophils percent Vitamin B12 serum (pmol/L) (logged) Platelet count (1000 cells/uL) Creatinine urine (umol/L) (logged) Mean Cell Hemoglobin Concentration (g/dL) Iron (umol/L) SBP average reported to examinee Bicarbonate (mmol/L) Segmented neutrophils percent Sodium (mmol/L) Potassium (mmol/L) Bilirubin total (umol/L) DBP average reported to examinee Albumin urine (mg/L) Creatinine (umol/L) Hemoglobin (g/dL) Mean cell hemoglobin (pg) Lactate Dehydrogenase (LDH) (U/L) (logged) Cotinine (ng/mL) Body Mass Index (kg/m2) (logged) Basophils number (1000 cells/uL) Alanine Aminotransferase (ALT) (U/L) (logged) Co-morbidity index |
| PC32F | 0.298 0.294 0.273 0.25 0.236 0.184 0.154 0.106 0.105 0.08 0.076 0.06 0.056 0.049 0.048 0.047 0.044 0.037 0.036 0.031 0.027 0.019 0.018 0.017 0.014 0.013 0.004 0.003 0.001 0 -0.368 -0.304 -0.263 -0.186 -0.173 -0.171 -0.156 -0.14 -0.119 -0.118 -0.113 -0.107 -0.076 -0.072 -0.064 -0.059 -0.058 -0.048 -0.047 -0.046 -0.032 -0.032 -0.025 -0.012 -0.009 -0.008 -0.008 -0.004 -0.001 | LBDFOLSI (logged) LBXSAPSI (logged) LBXCOT LBXRDW LBDSBUSI LBDSPHSI LBDFERSI (logged) BPXSAR BPXPLS LBXSNASI LBXGH LBXPCT LBXBAPCT LBXNEPCT LBXMCHSI LBXMCVSI LBDSCASI crAlbRat (logged) LBXEOPCT LBXCRP (logged) LBXMC fs2Score LBXSATSI (logged) LBDEONO LBXSASSI (logged) LBDSGBSI fs1Score LBXSC3SI LBXHGB LBDBANO LBDB12SI (logged) LDLV SSBNP (logged) LBDTIBSI LBXMPSI LBXSKSI LBDSTBSI LBXSLDSI (logged) LBDSCRSI LBXPLTSI LBDSUASI LBDSGLSI LBDLYMNO fs3Score LBXLYPCT URXUMASI LBDMONO BMXBMI (logged) LBXWBCSI (logged) LBXSCLSI LBXRBCSI LBDSALSI LBDIRNSI LBDNENO LBXHCT BPXDAR LBDSTPSI URXUCRSI (logged) LBXMOPCT | Folate serum (nmol/L) (logged) Alkaline Phosphatase (ALP) (IU/L) (logged) Cotinine (ng/mL) Red cell distribution width (percent) Blood Urea Nitrogen (mmol/L) Phosphorus (mmol/L) Ferritin (ug/L) (logged) SBP average reported to examinee 60 sec pulse (30 sec pulse X2) Sodium (mmol/L) Glycohemoglobin (%) Transferrin Saturation (%) Basophils percent Segmented neutrophils percent Mean cell hemoglobin (pg) Mean cell volume (fL) Calcium total (mmol/L) Log Urine Albumin-to-Creatinine Ratio (mg/g) (logged) Eosinophils percent CRP (mg/dL) (logged) Mean Cell Hemoglobin Concentration (g/dL) Self-health index Alanine Aminotransferase (ALT) (U/L) (logged) Eosinophils number (1000 cells/uL) Aspartate Aminotransferase (AST) (U/L) (logged) Globulin (g/L) Co-morbidity index Bicarbonate (mmol/L) Hemoglobin (g/dL) Basophils number (1000 cells/uL) Vitamin B12 serum (pmol/L) (logged) Low-Density Lipoprotein (mmol/L) NT-proBNP (pg/ml) (logged) Total iron binding capacity (umol/L) Mean platelet volume (fL) Potassium (mmol/L) Bilirubin total (umol/L) Lactate Dehydrogenase (LDH) (U/L) (logged) Creatinine (umol/L) Platelet count (1000 cells/uL) Uric acid (umol/L) Glucose (mmol/L) Lymphocyte number (1000 cells/uL) Healthcare use index Lymphocyte percent Albumin urine (mg/L) Monocyte number (1000 cells/uL) Body Mass Index (kg/m2) (logged) WBC count (1000 cells/uL) (logged) Chloride (mmol/L) Red blood cell count (million cells/uL) Albumin (g/L) Iron (umol/L) Segmented neutrophils number (1000 cell/uL) Hematocrit DBP average reported to examinee Protein total (g/L) Creatinine urine (umol/L) (logged) Monocyte percent |
| PC35F | 0.382 0.372 0.293 0.266 0.155 0.154 0.143 0.127 0.126 0.124 0.119 0.087 0.078 0.075 0.066 0.056 0.048 0.047 0.029 0.022 0.021 0.015 0.013 0.003 0 0 -0.396 -0.212 -0.199 -0.152 -0.133 -0.128 -0.122 -0.112 -0.108 -0.106 -0.091 -0.085 -0.069 -0.062 -0.061 -0.06 -0.05 -0.048 -0.048 -0.043 -0.041 -0.039 -0.028 -0.028 -0.028 -0.022 -0.019 -0.018 -0.014 -0.014 -0.007 -0.005 -0.003 | SSBNP (logged) LDLV LBDSPHSI BPXPLS LBDTIBSI LBXGH LBXCOT BPXDAR LBXSAPSI (logged) LBDSUASI LBDIRNSI LBDSBUSI URXUCRSI (logged) LBDFOLSI (logged) LBXMOPCT LBXPCT URXUMASI LBXCRP (logged) LBDFERSI (logged) LBXEOPCT LBXSLDSI (logged) LBXMPSI LBXSNASI LBDSGBSI LBXNEPCT LBXMC LBDSCRSI LBXRDW BPXSAR LBXSKSI LBXPLTSI BMXBMI (logged) LBDSGLSI LBXMCVSI LBXHGB LBXHCT LBXMCHSI LBXSCLSI LBXBAPCT crAlbRat (logged) LBDLYMNO LBXWBCSI (logged) LBDSTBSI LBDSALSI fs3Score LBDNENO LBXSC3SI LBXSATSI (logged) LBDMONO LBDSTPSI fs2Score LBDEONO LBDSCASI LBXRBCSI LBXLYPCT LBXSASSI (logged) fs1Score LBDB12SI (logged) LBDBANO | NT-proBNP (pg/ml) (logged) Low-Density Lipoprotein (mmol/L) Phosphorus (mmol/L) 60 sec pulse (30 sec pulse X2) Total iron binding capacity (umol/L) Glycohemoglobin (%) Cotinine (ng/mL) DBP average reported to examinee Alkaline Phosphatase (ALP) (IU/L) (logged) Uric acid (umol/L) Iron (umol/L) Blood Urea Nitrogen (mmol/L) Creatinine urine (umol/L) (logged) Folate serum (nmol/L) (logged) Monocyte percent Transferrin Saturation (%) Albumin urine (mg/L) CRP (mg/dL) (logged) Ferritin (ug/L) (logged) Eosinophils percent Lactate Dehydrogenase (LDH) (U/L) (logged) Mean platelet volume (fL) Sodium (mmol/L) Globulin (g/L) Segmented neutrophils percent Mean Cell Hemoglobin Concentration (g/dL) Creatinine (umol/L) Red cell distribution width (percent) SBP average reported to examinee Potassium (mmol/L) Platelet count (1000 cells/uL) Body Mass Index (kg/m2) (logged) Glucose (mmol/L) Mean cell volume (fL) Hemoglobin (g/dL) Hematocrit Mean cell hemoglobin (pg) Chloride (mmol/L) Basophils percent Log Urine Albumin-to-Creatinine Ratio (mg/g) (logged) Lymphocyte number (1000 cells/uL) WBC count (1000 cells/uL) (logged) Bilirubin total (umol/L) Albumin (g/L) Healthcare use index Segmented neutrophils number (1000 cell/uL) Bicarbonate (mmol/L) Alanine Aminotransferase (ALT) (U/L) (logged) Monocyte number (1000 cells/uL) Protein total (g/L) Self-health index Eosinophils number (1000 cells/uL) Calcium total (mmol/L) Red blood cell count (million cells/uL) Lymphocyte percent Aspartate Aminotransferase (AST) (U/L) (logged) Co-morbidity index Vitamin B12 serum (pmol/L) (logged) Basophils number (1000 cells/uL) |
| PC37F | 0.367 0.209 0.184 0.141 0.129 0.117 0.09 0.076 0.071 0.064 0.063 0.058 0.045 0.044 0.032 0.023 0.021 0.017 0.01 0.01 0.009 0.008 0.005 0.005 0.004 0.003 0.002 0.002 -0.504 -0.501 -0.214 -0.141 -0.135 -0.131 -0.129 -0.129 -0.113 -0.108 -0.093 -0.08 -0.076 -0.065 -0.051 -0.047 -0.047 -0.044 -0.032 -0.026 -0.019 -0.019 -0.018 -0.009 -0.009 -0.007 -0.005 -0.004 -0.003 -0.003 -0.001 | LBDSPHSI LBXMCVSI LBDSUASI LBXSLDSI (logged) LBXHCT LBDSTBSI LBXPLTSI LBDFERSI (logged) LBXSKSI LBDSGLSI BPXSAR LBXCOT LBDSBUSI crAlbRat (logged) LBXBAPCT BPXPLS LBXSATSI (logged) LBDLYMNO LBXMOPCT LBXMPSI LBXLYPCT fs2Score LBDNENO LBDSTPSI LBDMONO LBDSGBSI LBDBANO LBDSALSI LBDSCASI LBXMC LBXRDW LBXCRP (logged) LBXPCT SSBNP (logged) LBDIRNSI LDLV BMXBMI (logged) URXUCRSI (logged) LBXGH LBDFOLSI (logged) LBDSCRSI LBXSC3SI BPXDAR LBXEOPCT LBXSNASI URXUMASI LBDB12SI (logged) LBXHGB LBXRBCSI LBXSAPSI (logged) LBXWBCSI (logged) LBDEONO LBXMCHSI fs3Score fs1Score LBXSCLSI LBDTIBSI LBXSASSI (logged) LBXNEPCT | Phosphorus (mmol/L) Mean cell volume (fL) Uric acid (umol/L) Lactate Dehydrogenase (LDH) (U/L) (logged) Hematocrit Bilirubin total (umol/L) Platelet count (1000 cells/uL) Ferritin (ug/L) (logged) Potassium (mmol/L) Glucose (mmol/L) SBP average reported to examinee Cotinine (ng/mL) Blood Urea Nitrogen (mmol/L) Log Urine Albumin-to-Creatinine Ratio (mg/g) (logged) Basophils percent 60 sec pulse (30 sec pulse X2) Alanine Aminotransferase (ALT) (U/L) (logged) Lymphocyte number (1000 cells/uL) Monocyte percent Mean platelet volume (fL) Lymphocyte percent Self-health index Segmented neutrophils number (1000 cell/uL) Protein total (g/L) Monocyte number (1000 cells/uL) Globulin (g/L) Basophils number (1000 cells/uL) Albumin (g/L) Calcium total (mmol/L) Mean Cell Hemoglobin Concentration (g/dL) Red cell distribution width (percent) CRP (mg/dL) (logged) Transferrin Saturation (%) NT-proBNP (pg/ml) (logged) Iron (umol/L) Low-Density Lipoprotein (mmol/L) Body Mass Index (kg/m2) (logged) Creatinine urine (umol/L) (logged) Glycohemoglobin (%) Folate serum (nmol/L) (logged) Creatinine (umol/L) Bicarbonate (mmol/L) DBP average reported to examinee Eosinophils percent Sodium (mmol/L) Albumin urine (mg/L) Vitamin B12 serum (pmol/L) (logged) Hemoglobin (g/dL) Red blood cell count (million cells/uL) Alkaline Phosphatase (ALP) (IU/L) (logged) WBC count (1000 cells/uL) (logged) Eosinophils number (1000 cells/uL) Mean cell hemoglobin (pg) Healthcare use index Co-morbidity index Chloride (mmol/L) Total iron binding capacity (umol/L) Aspartate Aminotransferase (AST) (U/L) (logged) Segmented neutrophils percent |
| PC38F | 0.47 0.279 0.195 0.132 0.128 0.11 0.099 0.092 0.075 0.073 0.07 0.063 0.063 0.057 0.052 0.051 0.048 0.046 0.043 0.042 0.041 0.036 0.033 0.028 0.01 0.008 0.007 0.007 0.005 0.003 -0.329 -0.266 -0.234 -0.234 -0.184 -0.181 -0.18 -0.17 -0.149 -0.14 -0.137 -0.122 -0.102 -0.101 -0.091 -0.089 -0.078 -0.068 -0.065 -0.057 -0.04 -0.037 -0.032 -0.032 -0.025 -0.023 -0.012 -0.002 -0.001 | LBXCOT LBXSLDSI (logged) LBXSKSI LBXMC LBDSGBSI LBDFERSI (logged) LBDSUASI LBXRBCSI LBXSCLSI LBDNENO LBDSBUSI LBXBAPCT LBDSTPSI URXUCRSI (logged) LBXSASSI (logged) BPXDAR LBXWBCSI (logged) LBDSGLSI fs2Score LBDFOLSI (logged) LBDLYMNO LBXNEPCT LBXSC3SI LBDTIBSI LBXHGB URXUMASI LBDEONO LBDBANO LBDSTBSI LBDMONO LBDSPHSI LBXPLTSI LBXMPSI LBXSAPSI (logged) BPXPLS LBXCRP (logged) LBXMCVSI BMXBMI (logged) LBDSCRSI LBXSNASI LBXRDW LBXSATSI (logged) LBXMCHSI LBDSALSI LBXEOPCT LDLV LBXPCT fs3Score LBDIRNSI LBDSCASI LBXMOPCT crAlbRat (logged) LBXHCT SSBNP (logged) BPXSAR LBDB12SI (logged) LBXLYPCT LBXGH fs1Score | Cotinine (ng/mL) Lactate Dehydrogenase (LDH) (U/L) (logged) Potassium (mmol/L) Mean Cell Hemoglobin Concentration (g/dL) Globulin (g/L) Ferritin (ug/L) (logged) Uric acid (umol/L) Red blood cell count (million cells/uL) Chloride (mmol/L) Segmented neutrophils number (1000 cell/uL) Blood Urea Nitrogen (mmol/L) Basophils percent Protein total (g/L) Creatinine urine (umol/L) (logged) Aspartate Aminotransferase (AST) (U/L) (logged) DBP average reported to examinee WBC count (1000 cells/uL) (logged) Glucose (mmol/L) Self-health index Folate serum (nmol/L) (logged) Lymphocyte number (1000 cells/uL) Segmented neutrophils percent Bicarbonate (mmol/L) Total iron binding capacity (umol/L) Hemoglobin (g/dL) Albumin urine (mg/L) Eosinophils number (1000 cells/uL) Basophils number (1000 cells/uL) Bilirubin total (umol/L) Monocyte number (1000 cells/uL) Phosphorus (mmol/L) Platelet count (1000 cells/uL) Mean platelet volume (fL) Alkaline Phosphatase (ALP) (IU/L) (logged) 60 sec pulse (30 sec pulse X2) CRP (mg/dL) (logged) Mean cell volume (fL) Body Mass Index (kg/m2) (logged) Creatinine (umol/L) Sodium (mmol/L) Red cell distribution width (percent) Alanine Aminotransferase (ALT) (U/L) (logged) Mean cell hemoglobin (pg) Albumin (g/L) Eosinophils percent Low-Density Lipoprotein (mmol/L) Transferrin Saturation (%) Healthcare use index Iron (umol/L) Calcium total (mmol/L) Monocyte percent Log Urine Albumin-to-Creatinine Ratio (mg/g) (logged) Hematocrit NT-proBNP (pg/ml) (logged) SBP average reported to examinee Vitamin B12 serum (pmol/L) (logged) Lymphocyte percent Glycohemoglobin (%) Co-morbidity index |
| PC39F | 0.354 0.232 0.23 0.221 0.202 0.162 0.108 0.093 0.081 0.08 0.076 0.072 0.066 0.064 0.061 0.059 0.051 0.032 0.03 0.03 0.03 0.023 0.023 0.019 0.016 0.015 0.014 0.01 0.009 0.007 0.002 0.001 0 -0.49 -0.454 -0.189 -0.124 -0.119 -0.109 -0.109 -0.106 -0.086 -0.08 -0.063 -0.058 -0.056 -0.055 -0.047 -0.038 -0.037 -0.036 -0.036 -0.035 -0.031 -0.021 -0.008 -0.008 -0.007 -0.003 | LBXCRP (logged) LBXMCVSI LBDFERSI (logged) LBDTIBSI LBDSALSI LBDSCASI LBXCOT LBDSTBSI BMXBMI (logged) LBXGH LBDSBUSI LBXSAPSI (logged) LBXSKSI SSBNP (logged) BPXPLS fs2Score URXUCRSI (logged) LBXMCHSI URXUMASI LBXEOPCT LBXHCT BPXDAR LBXPLTSI LBDFOLSI (logged) LBXSLDSI (logged) LBXSATSI (logged) LBXSC3SI fs1Score LBXLYPCT LBXMPSI LBDIRNSI LBDEONO LBXMOPCT LBDSPHSI LBXMC LBDSGBSI LBXRBCSI LBDSCRSI LBXHGB LBDSGLSI LBXPCT LDLV LBXBAPCT fs3Score LBDSTPSI BPXSAR LBDMONO LBDNENO LBDSUASI LBXSCLSI LBXSASSI (logged) LBXSNASI LBXWBCSI (logged) crAlbRat (logged) LBDLYMNO LBDB12SI (logged) LBXNEPCT LBXRDW LBDBANO | CRP (mg/dL) (logged) Mean cell volume (fL) Ferritin (ug/L) (logged) Total iron binding capacity (umol/L) Albumin (g/L) Calcium total (mmol/L) Cotinine (ng/mL) Bilirubin total (umol/L) Body Mass Index (kg/m2) (logged) Glycohemoglobin (%) Blood Urea Nitrogen (mmol/L) Alkaline Phosphatase (ALP) (IU/L) (logged) Potassium (mmol/L) NT-proBNP (pg/ml) (logged) 60 sec pulse (30 sec pulse X2) Self-health index Creatinine urine (umol/L) (logged) Mean cell hemoglobin (pg) Albumin urine (mg/L) Eosinophils percent Hematocrit DBP average reported to examinee Platelet count (1000 cells/uL) Folate serum (nmol/L) (logged) Lactate Dehydrogenase (LDH) (U/L) (logged) Alanine Aminotransferase (ALT) (U/L) (logged) Bicarbonate (mmol/L) Co-morbidity index Lymphocyte percent Mean platelet volume (fL) Iron (umol/L) Eosinophils number (1000 cells/uL) Monocyte percent Phosphorus (mmol/L) Mean Cell Hemoglobin Concentration (g/dL) Globulin (g/L) Red blood cell count (million cells/uL) Creatinine (umol/L) Hemoglobin (g/dL) Glucose (mmol/L) Transferrin Saturation (%) Low-Density Lipoprotein (mmol/L) Basophils percent Healthcare use index Protein total (g/L) SBP average reported to examinee Monocyte number (1000 cells/uL) Segmented neutrophils number (1000 cell/uL) Uric acid (umol/L) Chloride (mmol/L) Aspartate Aminotransferase (AST) (U/L) (logged) Sodium (mmol/L) WBC count (1000 cells/uL) (logged) Log Urine Albumin-to-Creatinine Ratio (mg/g) (logged) Lymphocyte number (1000 cells/uL) Vitamin B12 serum (pmol/L) (logged) Segmented neutrophils percent Red cell distribution width (percent) Basophils number (1000 cells/uL) |

| **Supplementary Table 4. PC weights in LinAge2.** | | | | | |
| --- | --- | --- | --- | --- | --- |
| **PC (Male)** | **Weights** | **P-Value** | **PC (Female)** | **Weights** | **P-Value** |
| Chronological age | 0.0054220 | < 2e-16 | Chronological age | 0.0074772 | < 2e-16 |
| PC1M | 0.1798438 | < 2e-16 | PC1F | 0.2607816 | < 2e-16 |
| PC2M | 0.0341744 | 0.047737 | PC2F | 0.0551195 | 0.030598 |
| PC5M | 0.1333414 | 1.76e-10 | PC4F | 0.0656256 | 0.008313 |
| PC6M | 0.0693078 | 0.001074 | PC6F | 0.0805784 | 0.002204 |
| PC8M | 0.0492219 | 0.046424 | PC11F | 0.0788920 | 0.010210 |
| PC11M | 0.1156164 | 5.58e-05 | PC13F | 0.1761816 | 5.30e-07 |
| PC15M | 0.0681599 | 0.026055 | PC20F | 0.1916443 | 8.17e-06 |
| PC16M | 0.0739918 | 0.020443 | PC22F | 0.1376097 | 0.002230 |
| PC17M | 0.1031597 | 0.000752 | PC23F | 0.1006660 | 0.022460 |
| PC19M | 0.0778960 | 0.018922 | PC24F | 0.1192029 | 0.010945 |
| PC24M | 0.1012453 | 0.003816 | PC28F | 0.1469867 | 0.004360 |
| PC25M | 0.0769868 | 0.044519 | PC31F | 0.1109110 | 0.038749 |
| PC27M | 0.1032640 | 0.012580 | PC32F | 0.1897054 | 0.000791 |
| PC31M | 0.1326559 | 0.003628 | PC35F | 0.2200549 | 0.000213 |
| PC33M | 0.1198606 | 0.008850 | PC37F | 0.1413076 | 0.035837 |
| PC36M | 0.1011755 | 0.046204 | PC38F | 0.1253549 | 0.052444 |
| PC42M | 0.1711699 | 0.005489 | PC39F | 0.1463831 | 0.024302 |

| **Supplementary Table 5. PC interpretation in LinAge2.** | | | |  |
| --- | --- | --- | --- | --- |
| **PC (Male)** | **Causes of Death** | **Diseases / Lifestyle / Social Factors** | **Mechanisms** | **Interventions / Management** |
| PC1M | Early (within 0-5 years)   - Diabetes mellitus - Cardiovascular disease - Stroke - Chronic lung disease - Chronic kidney disease - Cancer - Alzheimer’s disease - Others   Late (within 10-20 years)   - Pneumonia - Diabetes mellitus - Cardiovascular disease - Stroke - Chronic lung disease - Chronic kidney disease - Cancer - Alzheimer’s disease - Others | Cardiometabolic syndrome   - Obesity - Hypertension - Hypercholesterolemia - Diabetes mellitus and insulin use - Diabetic complications - Cardiovascular disease, including congestive cardiac failure - Stroke   Organ impairment   - Cognitive impairment - Visual impairment - Thyroid disease - Chronic lung diseases (asthma, chronic bronchitis, emphysema) - Chronic kidney disease - Chronic liver disease - Arthritis - Osteoporosis - Anemia   Cancer  Lifestyle and Social factors   - Do not exercise (low vigorous activity, low moderate activity, and do less muscle strengthening) - Alcohol use - Low education - Low income | - Vascular aging - Metabolic aging - Inflammation - Neurodegeneration | - Screen for and consider appropriate management of cardiometabolic syndrome, organ impairment(s), and cancer - Screen for and consider appropriate management of cognitive impairment (including thyroid disease) - Manage obesity - Increase exercise - Reduce alcohol use - Consider age-appropriate vaccinations |
| PC2M | Early (within 0-5 years)   - Diabetes mellitus - Cardiovascular disease - Cancer - Others   Late (within 10-20 years)   - Pneumonia - Cardiovascular disease - Chronic lung disease - Cancer - Alzheimer’s disease - Others | Cardiovascular disease   - Cardiovascular disease, including congestive cardiac failure   Organ impairment   - Cognitive impairment - Thyroid disease - Emphysema - Chronic kidney disease - Arthritis - Osteoporosis - Anemia   Cancer  Lifestyle and Social factors   - Thin - High education - High income | - Vascular aging - Inflammation - Neurodegeneration | - Screen for and consider appropriate management of cardiovascular disease, organ impairment(s), and cancer - Screen for and consider appropriate management of cognitive impairment (including thyroid disease) - Consider age-appropriate vaccinations |
| PC5M | Early (within 0-5 years)   - Cardiovascular disease - Chronic lung disease - Chronic kidney disease - Cancer - Others   Late (within 10-20 years)   - Cardiovascular disease - Chronic lung disease - Cancer - Alzheimer’s disease - Others | Cardiovascular disease   - Hypertension - Diabetic peripheral neuropathy - Cardiovascular disease, including congestive cardiac failure - Stroke   Organ impairment   - Cognitive impairment - Visual impairment - Chronic lung diseases (asthma, chronic bronchitis, emphysema) - Arthritis - Spine fracture   Lifestyle and Social factors   - Thin - Cigarette smoking - Alcohol use - Do not exercise (low vigorous activity and do less muscle strengthening) - Low income | - Vascular aging (smoking-related) - Lung disease-related - Inflammation - Neurodegeneration | - Screen for and consider appropriate management of cardiovascular disease, vascular risk factors, organ impairment(s), and cancer - Screen for and consider appropriate management of cognitive impairment - Quit smoking - Reduce alcohol use - Increase exercise - Consider age-appropriate vaccinations |
| PC6M | Early (within 0-5 years)   - Others | Vascular risk factors   - Hypertension - Diabetic peripheral neuropathy - Peripheral arterial disease - Diabetic foot ulcers   Organ impairment   - Cognitive impairment - Thyroid disease - Asthma - Chronic liver disease - Chronic kidney disease - Anemia   Lifestyle and Social factors   - Cigarette smoking - Alcohol use - Do not exercise (low vigorous activity, low moderate activity, and do less muscle strengthening) - Low income | - Vascular aging (smoking-related) | - Screen for and consider appropriate management of vascular risk factors, especially diabetic complications of the lower limb - Screen for and consider appropriate management of organ impairment(s) - Quit smoking - Reduce alcohol use - Increase exercise |
| PC8M | Early (within 0-5 years)   - Others   Late (within 10-20 years)   - Others | Vascular risk factors   - Hypertension - Hypercholesterolemia   Organ impairment   - Chronic liver disease - Chronic kidney disease - Anemia - Wrist fracture - Visual impairment   Lifestyle and Social factors   - Obesity - Do not exercise (low moderate activity) | - Vascular aging - Inflammation | - Screen for and consider appropriate management of vascular risk factors - Screen for and consider appropriate management of organ impairment(s) - Manage obesity - Increase exercise |
| PC11M | Early (within 0-5 years)   - Stroke - Others | Organ impairment   - Emphysema - Visual impairment   Lifestyle and Social factors   - Thin - Alcohol use - Do not exercise (low vigorous and moderate activity) - Low education - Low income | - Indeterminate | - Screen for and consider appropriate management of organ impairment(s) - Reduce alcohol use - Increase exercise |
| PC15M | Early (within 0-5 years)   - Diabetes mellitus | Diabetic complications   - Diabetic peripheral neuropathy   Organ impairment   - Cognitive impairment - Visual impairment - Chronic bronchitis - Emphysema   Lifestyle and Social factors   - Thin - Alcohol use - Do not exercise (low vigorous activity, low moderate activity, and do less muscle strengthening) - Low education - Low income | - Inflammation | - Screen for and consider appropriate management of diabetes and its complications - Screen for and consider appropriate management of organ impairment(s) - Reduce alcohol use - Increase exercise |
| PC16M | Early (within 0-5 years)   - Stroke - Chronic lung disease - Chronic kidney disease - Cancer - Others   Late (within 10-20 years)   - Pneumonia - Diabetes mellitus - Cardiovascular disease - Chronic kidney disease - Cancer - Others | Lifestyle and Social factors   - Thin - Cigarette smoking - Alcohol use - Do not exercise (low vigorous activity and sedentary) - Low income | - Smoking-related | - Screen for and consider appropriate management of smoking-related diseases (if smoker) - Quit smoking - Reduce alcohol use - Increase exercise - Consider age-appropriate vaccinations |
| PC17M | Late (within 10-20 years)   - Diabetes mellitus | Vascular risk factors   - Hypertension   Lifestyle and Social factors   - Do not exercise (less muscle strengthening) | - Indeterminate | - Screen for and consider appropriate management of vascular risk factors - Increase exercise |
| PC19M | Early (within 0-5 years)   - Diabetes mellitus - Stroke - Others   Late (within 10-20 years)   - Alzheimer’s disease - Cancer | Cardiovascular disease   - Diabetes mellitus - Ischemic heart disease   Organ impairment   - Thyroid disease   Lifestyle and Social factors   - Thin - Do not exercise (low vigorous activity) - Low education - Low income | - Vascular aging - Neurodegeneration | - Screen for and consider appropriate management of cardiovascular disease and vascular risk factors - Screen for and consider appropriate management of cognitive impairment (including thyroid disease) - Screen for and consider appropriate management of cancer - Increase exercise |
| PC24M | - None | Lifestyle and Social factors   - Cigarette smoking | - Indeterminate | - Quit smoking |
| PC25M | Early (within 0-5 years)   - Cancer   Late (within 10-20 years)   - Others | Cardiovascular disease   - Hypertension - Hyperlipidemia - Cardiovascular disease, including congestive cardiac failure - Stroke   Lifestyle and Social factors   - Obesity - Low education | - Vascular aging - Inflammation | - Screen for and consider appropriate management of cardiovascular disease, vascular risk factors, and cancer - Manage obesity |
| PC27M | Late (within 10-20 years)   - Chronic lung disease - Cancer | Lifestyle and Social factors   - Do not exercise (low moderate activity and sedentary) | - Indeterminate | - Screen for and consider appropriate management of chronic lung disease and cancer - Increase exercise - Consider age-appropriate vaccinations |
| PC31M | Early (within 0-5 years)   - Cardiovascular disease   Late (within 10-20 years)   - Cardiovascular disease | Cardiovascular disease   - Diabetes mellitus - Cardiovascular disease, including congestive cardiac failure   Organ impairment   - Visual impairment   Cancer  Lifestyle and Social factors   - Obesity - Cigarette smoking - Do not exercise (low vigorous activity) - Low income | - Vascular aging (smoking-related) - Cardiac disease-related | - Screen for and consider appropriate management of diabetes and cardiovascular disease - Screen for and consider appropriate management of visual impairment and cancer - Manage obesity - Quit smoking - Increase exercise |
| PC33M | - None | Cardiovascular disease   - Angina   Lifestyle and Social factors   - High education | - Inflammation | - Screen for and consider appropriate management of cardiovascular disease |
| PC36M | Early (within 0-5 years)   - Diabetes mellitus | Vascular risk factors   - Diabetes mellitus   Lifestyle and Social factors   - Cigarette smoking | - Indeterminate | - Screen for and consider appropriate management of diabetes - Quit smoking |
| PC42M | - None | Vascular disease   - Peripheral arterial disease   Organ impairment   - Thyroid disease   Lifestyle and Social factors   - Do not exercise (low vigorous and moderate activity) | - Indeterminate | - Screen for and consider appropriate management of peripheral arterial disease and thyroid disease - Increase exercise |
| **PC (Female)** | **Causes of Death** | **Diseases / Lifestyle / Social Factors** | **Mechanisms** | **Interventions / Management** |
| PC1F | Early (within 0-5 years)   - Pneumonia - Cardiovascular disease - Stroke - Chronic lung disease - Cancer - Others   Late (within 10-20 years)   - Pneumonia - Diabetes mellitus - Cardiovascular disease - Stroke - Chronic lung disease - Chronic kidney disease - Cancer - Alzheimer’s disease - Others | Cardiometabolic syndrome   - Obesity - Hypertension - Hypercholesterolemia - Diabetes mellitus and insulin use - Diabetic complications - Cardiovascular disease, including congestive cardiac failure - Stroke   Organ impairment   - Cognitive impairment - Visual impairment - Chronic lung diseases (asthma, chronic bronchitis, emphysema) - Chronic kidney disease - Chronic liver disease - Arthritis - Osteoporosis - Anemia   Cancer  Falls  Lifestyle and Social factors   - Do not exercise (low vigorous activity, low moderate activity, and do less muscle strengthening) - Alcohol use - Low education - Low income | - Vascular aging - Metabolic aging - Inflammation - Neurodegeneration | - Screen for and consider appropriate management of cardiometabolic syndrome, organ impairment(s), and cancer - Screen for and consider appropriate management of geriatric syndromes (cognitive impairment, falls, osteoporosis) - Manage obesity - Increase exercise - Reduce alcohol use - Consider age-appropriate vaccinations |
| PC2F | Early (within 0-5 years)   - Cardiovascular disease - Stroke - Chronic lung disease - Cancer - Others   Late (within 10-20 years)   - Pneumonia - Diabetes mellitus - Cardiovascular disease - Stroke - Chronic lung disease - Chronic kidney disease - Cancer - Alzheimer’s disease - Others | Cardiovascular disease   - Hypertension - Hypercholesterolemia - Diabetic foot ulcers - Diabetic retinopathy - Cardiovascular disease, including congestive cardiac failure - Stroke   Organ impairment   - Cognitive impairment - Thyroid disease - Chronic bronchitis - Emphysema - Chronic liver disease - Arthritis - Osteoporosis - Hip fracture - Spine fracture   Cancer  Falls  Lifestyle and Social factors   - Thin - Do not exercise (sedentary) - High education - High income | - Vascular aging - Neurodegeneration | - Screen for and consider appropriate management of cardiovascular disease, vascular risk factors, organ impairment(s), and cancer - Screen for and consider appropriate management of geriatric syndromes (cognitive impairment, falls, osteoporosis) - Increase exercise - Consider age-appropriate vaccinations |
| PC4F | Early (within 0-5 years)   - Chronic lung disease   Late (within 10-20 years)   - Cardiovascular disease - Chronic kidney disease | Cardiovascular disease   - Hypertension - Diabetes mellitus and insulin use - Diabetic peripheral neuropathy - Cardiovascular disease, including congestive cardiac failure - Stroke   Organ impairment   - Cognitive impairment - Visual impairment - Asthma - Chronic bronchitis - Arthritis - Osteoporosis - Hip fracture - Anemia   Cancer  Falls  Lifestyle and Social factors   - Obesity - Do not exercise (low vigorous and moderate activity) - Low income | - Vascular aging - Inflammation | - Screen for and consider appropriate management of cardiovascular disease, vascular risk factors, organ impairment(s), and cancer - Screen for and consider appropriate management of geriatric syndromes (cognitive impairment, falls, osteoporosis) - Manage obesity - Increase exercise - Consider age-appropriate vaccinations |
| PC6F | Early (within 0-5 years)   - Pneumonia - Cardiovascular disease - Chronic lung disease - Cancer - Others   Late (within 10-20 years)   - Cardiovascular disease - Chronic lung disease - Cancer - Others | Cardiovascular disease   - Hypertension - Hypercholesterolemia - Diabetic peripheral neuropathy - Cardiovascular disease, including congestive cardiac failure   Organ impairment   - Thyroid disease - Chronic bronchitis - Chronic kidney disease - Arthritis - Osteoporosis - Hip fracture   Lifestyle and Social factors   - Obesity - Cigarette smoking - Alcohol use - Do not exercise (low vigorous activity, low moderate activity, and do less muscle strengthening) | - Vascular aging (smoking-related) - Inflammation | - Screen for and consider appropriate management of cardiovascular disease, vascular risk factors, and organ impairment(s) - Manage obesity - Quit smoking - Reduce alcohol use - Increase exercise - Consider age-appropriate vaccinations |
| PC11F | Early (within 0-5 years)   - Cardiovascular disease   Late (within 10-20 years)   - Others | Cardiovascular disease   - Hypertension - Diabetic retinopathy - Diabetic peripheral neuropathy - Peripheral arterial disease - Ischemic heart disease - Congestive cardiac failure - Stroke   Organ impairment   - Cognitive impairment - Arthritis   Lifestyle and Social factors   - Alcohol use - Do not exercise (low vigorous and moderate activity) - Low education - Low income | - Vascular aging | - Screen for and consider appropriate management of cardiovascular disease, vascular risk factors, diabetic complications, cognitive impairment, and arthritis - Reduce alcohol use - Increase exercise |
| PC13F | Early (within 0-5 years)   - Cardiovascular disease - Stroke - Chronic lung disease   Late (within 10-20 years)   - Cardiovascular disease - Cancer | Cardiovascular disease   - Diabetic peripheral neuropathy - Angina - Acute myocardial infarction - Stroke   Organ impairment   - Cognitive impairment - Visual impairment - Emphysema   Lifestyle and Social factors   - Obesity - Alcohol use - Do not exercise (low vigorous activity, low moderate activity, and do less muscle strengthening) - Low education - Low income | - Vascular aging | - Screen for and consider appropriate management of cardiovascular disease, vascular risk factors, cognitive impairment, visual impairment, chronic lung disease, and cancer - Manage obesity - Reduce alcohol use - Increase exercise - Consider age-appropriate vaccinations |
| PC20F | Early (within 0-5 years)   - Pneumonia - Cardiovascular disease - Others   Late (within 10-20 years)   - Chronic lung disease - Others | Cardiovascular disease   - Hypercholesterolemia - Angina - Congestive cardiac failure - Stroke   Organ impairment   - Visual impairment - Emphysema - Spine fracture - Anemia   Lifestyle and Social factors   - Thin - Do not exercise (low vigorous activity, low moderate activity, and do less muscle strengthening) - Low income | - Vascular aging | - Screen for and consider appropriate management of cardiovascular disease, vascular risk factors, and organ impairment(s) especially chronic lung disease - Increase exercise - Consider age-appropriate vaccinations |
| PC22F | Early (within 0-5 years)   - Others   Late (within 10-20 years)   - Chronic kidney disease - Others | Cardiovascular disease   - Hypertension - Insulin use - Congestive cardiac failure   Organ impairment   - Asthma   Lifestyle and Social factors   - Obesity | - Indeterminate | - Screen for and consider appropriate management of vascular risk factors, congestive cardiac failure, chronic kidney disease, and asthma - Manage obesity |
| PC23F | Early (within 0-5 years)   - Cardiovascular disease - Others   Late (within 10-20 years)   - Pneumonia - Chronic lung disease - Chronic kidney disease - Cancer | Vascular risk factors   - Diabetic foot ulcers   Organ impairment   - Anemia   Lifestyle and Social factors   - Thin - Cigarette smoking - Alcohol use | - Inflammation | - Screen for and consider appropriate management of vascular risk factors, diabetic complications, and anemia - Quit smoking - Reduce alcohol use |
| PC24F | Late (within 10-20 years)   - Chronic lung disease - Cancer - Alzheimer’s disease - Others | Lifestyle and Social factors   - Thin - Cigarette smoking - Alcohol use | - Indeterminate | - Quit smoking - Reduce alcohol use |
| PC28F | Early (within 0-5 years)   - Cardiovascular disease - Cancer   Late (within 10-20 years)   - Others | Cardiovascular disease   - Diabetic retinopathy - Diabetic peripheral neuropathy - Acute myocardial infarction   Lifestyle and Social factors   - Thin - High income | - Indeterminate | - Screen for and consider appropriate management of cardiovascular disease, vascular risk factors, diabetic complications, and cancer |
| PC31F | Early (within 0-5 years)   - Cardiovascular disease - Chronic lung disease   Late (within 10-20 years)   - Chronic lung disease - Chronic kidney disease - Cancer - Others | Vascular risk factors   - Hypercholesterolemia   Organ impairment   - Cognitive impairment - Emphysema - Arthritis - Hip fracture - Anemia   Lifestyle and Social factors   - Thin - Low income | - Indeterminate | - Screen for and consider appropriate management of vascular risk factors and organ impairment(s) especially chronic lung disease - Consider age-appropriate vaccinations |
| PC32F | Early (within 0-5 years)   - Stroke - Others   Late (within 10-20 years)   - Diabetes mellitus - Cancer - Others | Vascular risk factors   - Hypertension   Organ impairment   - Chronic bronchitis - Chronic kidney disease - Arthritis   Lifestyle and Social factors   - Cigarette smoking - Alcohol use - Do not exercise (low moderate activity and do less muscle strengthening) - Low education | - Indeterminate | - Screen for and consider appropriate management of vascular risk factors and organ impairment(s) - Quit smoking - Reduce alcohol use - Increase exercise |
| PC35F | Early (within 0-5 years)   - Cardiovascular disease - Chronic lung disease - Cancer - Others   Late (within 10-20 years)   - Pneumonia - Cardiovascular disease - Chronic lung disease - Alzheimer’s disease | Cardiovascular disease   - Hypercholesterolemia - Congestive cardiac failure   Lifestyle and Social factors   - Thin - Do not exercise (low vigorous activity, low moderate activity, and do less muscle strengthening) | - Vascular aging | - Screen for and consider appropriate management of cardiovascular disease, vascular risk factors, chronic lung disease, and cancer - Increase exercise - Consider age-appropriate vaccinations |
| PC37F | Early (within 0-5 years)   - Cardiovascular disease - Cancer   Late (within 10-20 years)   - Cancer - Others | Organ impairment   - Visual impairment | - Cancer-related | - Screen for and consider appropriate management of cancer, cardiovascular disease, and visual impairment |
| PC38F | - None | Organ impairment   - Emphysema   Lifestyle and Social factors   - Thin - Cigarette smoking - Low income | - Smoking-related | - Screen for and consider appropriate management of emphysema - Quit smoking |
| PC39F | Late (within 10-20 years)   - Pneumonia - Cardiovascular disease - Others | Vascular risk factors   - Hypertension   Organ impairment   - Chronic lung diseases (asthma, chronic bronchitis, emphysema) - Arthritis   Lifestyle and Social factors   - Obesity - Alcohol use - Do not exercise (low vigorous activity) - Low education - Low income | - Lung disease-related - Inflammation | - Screen for and consider appropriate management of vascular risk factors, chronic lung disease, and arthritis - Manage obesity - Reduce alcohol use - Increase exercise - Consider age-appropriate vaccinations |
